# *De Novo* Exposomic Geospatial Assembly of Chronic Disease Regions with Machine Learning & Network Analysis

**DOI:** 10.1101/2024.07.25.24310832

**Authors:** Andrew Deonarine, Ayushi Batwara, Roy Wada, Shoba Nair, Puneet Sharma, Joseph Loscalzo, Bisola Ojikutu, Kathryn Hall

## Abstract

**Background:** Determining spatial relationships between disease and the exposome is limited by available methodologies. aPEER (algorithm for Projection of Exposome and Epidemiological Relationships) uses machine learning (ML) and network analysis to find spatial relationships between diseases and the exposome in the United States.

**Methods:** Using aPEER we examined the relationship between 12 chronic diseases and 186 pollutants. PCA, K-means clustering, and map projection produced clusters of counties derived from pollutants, and the Jaccard correlation of these clusters with counties with high rates of disease was calculated. Pollution correlation matrices were used together with network analysis to identify the strongest disease-pollution relationships. Results were compared to *LISA*, Moran’s *I,* univariate, elastic net, and random forest regression.

**Findings:** aPEER produced 68,820 maps with human interpretable, distinct pollution-derived regions. Diseases with the strongest pollution associations were hypertension (*J*=0.5316, *p*=3.89×10^-208^), COPD (*J*=0.4545, *p*=8.27×10^-131^), stroke (*J*=0.4517, *p*=1.15×10^-127^), stroke mortality (*J*=0.4445, *p*=4.28×10^-125^), and diabetes mellitus (*J*=0.4425, *p*=2.34×10^-127^). Methanol, acetaldehyde, and formaldehyde were identified as strongly associated with stroke, COPD, stroke mortality, hypertension, and diabetes mellitus in the southeast United States (which correlated with both the Stroke and Diabetes Belts). Pollutants were strongly predictive of chronic disease geography and outperformed conventional prediction models based on preventive services and social determinants of health (using elastic net and random forest regression).

**Interpretation:** aPEER used machine learning to identify disease and air pollutant relationships with similar or superior AUCs compared to social determinants of health (SDOH) and healthcare preventive service models. These findings highlight the utility of aPEER in epidemiological and geospatial analysis as well as the emerging role of exposomics in understanding chronic disease pathology.

**Research in context:** *Evidence before this study:* Many chronic diseases, such as diabetes and stroke mortality, have well defined geographical distributions in the United States. While the reason for these distributions have been actively investigated for decades, limited studies have examined the role of pollution. To assess the current scientific literature available, we completed a structured review in Medline, Google Scholar, and PubMed for any publications in English up to June 24, 2024 using the search terms “stroke”, “cerebral infarction”, “isch(a)emic stroke”, “intracerebral h(a)emorrage”, “h(a)emorrhagic stroke”, or “subarachnoid h(a)emorrage”, “diabetes” AND “Stroke Belt”, “Stroke Region”, “Diabetes Belt”, “Diabetes Region”, or “Disease Belt”. Although there were multiple studies examining the role of genetics and poverty with relation to the geographical distribution of diseases, few examined pollution.

*Added value of this study:* In this study a novel machine learning algorithm was developed which modeled geospatial relationships between chronic disease rates for 3141 counties and county-level pollution measures in the United States. aPEER detected significant relationships between pollutants and several cardiometabolic conditions (using Jaccard correlation coefficient, hypertension (*J*=0.5316, *p*=3.89×10-208), COPD (*J*=0.4545, *p*=8.27×10-131), stroke (*J*=0.4517, *p*=1.15×10-127), stroke mortality (*J*=0.4445, *p*=4.28×10-125), and diabetes (*J*=0.4425, *p*=2.34×10-127)). Using just pollution measures, aPEER consistently identified a region in the southeast United States which correlated closely with both the Stroke and Diabetes Belts, and matched the distribution of multiple cardiometabolic diseases. It was possible to predict the geographical distribution of high chronic disease rates using elastic net and random forest regressions from pollution indicators with similar or superior accuracy (determined by receiver operator curves) compared to preventive healthcare or social determinants of health models.

*Implications of all the available evidence:* For the first time, it was possible to predict hypertension, COPD, stroke mortality, diabetes, and stroke rates from pollution indicators with comparable or superior accuracy compared to conventional models, and readily identify a region of increased pollution in the United States that closely matched the Stroke Belt using machine learning methods. These results highlight the utility of machine learning in exploring and analyzing spatial data, and the importance of pollution in predicting the geographical variation of disease, with implications for cardiometabolic disease pathogenesis and management.

## Introduction

Several diseases follow consistent geographical patterns: stroke, one of the leading causes of mortality in the United States, is geographically associated with a region in the southeast of the country known as the Stroke Belt ^1–5^, and more recently a closely related region with increased diabetes rates (the Diabetes Belt) has been defined ^6^. There is growing evidence that air pollution, in particular, ozone and particulate matter (PM2.5), can also influence the incidence of stroke,^7^ as well as other diseases such as asthma ^8,9^ and diabetes ^10^. To date, examination of geographical disease distributions by population-level variables is limited by the use of conventional statistical techniques, i.e., choropleths ^11,12^, local indicators of spatial association (LISA) ^13^, and the spatial autocorrelation statistic Moran’s *I* ^14^. These techniques cannot systematically examine the geographical associations between chronic disease, population level variables and high-dimensional indicators including air-borne chemicals and water pollutants ^15,16^. Machine learning and network analysis methodologies coupled with the availability of large chronic disease, demographic, and environmental exposure data,^17^ have created an opportunity to investigate more complex spatial relationships between disease and pollution.

While effects of pollution can be relatively small for some conditions, the ubiquity of this exposure elevates the absolute risk at the population level to that of traditional risk factors ^7^. The exposome (defined by Wild *et al.* as the complete set of life-course exposures an individual will encounter ^15,18^) encompasses pollutants that might impact an individual’s health. Understanding the regional links between some measures that comprise the exposome, such as air pollution measures and different chronic diseases could promote informed and targeted interventions and policies to mitigate risk in exposed populations ^19^.

Here, we present a novel, machine-learning pipeline called aPEER (**a**lgorithm for **P**rojection of **E**xposome and **E**pidemiological **R**elationships) that uses unsupervised machine-learning methods to integrate, visualize, and prioritize multiple indicators measured at varying spatial resolutions in a geographically mapped human-interpretable format (Figure 1). Using a combination of principal component analysis (PCA), K-means clustering, geographical projection, correlation and network analysis (using the Jaccard correlation coefficient ^20^) to quantify the correlation between groups of geographical subregions (counties), we identified pollutants in the exposome which were strongly geospatially associated with a disease. Novel disease-pollution relationships were then validated based on their ability to predict the geographical distribution of chronic diseases using elastic net and random-forest models. aPEER identified novel geospatial relationships between multiple chronic diseases and key pollutants that were strongly predictive of chronic disease prevalence. These findings underscore the importance of understanding the potential impact of the hundreds of environmental exposures (collectively referred to as the exposome) on chronic diseases.

**Figure 1.**
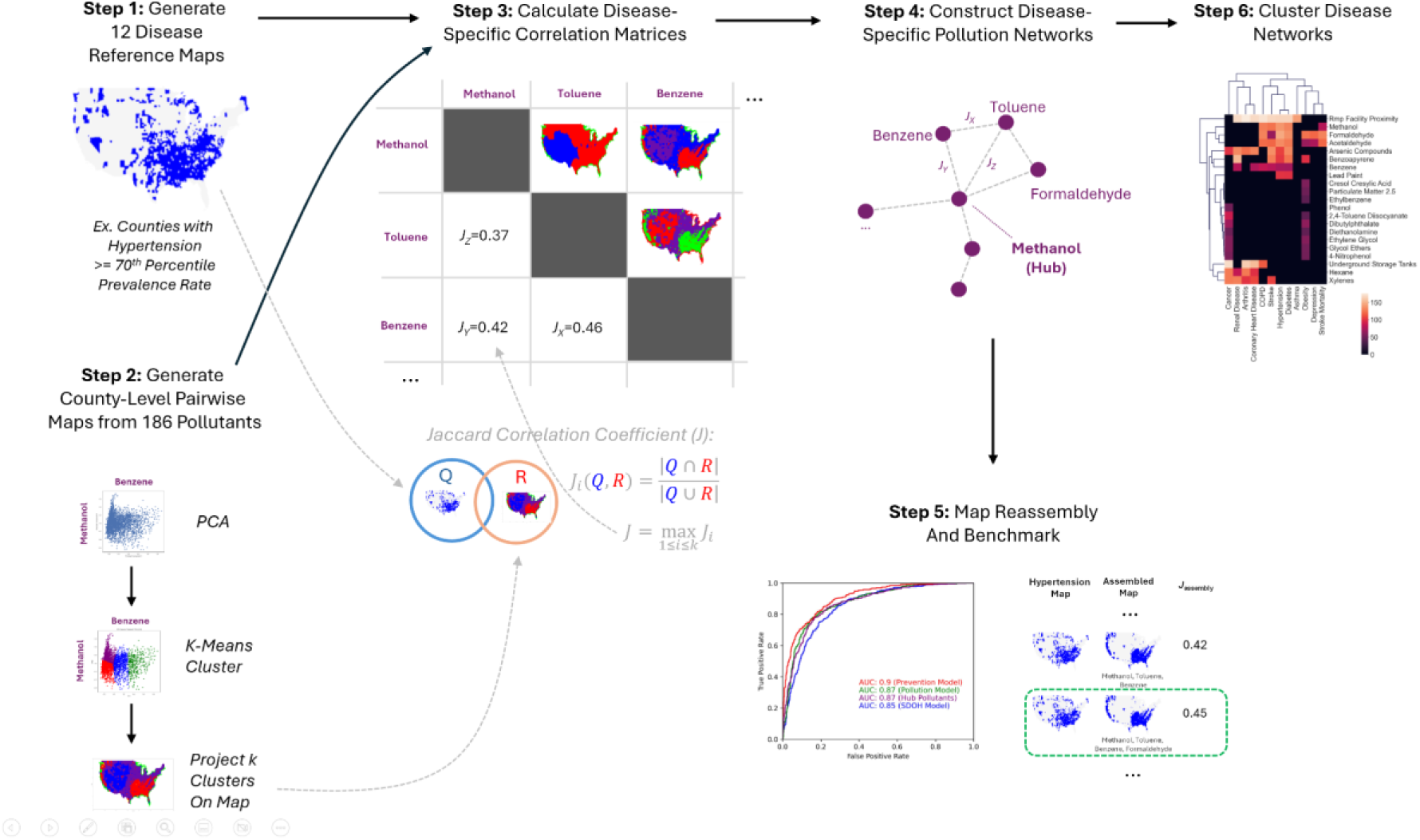
The 6-Step aPEER workflow. Step 1: Generate reference maps of chronic disease prevalence and stroke mortality (≥70th percentile). Step 2: Clusters derived from principal component analysis (PCA) and *k*-means clustering of 186 pollutants projected on a US map. Step 3: Compare disease and pollution maps using Jaccard correlation coefficient (*J*). Step 4: Network analysis used to prioritize strongest relationships and identify key disease-related pollutants. Step 5: Findings benchmarked by examining how closely geographical distribution of key pollutants resembles disease maps. Prediction of disease prevalence by pollutants compared to known predictors like risk factors and SDOH. Step 6: Examine relationships among disease-pollution hubs using hierarchical clustering analysis.

## Materials and Methods

### Overview

The aPEER (**a**lgorithm for **P**rojection of **E**xposome and **E**pidemiology **R**elationships) pipeline was developed to find geographical associations between chronic diseases and the exposome (Figure 1). In Step 1, we generated 12 reference maps of chronic disease prevalence and stroke mortality by selecting counties with chronic disease rates ≥ 70th percentile (we selected 70% as an illustrative example, but also completed analyses at the 60th, 80th and 90th percentiles as part of a sensitivity analysis described below). In Steps 2-4, we sought to find the subset of 186 possible pollutants whose geographical distribution best matched each disease reference map. To derive these pollution-disease relationships, we performed binary space decomposition of the 186 pollutants into pairs, and calculated pollution derived clusters of counties (Step 2) using principal component analysis (PCA), K-means clustering, and map projection for each pair of pollution indicators. We then calculated correlation matrices between these pollution-derived clusters and reference maps using the Jaccard correlation coefficient *J* (illustrated in Step 3 in Figure 1, which measured the correlation between the set of counties in pollution clusters and the counties in disease reference maps). Then we identified key pollutants using network analysis of the correlation matrices (Step 4) which produced lists of hubs (key pollutants) for each chronic disease.

The key pollutants for each chronic disease from Steps 2-4 were validated in Step 5: first, key pollutants (hubs) were used to “assemble” pollution clusters, and pollution-disease pair with the highest *J* correlation coefficients were ranked. Next, we assessed the ability of pollutants to predict the presence of counties with high disease rates using elastic net and random forest regression, and compared model performance to preventive healthcare and SDOH models. We also compared aPEER’s performance to known geospatial analysis methods Moran’s *I* and LISA, as well as a baseline elastic net regression model predicting county-level chronic disease rates from pollutants. Finally, we examined the relationship among disease-pollution hubs using hierarchical clustering analysis of the hubs identified from the networks (Step 6). All analyses and results were presented by following the MI-CLAIM checklist ^21^.

### Databases and Data Sources

The database generated for this study consisted of 226 indicators for 3,141 counties (the complete set of indicators from Center for Disease Control (CDC) PLACES, Environmental Protection Agency’s (EPA) EJSCREEN, and EPA AirToxScreen databases) and integrated into a dataframe in Python (version 3.9) using Pandas (version 1.3.4).

#### Chronic Disease data

Health-related indicators for 3,141 US counties including rates of chronic disease, participation in preventive services, and risk factors were extracted from the Behavioral Risk Factor Surveillance System (BRFSS) and available through the 2023 CDC PLACES database^22^ (Supplementary Table 1). From these datasets we identified 11 disease and health-related measures for analysis (based on the leading contributors to disability-adjusted life years (DALYs) in the United States^23^), specifically, arthritis, asthma, chronic obstructive pulmonary disease (COPD), cancer, coronary heart disease, depression, diabetes, hypertension, obesity, renal disease, and stroke. Stroke mortality data for ages 35 or older was downloaded from the CDC Stroke Death Rates database (between 2017-2019)^24^. High disease prevalence or high stroke-mortality counties were defined as having age-adjusted rates ≥ 70th percentile.

#### Pollution, SDOH, Demographic, and Geographical Data

Pollution data for 9 pollution indicators along with seven social determinants of health (SDOH) / health equity census-tract level measures was extracted from the Environmental Protection Agency (EPA) Environmental Justice (EJSCREEN) 2021 database^25^, together with 177 chemical ambient air concentrations from the EPA’s 2018 AirToxScreen database^26^ reported at the census block group level (in μg/m^3^), and calculated at the county level by population-weighting the census block group level exposures and then calculating the sum for each county from the blocks. Together, the EJSCREEN and AirToxScreen measures resulted in 186 pollution measures examined in this study. Geographical boundary information for counties, in the form of GeoJSON, were obtained from the US Census TIGER database^27^. The 9 EJSCREEN pollution indicators ^28^ included particulate matter 2.5 (PM2.5; µg/m^3^), ozone (parts per billion), traffic proximity (vehicles per day / meters), lead paint exposure (% of housing units built before 1960), superfund proximity (superfund site count / km), RMP facility proximity (facility count / km), hazardous waste proximity (count of hazardous waste facilities within 5 km (or nearest beyond 5 km), each divided by distance in kilometers), underground storage tanks (count of facilities (multiplied by a factor of 7.7) within a 1,500-foot buffered block group), and wastewater discharge (modeled toxic concentrations at stream segments within 500 meters, divided by distance in kilometers (km)) (Supplementary Table 1). The year of pollution exposure was selected to precede the year when chronic disease rates were reported.

### Machine Learning Analyses

The mean and median of all county-level variables (Table 1) from CDC PLACES, and EJSCREEN databases were calculated using the *NumPy* library (version 1.20.3). As a baseline model to benchmark aPEER, we conducted univariate and multivariate elastic net regressions (with 12 different disease-related measures as dependent variables, and county-level SDOH, pollution, and other indicators as independent variables) at the county level using the *statsmodels* library (version 0.12.1). A full description of this analysis is contained in Supplemental Methods Section 1.

**Table 1.** County-level descriptive statistics for chronic disease and healthcare, pollution indicators and demographic data for 3141 counties used in this study (N = number of counties equal to or above a percentile cutoff; * = chronic diseases/indicators that are modeled in this study).

|  | Mean | Std. Deviation | 60% ile | N | 70% ile | N | 80% ile | N | 90% ile | N |
| --- | --- | --- | --- | --- | --- | --- | --- | --- | --- | --- |
| **EPA EJSCREEN Demographics (%)** |  |  |  |  |  |  |  |  |  |  |
| Minority | 23.75 | 20.22 | 22.84 | 1257 | 30.91 | 943 | 40.256 | 629 | 54.78 | 315 |
| Low Income | 35.58 | 9.97 | 37.65 | 1257 | 40.84 | 943 | 43.833 | 629 | 48.30 | 315 |
| Less than HS Education | 13.14 | 6.32 | 13.50 | 1257 | 15.58 | 943 | 18.161 | 629 | 21.37 | 315 |
| Linguistic Isolation | 1.88 | 3.20 | 1.21 | 1257 | 1.70 | 943 | 2.558 | 629 | 4.59 | 315 |
| Under 5 yrs | 5.82 | 1.26 | 5.99 | 1257 | 6.27 | 943 | 6.590 | 629 | 7.12 | 315 |
| Over 64 yrs | 18.79 | 4.66 | 19.44 | 1257 | 20.54 | 943 | 22.017 | 629 | 24.71 | 315 |
| Unemployed | 5.36 | 2.73 | 5.57 | 1257 | 6.19 | 943 | 7.036 | 629 | 8.43 | 315 |
| **EPA EJSCREEN Pollution Measures** |  |  |  |  |  |  |  |  |  |  |
| Lead Paint | 0.29 | 0.15 | 0.31 | 1257 | 0.37 | 943 | 0.43 | 629 | 0.50 | 315 |
| Diesel Particulate Matter | 0.11 | 0.08 | 0.11 | 1257 | 0.13 | 943 | 0.15 | 629 | 0.20 | 315 |
| Air Toxic Cancer Risk | 21.23 | 11.21 | 23.49 | 1257 | 30.00 | 958 | 30.00 | 788 | 30.00 | 315 |
| Air Toxic Respiratory Index | 0.27 | 0.15 | 0.30 | 1440 | 0.33 | 943 | 0.40 | 666 | 0.43 | 315 |
| Traffic Proximity | 38.26 | 157.16 | 0.00 | 3141 | 0.00 | 3141 | 13.44 | 629 | 103.57 | 315 |
| Wastewater discharge | 0.08 | 1.35 | 0.00 | 3141 | 0.00 | 3141 | 0.00 | 3141 | 0.00 | 315 |
| Superfund Proximity | 0.06 | 0.10 | 0.03 | 1257 | 0.04 | 943 | 0.08 | 629 | 0.16 | 315 |
| RMP Facility Proximity | 0.52 | 0.55 | 0.48 | 1257 | 0.65 | 943 | 0.89 | 629 | 1.30 | 315 |
| Hazardous Waste Proximity | 0.44 | 1.00 | 0.26 | 1257 | 0.40 | 943 | 0.61 | 629 | 1.05 | 315 |
| Ozone | 38.32 | 12.68 | 42.02 | 1257 | 43.24 | 943 | 44.39 | 629 | 47.67 | 315 |
| Particulate Matter 2.5 | 7.13 | 2.57 | 8.18 | 1257 | 8.58 | 943 | 8.91 | 629 | 9.30 | 315 |
| Underground Storage Tanks | 1.52 | 2.04 | 1.30 | 1257 | 1.78 | 943 | 2.38 | 629 | 3.38 | 315 |
| **CDC PLACES Health Measures (%)** |  |  |  |  |  |  |  |  |  |  |
| Healthcare Access | 16.14 | 6.55 | 16.1 | 1260 | 17.8 | 950 | 20 | 633 | 24.5 | 316 |
| *Arthritis | 29.28 | 4.69 | 30.5 | 1257 | 31.7 | 962 | 33.2 | 635 | 35.2 | 321 |
| Binge Drinking | 15.72 | 2.71 | 16.2 | 1295 | 17 | 961 | 18 | 629 | 19.3 | 324 |
| *Hypertension | 36.91 | 6.47 | 38.1 | 1273 | 39.8 | 954 | 42.1 | 634 | 45 | 317 |
| BP Medication | 76.83 | 7.32 | 78.7 | 1296 | 79.5 | 979 | 80.3 | 645 | 81.4 | 318 |
| *Cancer | 7.55 | 1.16 | 7.8 | 1380 | 8.1 | 1024 | 8.5 | 631 | 9 | 328 |
| *Asthma | 9.95 | 0.92 | 10.1 | 1337 | 10.4 | 983 | 10.7 | 679 | 11.2 | 318 |
| Cervical Cancer Screen | 80.84 | 2.49 | 81.6 | 1279 | 82.2 | 987 | 82.9 | 656 | 83.9 | 318 |
| *Coronary Heart Disease | 8.20 | 1.58 | 8.6 | 1328 | 9.1 | 945 | 9.5 | 656 | 10.1 | 346 |
| Annual Checkup | 75.43 | 3.92 | 76.9 | 1257 | 77.7 | 951 | 78.6 | 634 | 79.8 | 328 |
| Cholesterol Screen | 84.91 | 7.46 | 86.2 | 1300 | 86.8 | 981 | 87.6 | 667 | 88.7 | 325 |
| Colon Cancer Screen | 70.34 | 4.62 | 71.9 | 1258 | 73.1 | 950 | 74.4 | 638 | 75.9 | 321 |
| *COPD | 8.64 | 2.24 | 9 | 1309 | 9.6 | 989 | 10.4 | 664 | 11.6 | 331 |
| Core Male Prevention | 42.36 | 5.14 | 43.4 | 1278 | 44.8 | 945 | 46.5 | 631 | 49 | 328 |
| Core Female Prevention | 36.71 | 4.48 | 37.7 | 1261 | 38.9 | 945 | 40.4 | 633 | 42.5 | 315 |
| Smoking | 18.92 | 3.97 | 19.7 | 1275 | 20.8 | 947 | 22 | 653 | 24 | 319 |
| Dental Services | 59.70 | 7.52 | 62.6 | 1262 | 64.4 | 949 | 66.3 | 644 | 68.8 | 317 |
| *Depression | 21.09 | 3.15 | 21.9 | 1270 | 22.8 | 951 | 23.9 | 635 | 25.4 | 324 |
| *Diabetes | 12.73 | 2.62 | 13.1 | 1265 | 13.9 | 947 | 14.9 | 629 | 16.2 | 322 |
| Poor General Health | 17.63 | 4.70 | 18.3 | 1257 | 20 | 945 | 21.8 | 629 | 24 | 315 |
| High Cholesterol | 34.98 | 4.49 | 36.3 | 1268 | 37.2 | 956 | 38.2 | 630 | 39.5 | 322 |
| *Renal Disease | 3.51 | 0.59 | 3.6 | 1399 | 3.8 | 972 | 4 | 667 | 4.3 | 338 |
| Limited Phys. Activity | 27.02 | 5.32 | 28 | 1272 | 29.8 | 947 | 31.6 | 634 | 34 | 324 |
| Mammogram | 70.52 | 3.95 | 71.7 | 1282 | 72.7 | 977 | 73.9 | 661 | 75.3 | 326 |
| Poor Mental Health | 14.65 | 2.03 | 15.3 | 1285 | 15.8 | 996 | 16.4 | 658 | 17.2 | 327 |
| *Obesity | 35.97 | 4.67 | 37.4 | 1271 | 38.4 | 972 | 39.6 | 650 | 41.2 | 325 |
| Poor Physical Health | 12.05 | 2.38 | 12.5 | 1276 | 13.2 | 958 | 14 | 648 | 15.2 | 319 |
| Sleep | 33.36 | 3.70 | 34.2 | 1291 | 35.2 | 971 | 36.5 | 635 | 38.2 | 318 |
| *Stroke | 3.87 | 0.86 | 4 | 1290 | 4.2 | 1034 | 4.5 | 706 | 5 | 350 |
| Tooth Loss | 13.18 | 3.80 | 13.6 | 1273 | 14.8 | 951 | 16.4 | 635 | 18.3 | 319 |
| **CDC Stroke Data (Rate)** |  |  |  |  |  |  |  |  |  |  |
| *Stroke Mortality | 39.32 | 8.85 | 40.6 | 1267 | 43.1 | 949 | 46 | 633 | 50.3 | 316 |

To identify and refine the strongest geographical disease-pollution associations, aPEER used a 6-step process (Figure 1):

## Step 1 - Generate 12 Disease Reference Maps

The 12 county-level disease reference maps of counties with disease prevalence rates ≥ 70th percentile were generated for each disease. National maps of the mainland United States were then generated at the county level using GeoJSON derived from the TIGER database, and the *PolygonPatch* function from the *descartes* (version 1.1.0) library.

## Step 2 - Generate County-Level Pairwise Maps from 186 Pollutants

We grouped counties in the United States into distinct clusters by calculating principal component analysis (PCA) with the two normalized pollution measures for all pairwise combinations of 186 pollutants (17,205 combinations). We then calculated K-means clustering (varying the value *k* = {2,3,4,5}) using the first two principal components for each pair, which produced a set of clusters of counties (using the *PCA* and *KMeans* functions in the *sklearn* library version 0.24.2, see Supplementary Methods Part 2 for more details). This resulted in 68,820 sets of county clusters, with each set corresponding to a pair of pollutants and a specific value *k*. Each set of clusters was then projected onto a map of the US (using *PolygonPatch* in the *descartes* library (version 1.1.0)).

## Step 3 - Calculate Disease-Specific Correlation Matrices

We then identified which clustered maps (created from pairs of pollutants) were most similar to each of the 12 disease reference maps. To assess this similarity, we calculated the Jaccard correlation coefficient *J* which measures the similarity between the sets of counties defined by chronic diseases, as well as the sets of counties in each of the different clusters calculated from pairs of pollutants. *J* is defined as the number of items in the intersection between two sets, divided by the number of items in the union of two sets ^20^. To calculate *J* for a disease reference map and pollution pairs (with the multiple pollution clusters generated by K-means clustering with *k* = {2,3,4,5}) we calculated all possible *J* values for disease-pollution clusters, and then identified the maximum *J* value as defined in equation 1:

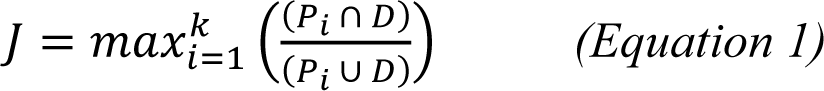

where *P*_i_ are counties in a cluster, and *D* the set of counties with disease prevalence ≥ 70th percentile, and *i* is a specific cluster *k*. A Fisher Exact Test was used to derive the *p*-value for each Jaccard correlation coefficient, and the *p*-value was designated as statistically significant using a Bonferroni-adjustment. We then generated a pairwise pollution correlation matrix for each disease resulting in 12 correlation matrices of Jaccard correlation coefficients (one for each disease) which is fully described in Supplemental Methods Section 2.

## Step 4 - Construct Disease Specific Pollution Networks

To identify pollutants most strongly associated with a disease, a network was constructed from the matrix of Jaccard correlation coefficients with *p* << 0.001 after Bonferroni adjustment. In this network, nodes constituted pollution indicators, and edges represented a statistically-significant Jaccard correlation coefficient associated with a pair of pollutants (nodes) for a disease. The length of each edge was scaled inversely proportional to the value of the Jaccard similarity coefficient. Hubs were then identified based on the number of connections (degree) for each node (see Supplemental Methods Section 3). Networks were calculated using the *networkx* library (version 2.6.3) in Python, and statistics including network density, diameter, and triadic closure were calculated using the *networkx* density, diameter, and transitivity functions, respectively. A full description of the network construction is detailed in Supplemental Methods Section 3.

## Step 5 - Map Assembly and Benchmark

We then validated the pollution hub-disease relationships using elastic net and random forest regression (to determine if the geographical distribution of a disease could be predicted with high accuracy from the hub pollutants), as well as performing a process of map assembly, which involves creating geographical clusters from the hub pollutants that match the disease reference maps.

To calculate elastic net and random forest regressions, we defined the dependent variable *y* as any county that was identified in a disease reference map as having a disease prevalence or stroke mortality rate ≥ 70th percentile as having a value of *1*, or *0* if it did not meet this rate cutoff. We then defined four models based on different combinations of independent variables: a “pollution” model consisting of all 186 county-level pollutants as independent variables, a “hub” model consisting only of the county-level hub pollutants, a “SDOH” (social determinants of health) model consisting of 3 county-level measures, including percent minority, low income, and less than high school education, and a “prevention” model consisting of the county level rates of mammograms, core female prevention services, core male prevention services, dental services, and annual checkup rates. Elastic net regression was completed using the *LogisticRegressionCV* function in *sklearn* (see Supplemental Methods Section 4 for full details), while random forest models were developed using the *xgboost* library (see Supplemental Methods Section 5 for full details). We also examined if known geospatial analysis techniques, namely Moran’s *I* and LISA using the *splot* library (version 1.1.5)) could detect the clusters identified by aPEER (see Supplemental Methods Section 6 for full details).

We then assessed if hub pollutants produced maps that resembled chronic diseases through a process of map “assembly”. In map assembly, the pair of hub pollutants with the highest Jaccard correlation coefficient (from the correlation matrices calculated previously) were identified, and the cluster of counties corresponding to this highest value were plotted on a map (“assembled”) for comparison to the 12 disease reference maps. We then ranked the pollution pair-disease relationships by Jaccard correlation coefficient to identify the strongest associations between the set of pollution cluster counties and the set of high disease rate counties.

## Step 6 - Cluster Disease Networks

We examined the relationships between diseases and pollutants by clustering the pollution hub degrees for each of the 12 chronic diseases measures using the clustermap feature in the seaborn library (version 0.11.2). For comparison, we also clustered the top 5 statistically significant ꞵ coefficients from elastic net regression, and top 5 feature importance rank values from random forest regression, and compared the clustering pattern across the three different disease-pollution clustering approaches.

Given that this analysis was performed with counties that had chronic disease prevalences ≥ 70th percentile, we examined the robustness of the disease-pollution associations by varying the percentile cutoff and repeating steps 1-6 at the 60th, 80th, and 90th percentiles. We then calculated the resulting disease-pollution associations and clustered them, and examined the resulting patterns across the different cutoffs.

### Ethics Approval

Ethics approval was not required for this investigation because the data was publicly available, as no individual-level data was incorporated and small re-identifiable populations were not identified.

### Code and Data Availability

The aPEER core algorithm, which creates clustered, color-coded maps from high dimensional data, can be downloaded from: https://github.com/adeonarinebphc/apeer/.

## Results

In Table 1, the descriptive statistics of the county-level data used in this study are described, highlighting measures from EPA’s EJSCREEN and CDC PLACES data. We then identified counties with chronic disease rates ≥ 70% (resulting in about 950 counties for each disease, the resulting maps for each disease are depicted in Supplementary Figure 1). In Supplementary Table 1, baseline univariate and multivariate elastic net regressions are presented, with the top beta coefficients listed for the 12 chronic disease measures. The highest multivariate beta coefficients include those for carbon tetrachloride in the obesity model (β = 1199.37, *p* = 2.49×10^-17^) and in the arthritis model (β = 724.12, *p* = 4.59×10^-7^) and that for formaldehyde with stroke mortality (β = 565.81, *p* = 2.77 x 10^-7^).

In Figure 2, example correlation matrices depicting the geospatial relationships between pairs of pollutants and hypertension (Figure 2A) and of pollutants and stroke mortality (Figure 2B) are illustrated (hypertension and stroke mortality were later found to be among the highest disease-pollution associations determined by aPEER using map assembly). Acetaldehyde and formaldehyde had many of the highest associations, with the highest correlations found with pollution pairs acetaldehyde-benzo(a)pyrene (*J* = 0.5315, *p* << 0.01), formaldehyde-diesel PM (*J* = 0.5307, *p* << 0.01), and acetaldehyde-1,3-butadiene (*J* = 0.5274, *p* << 0.01), while a similar pattern of strong associations was found with acetaldehyde and formaldehyde and stroke mortality (Figure 2B), with the highest associations being benzo(a)pyrene-acetaldehyde (*J* = 0.4579, *p* << 0.01) and formaldehyde-benzene *J*=0.4578, p << 0.01). The correlation matrices for the remaining chronic disease indicators are presented in Supplementary Figure 2.

**Figure 2.**
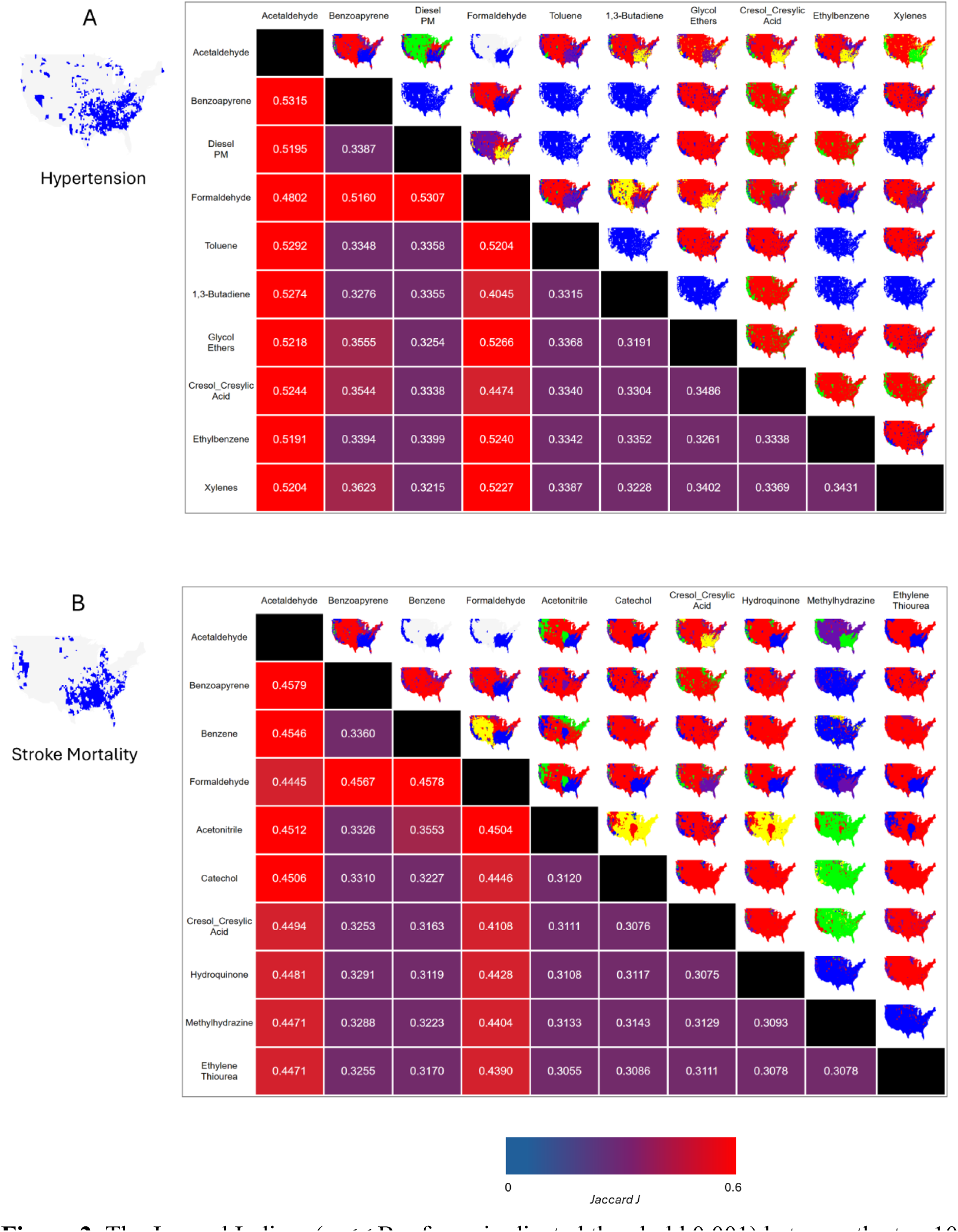
The Jaccard Indices (*p* << Bonferroni-adjusted threshold 0.001) between the top 10 pollutants (ranked by Jaccard *J*) for (A) hypertension and (B) stroke mortality (map colors are arbitrary).

Using the correlation matrices, we then calculated pollution networks for each disease and identified the hubs in the network, with Figure 3 presenting the networks for hypertension (Figure 3A) and stroke mortality (Figure 3B), together with elastic net and random forest models predicting disease geography from the hub pollutants. Methanol, acetaldehyde, and formaldehyde were identified as hubs in both hypertension (8 hubs) and stroke mortality (3 hubs). Validating the pollution hubs using elastic net and random forest models revealed very specific patterns in the area under the curve (AUCs), with the prevention model performing the best in elastic net models for hypertension (AUC = 0.9) and stroke mortality (AUC = 0.8), while the pollution model (consisting of all 186 pollutants) performed best (AUC = 0.93 and AUC = 0.87) for hypertension and stroke mortality with random forest models (Figure 3). The hub pollutant models consistently outperformed the SDOH models irrespective of method for both hypertension and stroke mortality (AUC = 0.79-0.9). In general, the pollution model outperformed all other models when predicting the geographical distribution of the other chronic diseases, especially with random forest models (see Supplementary Figure 3), with the highest AUC noted for depression (AUC = 0.94 (random forest), AUC = 0.81 (elastic net)) followed by hypertension (AUC = 0.93 (random forest), AUC = 0.87 (elastic net)). The calibration curves for the various elastic net and random forest models are presented in Supplemental Figure 4, with higher accuracy in general found for random forest models.

**Figure 3.**
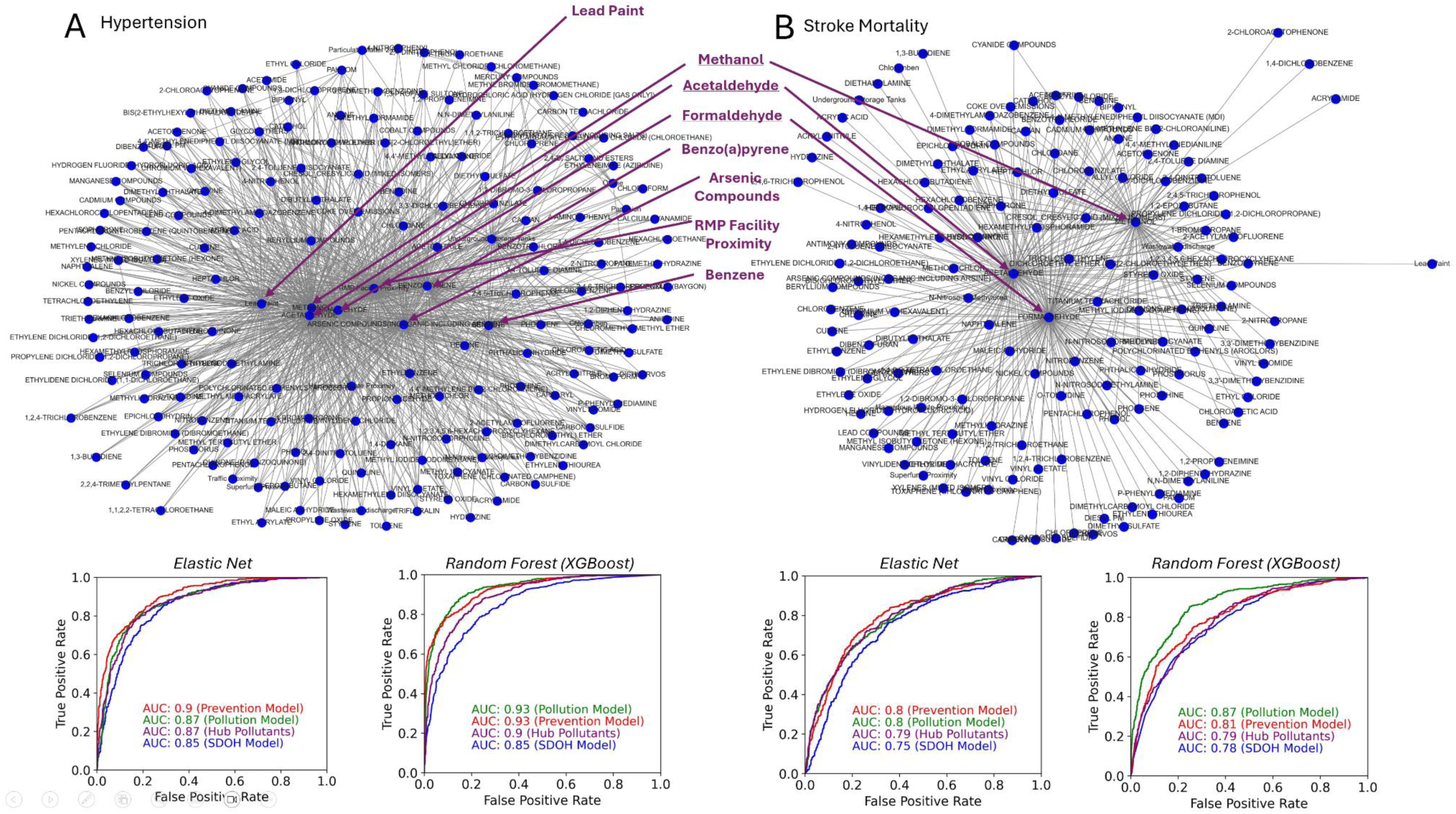
Pollution networks for (A) hypertension and (B) stroke mortality, with elastic net and random forest models predicting the geographical distribution of a given disease using pollution hubs compared to SDOH, prevention, and pollution (all pollution features) models.

After identifying pollution hubs, we compared them to the beta coefficients from elastic net and important features derived from random forest (Figure 4). We found that formaldehyde, acetaldehyde, and methanol were consistently highly predictive of hypertension and stroke mortality. The ordering of formaldehyde, acetaldehyde, and methanol was very similar between the aPEER pollution hubs and the beta coefficients derived by elastic net (Supplementary Figure 4, and calibration curves for elastic net and random forest regression are depicted in Supplementary Figure 5). Formaldehyde, acetaldehyde, and methanol were also found to be among the top 10 pollution hubs for COPD, depression, diabetes, and stroke, (see Supplementary Figure 6).

**Figure 4.**
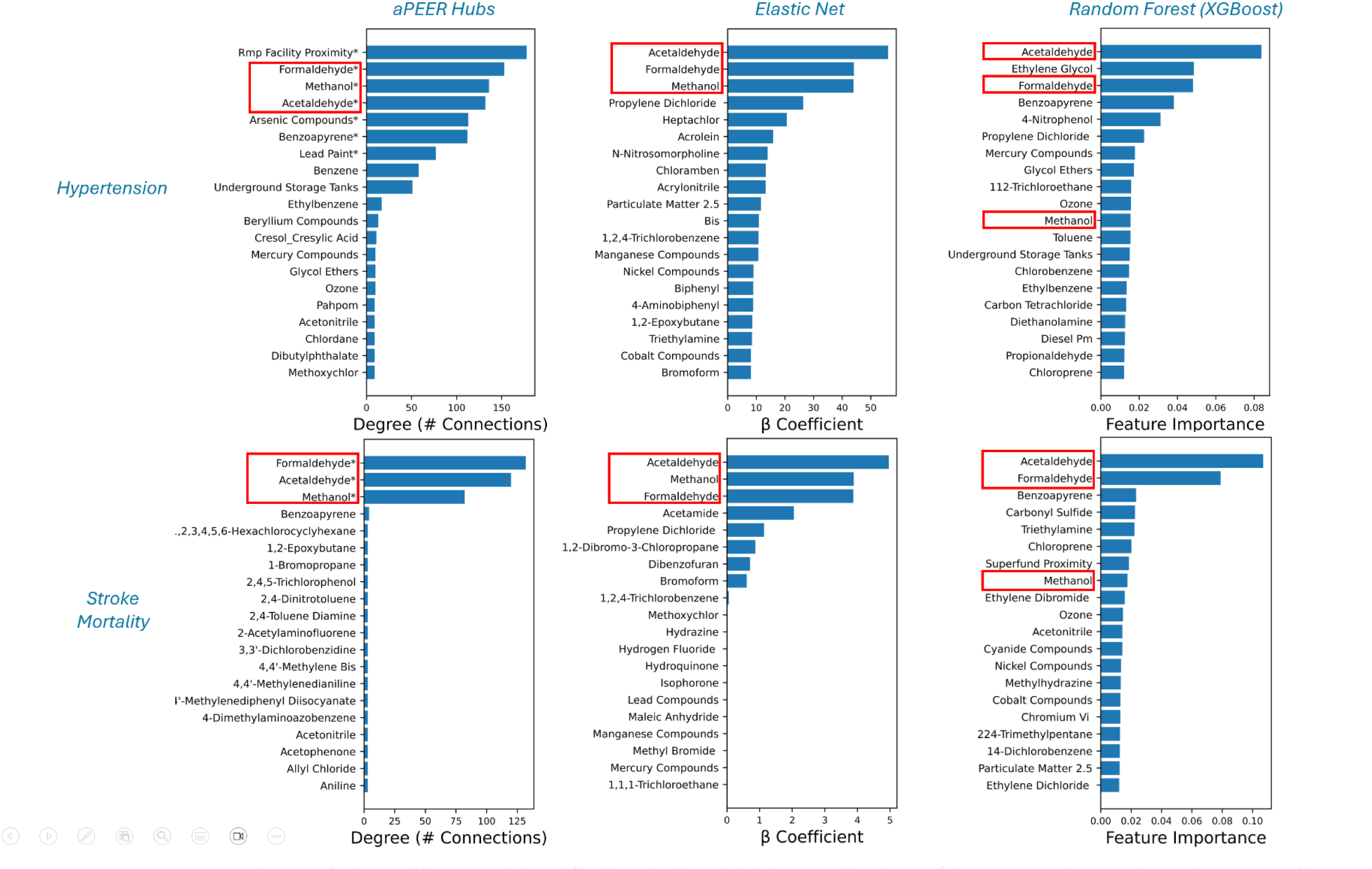
A comparison of the pollutants identified as being highly predictive of hypertension and stroke mortality using aPEER (pollution hubs), elastic net (β coefficients), and random forest models (importance). Three pollutants (formaldehyde, methanol, and acetaldehyde) consistently appeared irrespective of the analysis method employed (highlighted in red).

We then determined the strongest disease-pollution associations by assembling pollution maps from pairs of pollutants and ranking the associations by Jaccard correlation coefficient (Figure 5A), and clustered aPEER pollution hub degree, pollution-related elastic net β coefficients, and random forest pollution-associated feature importance values (Figure 5B). Four out of the five top assembled map-chronic disease relationships were related to cardiometabolic conditions, including the acetaldehyde-benzo(a)pyrene pollution pair for hypertension (*J*=0.5316, *p*=3.89×10^-208^), formaldehyde-glycol ether for COPD (*J*=0.4545, *p*=8.27×10^-131^), acetaldehyde-benzo(a)pyrene for stroke (*J*=0.4517, *p*=1.15×10^-127^), acetaldehyde-formaldehyde for stroke mortality (*J*=0.4445, *p*=4.28×10^-125^), and acetaldehyde-benao(a)pyrene for diabetes (*J*=0.4425, *p*=2.34×10^-127^) (Figure 5A). In Figure 5B, a consistent pattern of formaldehyde, acetaldehyde, and methanol clustering together is apparent; these pollutants also clustered together using β coefficients from elastic net. These relationships were partly noted after clustering feature importance from random forest analysis, with acetaldehyde clustering separately from methanol and formaldehyde (the full list of disease-pollution associations is presented in Supplementary Figure 7). To assess the robustness of the pollution-disease associations noted with aPEER, we completed a sensitivity analysis by varying the chronic disease cutoff, using the ≥ 60th, ≥ 70th, ≥80th, and ≥ 90th percentiles (Supplementary Figure 8). Clustering the results from aPEER and elastic net results mostly showed acetaldehyde, formaldehyde, and methanol grouping together at the ≥ 70th percentile cutoff, while less consistent results were noted with random forest regression, suggesting that aPEER and elastic net may be methodologically similar.

**Figure 5.**
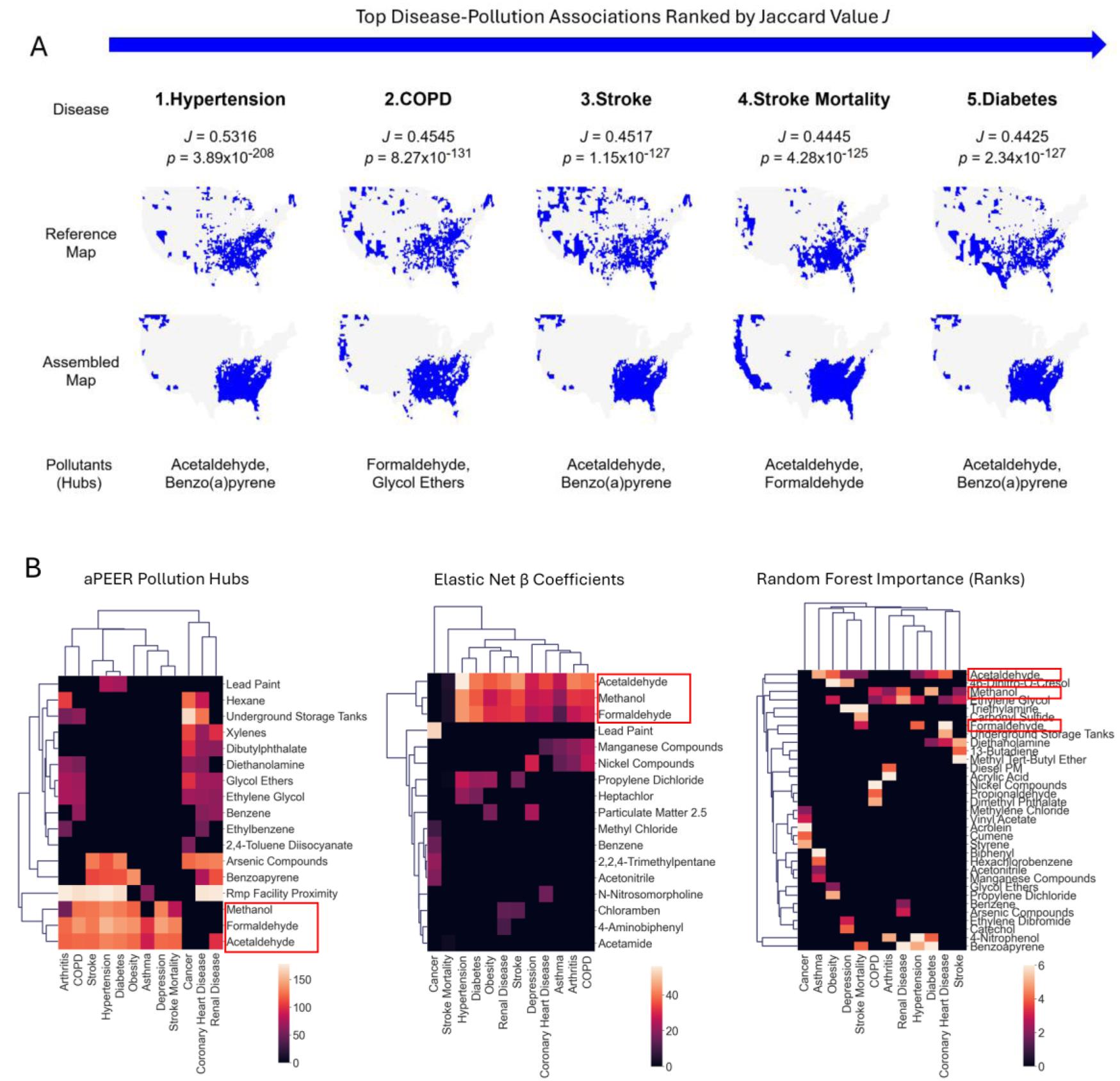
(A) The top 5 disease-pollution associations derived from assembled pollution maps (70th percentile) ranked by Jaccard correlation coefficients, and (B) clustered heatmap of aPEER pollution hubs, elastic net ꞵ coefficients, and random forest importance features showing methanol, formaldehyde, and acetaldehyde closely clustered together, and strongly associated with multiple cardiometabolic diseases (highlighted in red).

We compared the findings from aPEER with Moran’s *I* and LISA, and found that neither of these methods identified statistically significant geographical patterns in pollution or selected diseases (Supplementary Figure 9 and 10), indicating that aPEER may be more robust when detecting geospatial patterns in pollution data. We also examined if the disease-pollution relationships identified by aPEER were confounded by population levels, but no significant relationships were noted with selected diseases and pollutants (Supplementary Figure 11).

Additionally, we examined if aPEER was identifying the similarities between disease and pollution distributions, but no distributional similarities were apparent (Supplementary Figure 12). The pollutants identified by aPEER as important (hubs) were not identified in the original baseline elastic net model, highlighting the limitations of the baseline model.

## Discussion

In this investigation, it was possible to identify novel geospatial disease-pollution relationships using the aPEER algorithm between 12 chronic disease indicators and 186 pollutants, particularly between hypertension, diabetes, stroke mortality, and stroke and the pollutants acetaldehyde, formaldehyde, and methanol (Figure 5). The associations between acetaldehyde, formaldehyde, and methanol and cardiometabolic diseases identified through correlation matrices (Figure 2) and network analysis (Figure 3) was confirmed by elastic net and random forest regression (Figure 4), while statistically significant geographical distributions of diseases were not noted using conventional methods such as Moran’s *I*, LISA, or benchmark univariate / elastic net regression models. These associations were also persistent even when we performed a sensitivity analysis varying the cutoffs from the 60th-90th percentile, with consistent clustering of acetaldehyde, formaldehyde, and methanol prominently noted at the 70th percentile in the aPEER and elastic net regression analysis (Supplementary Figure 8), suggesting that at and above this threshold pollutants begin to play a significant role in disease prevalence for several cardiometabolic conditions. To our knowledge, this is the first time that air pollutants were found to be better at predicting cardiometabolic disease than conventional models based on healthcare system measures and the SDOH. The fact that aPEER generated a region from the exposome (especially acetaldehyde, formaldehyde, and methanol) that strongly resembled both the Stroke Belt and Diabetes Belts provides strong evidence for a potential linkage between stroke mortality, hypertension, diabetes, stroke and other cardiometabolic conditions and these pollutants. From the results of this study, three main conclusions can be drawn.

Firstly, aPEER identified a region in the southeast United States defined by hub pollutants which is roughly correlated with both the Stroke Belt and Diabetes Belts (Figure 5A), and was highly associated with stroke, COPD, diabetes, hypertension, and stroke mortality. Partial explanations for regional variations in chronic diseases focus on risk factors, comorbidities, lifestyle, and SDOH factors together with the impacts of structural and environmental racism ^1,29,30^. As well, aPEER identified regions on the west coast that had high rates of stroke mortality, which are not identified using the conventional Stroke Belt definition. Air pollution measures such as diesel particulate matter and PM2.5 are known to contribute to inflammation and stroke, and this may be one of the major pathways through which air pollution results in an increase in stroke rates ^31–34^. Many of the air pollutants identified by aPEER, particularly acetaldehyde, formaldehyde, and methanol, have known links to chronic diseases. For instance, formaldehyde has been associated with stroke mortality ^35^, and hypertension ^35,36^, but there have been fewer studies characterizing these relationships in the United States. It is possible that these pollutants directly contribute to the pathogenesis of cardiometabolic diseases. Another pathway may be indirect, where air pollutants contribute to risk factors for stroke and diabetes ^37–41^. More recently, an investigation found an association between organic aerosols and the Stroke Belt ^42^, and recapitulated very similar results to those found in this investigation. Importantly, in contrast to the observational associations observed by Pye *et al.* ^42^, our analysis uniquely demonstrated that it is possible to assemble the Stroke Belt from hub pollutants (Figure 5B), and that these pollutants perform nearly the same or better than established preventive services and SDOH reference models.

The ability of aPEER to produce explainable, human-interpretable maps from simple pollution combinations partly addresses the explainable artificial intelligence (XAI) ^43^ problem of other machine learning techniques such as elastic net regression and random forest regressions, which rely on “black-box” coefficient optimization and creation of abstract decision trees, respectively. In the large correlation matrix of clustered maps there are several unique maps with distinct geographical distributions, and aPEER could be used to better understand how climate change, pollution, and features such as geographical elevation (which may be associated with some of the clusters) are correlated with disease distribution. aPEER can be used as a tool to rapidly explore geospatial ecological correlations and generate hypotheses in epidemiological studies involving high dimensional data.

Secondly, previous investigations have identified PM2.5 ^44,45^, ozone ^46^, and selenium (deficiency) ^1^ (and have ruled out mercury ^47^) as being associated with increased Stroke Belt stroke rates, but few significant environmental predictions or associations have been otherwise noted. Additionally, previous studies focused on SDOH/equity factors (and in particular the African American population) and the possible cultural and genetic causes ^48^ of increased stroke ^2,29^; by contrast, this investigation identified modifiable environmental factors that comprise the exposome, in particular air pollution, that might further explain this risk. Our results may indicate that issues such as environmental racism and exposure to specific compounds should be prioritized for investigation and intervention not only for stroke mortality, but also for general life expectancy and other chronic diseases with high AUCs (see Supplementary Figure 4A and 4B).

Thirdly, this investigation highlights the role of unsupervised machine learning in analyzing geographical information and finding associations between different indicators. By combining dimensionality reduction, clustering, and regression analysis for validation, it was possible to detect associations between pollution indicators and chronic diseases that would not normally be detectable. For instance, using an elastic net regression model to predict chronic disease rates from 186 different pollutants identified different pollutants compared to aPEER (except for stroke mortality, where formaldehyde and methanol were found to be significant). This observation may partly explain why pollution indicators have not been extensively studied previously for different chronic diseases. For example, Ji *et al*. used a combination of machine learning and multilevel modeling to analyze environmental and SDOH associations with stroke, and identified ozone as having a strong association. While the relationship with ozone was replicated in our analysis, it did not appear to be the strongest relationship ^46^. This difference in outcome may be partly due to the data employed by Ji *et al.,* a study that used CDC 500 Cities data, which is a subset of the CDC PLACES data used here.

Discovering relationships between multiple diseases and the exposome was not possible using conventional methods such as baseline elastic net regression, LISA, or Moran’s *I*, highlighting aPEER’s utility as a novel geospatial analysis tool. aPEER is not limited to pollution or stroke data, and can be extended to include other exposomic or geospatial data, and the association of those data with the geographical distribution of other chronic diseases. In addition, aPEER produces clearly demarcated cluster boundaries, which reduces the need for arbitrary thresholds that sometimes are used to identify geographical regions. Hence, aPEER could be used as a general epidemiological tool to investigate the geographical relationships between different geospatial measurements (such as pollution and disease rates) at different geographical resolutions (such as counties, census-tracts, zip codes, census blocks, and precincts). This method could be further enhanced through the incorporation of satellite imagery to understand better how the built environment could enhance the prediction of disease rates; in this vein, we are investigating whether different correlations (such as an area-weighted Jaccard correlation coefficient or tetrachoric correlation) and different clustering methods (such as generating pairwise disease maps and using 1-dimensional clustering algorithms) would yield better results.

Limitations of this investigation include the ecological nature of the data and relationships examined: although different geographical resolutions were used and were found to be concordant, these relationships should be confirmed using individual-level diagnosis of different chronic diseases and exposures to air pollution and other pollution indicators. Additionally, this modeling work was completed in the United States, and generalizability to other countries is yet to be determined.

In summary, using aPEER, a novel machine learning algorithm designed to investigate spatial relationships between pollutants and stroke, we identified key pollutants associated with multiple chronic diseases, such as stroke, hypertension, and diabetes, and life expectancy. For the first time, it was possible to identify pollutants that predicted the geospatial distribution of chronic diseases with higher accuracy than conventional preventive and SDOH factors, highlighting the importance of the exposome in the pathogenesis of multiple chronic diseases, and the role that modifiable environmental exposures play in disease.

## Data Availability

The database generated for this study consisted of 226 indicators for 3,141 counties (the complete set of indicators from Center for Disease Control (CDC) PLACES, Environmental Protection Agency's (EPA) EJSCREEN, and EPA AirToxScreen databases) and integrated into a dataframe in Python (version 3.9) using Pandas (version 1.3.4). Chronic Disease data: Health-related indicators for 3,141 US counties including rates of chronic disease, participation in preventive services, and risk factors were extracted from the Behavioral Risk Factor Surveillance System (BRFSS) and available through the 2023 CDC PLACES database22 (Supplementary Table 1). From these datasets we identified 11 disease and health-related measures for analysis (based on the leading contributors to disability-adjusted life years (DALYs) in the United States23), specifically, arthritis, asthma, chronic obstructive pulmonary disease (COPD), cancer, coronary heart disease, depression, diabetes, hypertension, obesity, renal disease, and stroke. Stroke mortality data for ages 35 or older was downloaded from the CDC Stroke Death Rates database (between 2017-2019)24. High disease prevalence or high stroke-mortality counties were defined as having age-adjusted rates > = 70th percentile. Pollution, SDOH, Demographic, and Geographical Data: Pollution data for 9 pollution indicators along with seven social determinants of health (SDOH) / health equity census-tract level measures was extracted from the Environmental Protection Agency (EPA) Environmental Justice (EJSCREEN) 2021 database25, together with 177 chemical ambient air concentrations from the EPA's 2018 AirToxScreen database26 reported at the census block group level (in ug/m3), and calculated at the county level by population-weighting the census block group level exposures and then calculating the sum for each county from the blocks. Together, the EJSCREEN and AirToxScreen measures resulted in 186 pollution measures examined in this study. Geographical boundary information for counties, in the form of GeoJSON, were obtained from the US Census TIGER database27. The 9 EJSCREEN pollution indicators 28 included particulate matter 2.5 (PM2.5; ug/m3), ozone (parts per billion), traffic proximity (vehicles per day / meters), lead paint exposure (%; of housing units built before 1960), superfund proximity (superfund site count / km), RMP facility proximity (facility count / km), hazardous waste proximity (count of hazardous waste facilities within 5 km (or nearest beyond 5 km), each divided by distance in kilometers), underground storage tanks (count of facilities (multiplied by a factor of 7.7) within a 1,500-foot buffered block group), and wastewater discharge (modeled toxic concentrations at stream segments within 500 meters, divided by distance in kilometers (km)) (Supplementary Table 1). The year of pollution exposure was selected to precede the year when chronic disease rates were reported.

https://www.census.gov/geographies/mapping-files/time-series/geo/tiger-line-file.html

https://www.epa.gov/ejscreen

https://www.epa.gov/AirToxScreen

https://www.cdc.gov/places/index.html

## Declaration of interests

There are no conflicts of interest to declare.

### Acknowledgments

The authors would like to thank Dr. Sian Tsui (Chan School of Public Health, Harvard Medical School) and Dr. Ella Douglas-Durham (Boston Public Health Commission) for helping to review this manuscript, and the Boston Public Health Commission for providing the information technology services to support this work.

## Author contributions

Conceptualization: AD, AB, RW, KH

Methodology: AD, AB, RW, JL, KH

Investigation: AD, AB

Visualization: AD, AB

Funding acquisition: PS, BO, KH

Project administration: AD, KH

Supervision: KH

Writing – original draft: AD, RW, SN, KH

Writing – review & editing: AD, AB, RW, SN, PS, JL, BO, KH

## Data and materials availability

Sample code is available on Github (https://github.com/adeonarinebphc/apeer) and supplemental tables and figures are available in Supplemental Materials.

## References and Notes

1 Merrill PD, Ampah SB, He K, et al. Association between trace elements in the environment and stroke risk: The reasons for geographic and racial differences in stroke (REGARDS) study. J Trace Elem Med Biol 2017; 42: 45–9.

2 Howard G, Labarthe DR, Hu J, Yoon S, Howard VJ. Regional differences in African Americans’ high risk for stroke: the remarkable burden of stroke for Southern African Americans. Ann Epidemiol 2007; 17: 689–96.

3 Esenwa C, Ilunga Tshiswaka D, Gebregziabher M, Ovbiagele B. Historical Slavery and Modern-Day Stroke Mortality in the United States Stroke Belt. Stroke 2018; 49: 465–9.

4 Howard G, Howard VJ. Twenty Years of Progress Toward Understanding the Stroke Belt. Stroke 2020; 51: 742–50.

5 Lanska DJ, Kuller LH. The geography of stroke mortality in the United States and the concept of a stroke belt. Stroke 1995; 26: 1145–9.

6 Myers CA, Slack T, Broyles ST, Heymsfield SB, Church TS, Martin CK. Diabetes prevalence is associated with different community factors in the diabetes belt versus the rest of the United States. Obesity 2017; 25: 452–9.

7 Lee KK, Miller MR, Shah ASV. Air Pollution and Stroke. J Stroke Cerebrovasc Dis 2018; 20: 2–11.

8 Zanobetti A, Ryan PH, Coull BA, et al. Early-Life Exposure to Air Pollution and Childhood Asthma Cumulative Incidence in the ECHO CREW Consortium. JAMA Netw Open 2024; 7: e240535.

9 Zhang Y, Yin X, Zheng X. The relationship between PM2.5 and the onset and exacerbation of childhood asthma: a short communication. Front Pediatr 2023; 11: 1191852.

10 Valdez RB, Tabatabai M, Al-Hamdan MZ, et al. Association of diabetes and exposure to fine particulate matter (PM2.5) in the Southeastern United States. Hygiene and Environmental Health Advances 2022; 4: 100024.

11 Center for Disease Control. PLACES: Local Data for Better Health. Center for Disease Control. https://experience.arcgis.com/experience/22c7182a162d45788dd52a2362f8ed65 (accessed Sept 20, 2022).

12 Center for Disease Control. Social Determinants of Health - United States Disease Surveillance System. Center for Disease Control. https://gis.cdc.gov/grasp/diabetes/diabetesatlas-sdoh.html (accessed Sept 20, 2022).

13 Anselin L. Local Indicators of Spatial Association - LISA. Geogr Anal 1995; 27: 93–115.

14 Moran PAP. Notes on continuous stochastic phenomena. Biometrika 1950; 37: 17–23.

15 Wild CP. Complementing the genome with an ‘exposome’: the outstanding challenge of environmental exposure measurement in molecular epidemiology. Cancer Epidemiol Biomarkers Prev 2005; 14: 1847–50.

16 Smith MT, Rappaport SM. Building exposure biology centers to put the E into ‘G x E’ interaction studies. Environ Health Perspect 2009; 117: A334–5.

17 Vrijheid M. The exposome: a new paradigm to study the impact of environment on health. Thorax 2014; 69: 876–8.

18 Vermeulen R, Schymanski EL, Barabási A-L, Miller GW. The exposome and health: Where chemistry meets biology. Science 2020; 367: 392–6.

19 Kulick ER, Kaufman JD, Sack C. Ambient Air Pollution and Stroke: An Updated Review. Stroke 2023; 54: 882–93.

20 Jaccard P. THE DISTRIBUTION OF THE FLORA IN THE ALPINE ZONE.^1^. New Phytol 1912; 11: 37–50.

21 Norgeot B, Quer G, Beaulieu-Jones BK, et al. Minimum information about clinical artificial intelligence modeling: the MI-CLAIM checklist. Nat Med 2020; 26: 1320–4.

22 Greenlund KJ, Lu H, Wang Y, et al. PLACES: Local Data for Better Health. Prev Chronic Dis 2022; 19: E31.

23 US Burden of Disease Collaborators, Mokdad AH, Ballestros K, et al. The State of US Health, 1990-2016: Burden of Diseases, Injuries, and Risk Factors Among US States. JAMA 2018; 319: 1444–72.

24 Center for Disease Control. Stroke Death Rates, total population 35 and Older. Center for Disease Control. 2022; published online Sept 27. https://www.cdc.gov/dhdsp/maps/national_maps/stroke_all.htm (accessed Nov 14, 2022).

25 Owusu C, Flanagan B, Lavery AM, et al. Developing a granular scale environmental burden index (EBI) for diverse land cover types across the contiguous United States. Sci Total Environ 2022; 838: 155908.

26 Environmental Protection Agency. 2018 AirToxScreen: Assessment results. Environmental Protection Agency. 2022; published online Aug 16. https://www.epa.gov/AirToxScreen/2018-airtoxscreen-assessment-results (accessed Oct 31, 2022).

27 Her YG, Yu Z. Mapping the US Census data using the TIGER/line shapefiles. EDIS 2021; 2021. DOI:10.32473/edis-ae557-2021.

28 Environmental Protection Agency. Overview of Demographic Indicators in EJSCREEN. Environmental Protection Agency. 2023; published online July 25. https://www.epa.gov/ejscreen/overview-environmental-indicators-ejscreen (accessed June 25, 2024).

29 Longstreth WT. The REasons for Geographic And Racial Differences in Stroke (REGARDS) Study and the National Institute of Neurological Disorders and Stroke (NINDS). Stroke 2006; 37: 1147–1147.

30 Jones AC, Chaudhary NS, Patki A, et al. Neighborhood Walkability as a Predictor of Incident Hypertension in a National Cohort Study. Front Public Health 2021; 9: 611895.

31 Sîrbu CA, Stefan I, Dumitru R, et al. Air Pollution and Its Devastating Effects on the Central Nervous System. Healthcare (Basel) 2022; 10. DOI:10.3390/healthcare10071170.

32 Hantrakool S, Kumfu S, Chattipakorn SC, Chattipakorn N. Effects of Particulate Matter on Inflammation and Thrombosis: Past Evidence for Future Prevention. Int J Environ Res Public Health 2022; 19. DOI:10.3390/ijerph19148771.

33 Li X-Y, Yu X-B, Liang W-W, et al. Meta-analysis of association between particulate matter and stroke attack. CNS Neurosci Ther 2012; 18: 501–8.

34 Huang K, Liang F, Yang X, et al. Long term exposure to ambient fine particulate matter and incidence of stroke: prospective cohort study from the China-PAR project. BMJ 2019; 367: l6720.

35 Ban J, Su W, Zhong Y, Liu C, Li T. Ambient formaldehyde and mortality: A time series analysis in China. Sci Adv 2022; 8: eabm4097.

36 Wang S, Han Q, Wei Z, Wang Y, Deng L, Chen M. Formaldehyde causes an increase in blood pressure by activating ACE/AT1R axis. Toxicology 2023; 486: 153442.

37 Teichert T, Herder C. Air Pollution, Subclinical Inflammation and the Risk of Type 2 Diabetes. Environmental Influences on the Immune System. 2016; : 243–71.

38 Liu F, Chen G, Huo W, et al. Associations between long-term exposure to ambient air pollution and risk of type 2 diabetes mellitus: A systematic review and meta-analysis. Environmental Pollution. 2019; 252: 1235–45.

39 Park SK, Wang W. Ambient Air Pollution and Type 2 Diabetes Mellitus: A Systematic Review of Epidemiologic Research. Current Environmental Health Reports. 2014; 1: 275– 86.

40 Srivastava S. Effects of Environmental Polycyclic Aromatic Hydrocarbons Exposure and Pro-Inflammatory Activity on Type 2 Diabetes Mellitus in US Adults. Open Journal of Air Pollution. 2022; 11: 29–46.

41 Meo SA, Memon AN, Sheikh SA, et al. Effect of environmental air pollution on type 2 diabetes mellitus. Eur Rev Med Pharmacol Sci 2015; 19: 123–8.

42 Pye HOT, Ward-Caviness CK, Murphy BN, Appel KW, Seltzer KM. Secondary organic aerosol association with cardiorespiratory disease mortality in the United States. Nat Commun 2021; 12: 7215.

43 Ghnemat R, Alodibat S, Abu Al-Haija Q. Explainable Artificial Intelligence (XAI) for Deep Learning Based Medical Imaging Classification. J Imaging Sci Technol 2023; 9. DOI:10.3390/jimaging9090177.

44 Chen J, Hoek G. Long-term exposure to PM and all-cause and cause-specific mortality: A systematic review and meta-analysis. Environ Int 2020; 143: 105974.

45 Yuan S, Wang J, Jiang Q, et al. Long-term exposure to PM2.5 and stroke: A systematic review and meta-analysis of cohort studies. Environ Res 2019; 177: 108587.

46 Ji J, Hu L, Liu B, Li Y. Identifying and assessing the impact of key neighborhood-level determinants on geographic variation in stroke: a machine learning and multilevel modeling approach. BMC Public Health 2020; 20: 1666.

47 Chen C, Xun P, McClure LA, et al. Serum mercury concentration and the risk of ischemic stroke: The REasons for Geographic and Racial Differences in Stroke Trace Element Study. Environment International. 2018; 117: 125–31.

48 Armstrong ND, Srinivasasainagendra V, Patki A, et al. Genetic Contributors of Incident Stroke in 10,700 African Americans With Hypertension: A Meta-Analysis From the Genetics of Hypertension Associated Treatments and Reasons for Geographic and Racial Differences in Stroke Studies. Frontiers in Genetics. 2021; 12. DOI:10.3389/fgene.2021.781451.

49 Vallabhajosyula RR, Chakravarti D, Lutfeali S, Ray A, Raval A. Identifying hubs in protein interaction networks. PLoS One 2009; 4: e5344.

50 Wang H, Song M. Ckmeans.1d.dp: Optimal k-means Clustering in One Dimension by Dynamic Programming. R J 2011; 3: 29–33.

