## Supplemental Figures and Tables for "*De Novo* Exposomic Geospatial Assembly of Chronic Disease Regions with Machine Learning & Network Analysis"

**The PDF file includes:**

Materials and Methods
Figures 1-12
Tables 1-2

**Supplemental Methods**

**1. Univariate and Elastic Net Baseline Regression Models**

The relationships between 186 pollutants and 12 chronic disease measures was examined using univariate regression and multivariate elastic net regression. For each chronic disease, the county-level chronic disease rate (as a percentage of the population) was modeled as a function of independent pollution variables. First, pollution variables were normalized using the *normalize* function from the *sklearn* library (version 1.5.0), and then univariate analyses were performed using the *OLS* and *fit* functions from *statsmodels* (version 0.14.1), and the beta coefficients reported, along with associated *p*-values. To obtain the beta coefficients from the multivariate models in which all 186 pollution variables were used to predict the rates of the 12 different chronic disease indicators, the *ElasticNetCV* function from the *sklearn* library was used. First, pollution data was normalized using the *normalize* function from the *sklearn* library, and randomly split into training and test sets using a 50:50 train:test split with the *train_test_split* function in *sklearn*. A hyperparameter grid search was performed using ElasticNetCV using 3-fold cross validation, and L1 ratios of (0, 1, 0.01) and alpha values (1e-5, 1e-4, 1e-3, 1e-2, 1e-1, 0.0, 1.0, 10.0, 100.0). The β-coefficients, *p*-values, and overall $r^2$ values were then reported, and p-values were Bonferroni adjusted for multiple comparisons, with significance indicated by an asterisk. The Python code used to complete these calculations can be found in "Code and Data Availability."

**2. Calculation of Disease-Specific Correlation Matrices**

To examine the relationship between diseases and pollutants, Jaccard correlation matrices were calculated, which quantified the relationship of pairs of pollutants with specific chronic diseases. The pollution dataset consisted of 186 pollutants, and this combinatorial space was reduced using binary decomposition to 17,205 pairwise pollution combinations, which were used to construct a correlation matrix (using the Jaccard correlation coefficient) using the following steps. First, pollution measures were normalized using the *normalize* function in *sklearn*, and pollutants were divided into pairs. Then, principal component analysis (PCA) was calculated using the *PCA* function in *sklearn*, and the top two principal components were then clustered using the *KMeans* function in *sklearn*. For each pairwise combination of pollutants, 4 maps were generated corresponding to the number of clusters $k = \{2,3,4,5\}$ (resulting in 17,205 x 4 = 68,820 maps). The clustering generated sets of counties which could then be color-coded and mapped, allowing for human-interpretable output.

Once the clusterings for a pair of pollutants was calculated using PCA and k-means clustering, the correlation between the counties that are ≥ 70th percentile and the counties in each cluster were then calculated using the Jaccard correlation coefficient. Treating the counties defined by diseases and pairs of clustered pollutants as sets, the Jaccard correlation coefficient can be defined as the number of counties in the intersection of the two sets of counties divided by the union of the counties between the two sets. The Jaccard correlation coefficient was calculated for each cluster for the various clustering cutoffs $k = \{2,3,4,5\}$, and the largest Jaccard correlation was reported for a particular pair of pollutants versus a specific chronic disease (see equation 1). This process resulted in 12 chronic disease-related correlation matrices. *P*-values were then calculated for each Jaccard correlation coefficient using the Fisher exact test method, implemented in *scipy* (version ) library function *fisher_exact*. To determine which Jaccard correlation coefficients were statistically significant, the p-value threshold for significance was adjusted using the Bonferroni method, resulting in $\alpha = 0.01 / (17{,}205 \times 4) = 1.45 \times 10^{-7}$.

**3. Chronic Disease-Related Pollution Network Construction**

The correlation matrices produced several statistically significant disease-pollution pair relationships quantified by high Jaccard values. A systematic way of identifying which pollutants is important from the statistically significant relationships in the correlation matrices for each disease is to construct a chronic disease-related pollution network from each correlation matrix, and identify the pollution hubs. To construct the networks, pollutants in the correlation matrix were nodes, and the edges between each pollutant was a statistically significant (with Bonferroni adjustment) Jaccard correlation coefficient ≥ 84th percentile (half a standard deviation above the mean). The networkx library (version 3.2.1) was used to calculate the networks, and node degree (the number of connections for each node) was calculated. Typically, there is no well established method to determine whether a node was a hub by degree, with arbitrary cutoffs usually used [49]. To determine which node was a hub based on degree, the *ckmeans* function in the *ckmeans_1d_dp* library (version 4.3.4.4 [50]) was used to cluster nodes based on degree (with $k$=2), and nodes (pollutants) that fell into the cluster with the higher average degrees were designated as hubs.

**4. Elastic Net Regression**

Elastic net regression models were then developed in which the dependent variable consisted of whether a county was ≥ 70th percentile for a disease (assigned a value of 1) or below (assigned a value of 0). County-level pollution data, comprising the independent variables in this model, were normalized using the *normalize* function from the *sklearn* library, and randomly split into training and test sets using a 50:50 train:test split with the *train_test_split* function in *sklearn*. Elastic net regression using three-fold validation was performed using the *LogisticRegressionCV* function from the *sklearn* library (with the stochastic average gradient SAGA solver) which performed a hyperparameter grid search with the following parameters: regularization strength Cs = (0.001, 0.01, 0.05, 0.1, 1.0, 100.0), “elasticnet” penalty, “roc_auc” scoring, and L1 ratios consisting of (0.15, 0.25, 0.5, 0.75), and a random state of 0. The Python code for this regression model can be found in “Code and Data Availability.”

**5. Random Forest Regression**

To create random-forest regression models, county-level data was randomly divided into training and test sets using a 50:50 ratio with the *train_test_split* function in *sklearn*, with the dependent and independent variables defined in the same fashion as the elastic net models developed with *LogisticRegressionCV* above. Random forest regression models were calculated using the *xgboost* (version 1.6.2) library. A hyperparameter grid search was performed with minimum child weight (1,5, 10), gamma (0.5, 1, 1.5, 2, 5), subsample (0.6, 0.8, 1.0), percentage of features (colsample_bytree) (0.6, 0.8, 1.0) and maximum tree depth (3, 4, 5). The Python code for creating the xgboost-based random forest models can be found in “Code and Data Availability.”

### 6. LISA and Moran's *I* Calculations

The rates of chronic disease indicators (at the county level) along with pollutants were analyzed for spatial autocorrelation by calculating Moran's *I* and LISA. Moran's I is a method for calculating spatial autocorrelation (a phenomenon in which a particular measure is correlated with the same measure in other close by geographical regions) and is a generally used method for examining geospatial patterns in data [14]. LISA, which stands for local indicators of spatial association, involves calculating a local Moran's *I* (ie. for a specific geographical region (like a county or set of counties) rather than across the entire set of counties) and then plotting the statistically significant values, which can be used to identify geospatial clusters [13]. To calculate Moran's *I* the *splot* library was used (version 1.1.5), and spatial weights were calculated using the *lag_spatial* function, and then the *Moran* function (for the global I) and *moran_local* function (for the local Moran's *I*, used to construct LISA maps). LISA maps were then generated using the *lisa_cluster* function in *splot*. We calculated global Moran's *I* for each pollutant and assessed the statistical significance of these values using a Manhattan plot (using a Bonferroni-adjusted threshold for significance) which was plotted using the *plot* function from *matplotlib* (version 3.8.2).

### Figure and Table Captions

Figure 1. The 6-Step aPEER workflow. Step 1: Generate reference maps of chronic disease prevalence and stroke mortality (≥70th percentile). Step 2: Clusters derived from principal component analysis (PCA) and k-means clustering of 186 pollutants projected on a US map. Step 3: Compare disease and pollution maps using Jaccard correlation coefficient (J). Step 4: Network analysis used to prioritize strongest relationships and identify key disease-related pollutants. Step 5: Findings benchmarked by examining how closely geographical distribution of key pollutants resembles disease maps. Prediction of disease prevalence by pollutants compared to known predictors like risk factors and SDOH. Step 6: Examine relationships among disease-pollution hubs using hierarchical clustering analysis.

Table 1. County-level descriptive statistics for chronic disease and healthcare, pollution indicators and demographic data for 3141 counties used in this study (N = number of counties equal to or above a percentile cutoff; * = chronic diseases/indicators that are modeled in this study).

Figure 2. The Jaccard Indices ($p \ll$ Bonferroni-adjusted threshold 0.001) between the top 10 pollutants (ranked by Jaccard J) for (A) hypertension and (B) stroke mortality (map colors are arbitrary).

Figure 3. Pollution networks for (A) hypertension and (B) stroke mortality, with elastic net and random forest models predicting the geographical distribution of a given disease using pollution hubs compared to SDOH, prevention, and pollution (all pollution features) models.

Figure 4. A comparison of the pollutants identified as being highly predictive of hypertension and stroke mortality using aPEER (pollution hubs), elastic net (β coefficients), and random forest models (importance). Three pollutants (formaldehyde, methanol, and acetaldehyde) consistently appeared irrespective of the analysis method employed (highlighted in red).

Figure 5. (A) The top 5 disease-pollution associations derived from assembled pollution maps (70th percentile) ranked by Jaccard correlation coefficients, and (B) clustered heatmap of aPEER pollution hubs, elastic net β coefficients, and random forest importance features showing methanol, formaldehyde, and acetaldehyde closely clustered together, and strongly associated with multiple cardiometabolic diseases (highlighted in red).

Supplementary Table 1. Data sources for disease, healthcare, demographic and pollution measures and the models into which they were incorporated (the “Model” column). Data sources included CDC places 2021, EPA EJSCREEN, and the Air Tox Screen from 2018. Note the pollution data was collected at an earlier time point to the healthcare and disease prevalence data.

Supplementary Table 2. Univariate and multivariate regression showing the top β coefficient values predicting each disease / health indicator (if more than 20 were present, only the top 20 were presented). * = statistically significant with Bonferroni-adjustment (padj).

Supplementary Figure 1. Chronic disease reference maps where counties with rates ≥ 70th percentile are highlighted in blue.

Supplementary Figure 2. Pairwise pollution correlation matrices for the top 10 pollutants (by Jaccard correlation coefficient) associated with the chronic diseases. For each pairwise combination of pollutants, a map was calculated using aPEER, and the Jaccard index was calculated relative to a chronic disease (asthma, arthritis, etc). The map with the highest Jaccard index out of the possible maps out of clustering values k={2, 3, 4, 5} was then identified for each pairwise combination.

Supplementary Figure 3. aPEER pollution networks constructed from Jaccard correlation coefficients for 12 health-related indicators.

Supplementary Figure 4. Receiver-operator (ROC) curves and area under the curve (AUC) values for elastic net and random forest (XGBoost) models predicting whether a county was within (1) or outside (0) a given disease map, where independent variables consisted of (1) preventive healthcare measures, (2) SDOH measures, (3) all pollutants in this study, or (4) just the hub pollutants from aPEER network analysis for a respective disease.

Supplementary Figure 5A. Calibration curves for XGBoost random forest models.

Supplementary Figure 5B. Calibration curves for elastic net regression models.

Supplementary Figure 6. Comparison of aPEER pollution hubs (left), pollution-associated Elastic Net β coefficients (middle), and random forest-associated pollution feature importance (right) for each disease.

Supplementary Figure 7. The strongest disease-pollution associations, ranked by Jaccard correlation coefficient (map assembly derived from the top 2 hub pollutants identified by pairwise Jaccard correlation coefficient).

Supplementary Figure 8. Sensitivity analysis of disease-pollution associations. Clustering the results from aPEER, elastic net, and random forest-derived pollution data at different disease thresholds showed consistent patterns for aPEER and elastic net.

Supplementary Figure 9. Spatial analysis benchmarks using Moran's I (scatterplots) and LISA (county-level maps of the US). Calculation of Moran's I and LISA maps for stroke mortality rate and different pollution indicators. Note that none of the Moran's I's or LISA maps were statistically significant ($p >> 0.1$), and no discernable patterns appeared even with a significantly relaxed p-value threshold ($p < 0.1$).

Supplementary Figure 10. Calculation of Moran's I for 186 pollutants. Manhattan plot of the p-values associated with Moran's I and the Bonferroni-adjusted threshold of statistical significance (red). No pollutants were found to exhibit statistically-significant geospatial clustering.

Supplementary Figure 11. County-level pollution/population correlation. The correlation between stroke mortality and population at the county level (natural-log transformed).

Supplementary Figure 12. County-level pollution density plots. Density plots with normalized data for selected pollutants and diseases.

| Data Source | Measure | Model | Description |
|---|---|---|---|
| CDC PLACES 2021 | ACCESS2_CrudePrev | | Current lack of health insurance among adults aged 18–64 years |
| CDC PLACES 2021 | ARTHRITIS_CrudePrev | | Arthritis among adults aged ≥18 years |
| CDC PLACES 2021 | BINGE_CrudePrev | | Binge drinking among adults aged ≥18 years |
| CDC PLACES 2021 | BPHIGH_CrudePrev | Chronic Disease | High blood pressure among adults aged ≥18 years |
| CDC PLACES 2021 | BPMED_CrudePrev | | Taking medicine for high blood pressure control among adults aged ≥18 years with high blood pressure |
| CDC PLACES 2021 | CANCER_CrudePrev | Chronic Disease | Cancer among adults aged ≥18 years |
| CDC PLACES 2021 | CASTHMA_CrudePrev | Chronic Disease | Current asthma prevalence among adults aged ≥18 years |
| CDC PLACES 2021 | CERVICAL_CrudePrev | | Cervical cancer screening among adult women aged 21–65 years |
| CDC PLACES 2021 | CHD_CrudePrev | Chronic Disease | Coronary heart disease among adults aged ≥18 years |
| CDC PLACES 2021 | CHECKUP_CrudePrev | | Visits to doctor for routine checkup within the past year among adults aged ≥18 years |
| CDC PLACES 2021 | CHOLSCREEN_CrudePrev | | Cholesterol screening among adults aged ≥18 years |
| CDC PLACES 2021 | COLON_SCREEN_CrudePrev | | Fecal occult blood test, sigmoidoscopy, or colonoscopy among adults aged 50–75 years |
| CDC PLACES 2021 | COPD_CrudePrev | Chronic Disease | Chronic obstructive pulmonary disease among adults aged ≥18 years |
| CDC PLACES 2021 | COREM_CrudePrev | | Older adults aged ≥65 years who are up to date on a core set of clinical preventive services by age and sex |
| CDC PLACES 2021 | COREW_CrudePrev | | Older adults aged ≥65 years who are up to date on a core set of clinical preventive services by age and sex |
| CDC PLACES 2021 | CSMOKING_CrudePrev | Chronic Disease | Current smoking among adults aged ≥18 years |
| CDC PLACES 2021 | DENTAL_CrudePrev | | Visits to dentist or dental clinic among adults aged ≥18 years |
| CDC PLACES 2021 | DEPRESSION_CrudePrev | | Depression among adults aged ≥18 years |
| CDC PLACES 2021 | DIABETES_CrudePrev | Chronic Disease | Diagnosed diabetes among adults aged ≥18 years |
| CDC PLACES 2021 | GHLTH_CrudePrev | | Fair or poor self-rated health status among adults aged ≥18 years |
| CDC PLACES 2021 | HIGHCHOL_CrudePrev | Chronic Disease | High cholesterol among adults aged ≥18 years who have been screened in the past 5 years |
| CDC PLACES 2021 | KIDNEY_CrudePrev | Chronic Disease | Chronic kidney disease among adults aged ≥18 years |
| CDC PLACES 2021 | LPA_CrudePrev | | No leisure-time physical activity among adults aged ≥18 years |
| CDC PLACES 2021 | MAMMOUSE_CrudePrev | | Mammography use among women aged 50-74 years |
| CDC PLACES 2021 | MHLTH_CrudePrev | | Mental health not good for ≥14 days among adults aged ≥18 years |
| CDC PLACES 2021 | OBESITY_CrudePrev | Chronic Disease | Obesity among adults aged ≥18 years |
| CDC PLACES 2021 | PHLTH_CrudePrev | | Physical health not good for ≥14 days among adults aged ≥18 years |
| CDC PLACES 2021 | SLEEP_CrudePrev | | Sleeping less than 7 hours among adults aged ≥18 years |
| CDC PLACES 2021 | STROKE_CrudePrev | Chronic Disease | Stroke among adults aged ≥18 years |
| CDC PLACES 2021 | TEETHLOST_CrudePrev | | All teeth lost among adults aged ≥65 years |
| CDC Stroke Mortality | Stroke Mortality | Chronic Disease | Stroke Mortality in Adults ≥ 35 years |
| EPA EJSCREEN | ozone | Air Pollution, Pollution | Ozone |
| EPA EJSCREEN | ptraf | Pollution | Traffic Proximity |
| EPA EJSCREEN | pwdis | Pollution | Wastewater discharge |
| EPA EJSCREEN | pnpl | Pollution | Superfund Proximity |
| EPA EJSCREEN | prmp | Pollution | RMP Facility Proximity |
| EPA EJSCREEN | ptsdf | Pollution | Hazardous Waste Proximity |
| EPA EJSCREEN | ust | Pollution | Underground Storage Tanks |
| EPA EJSCREEN | pre1960pct | Pollution | Lead Paint |
| EPA EJSCREEN | MINORPCT | Demographic | Percent Minority |
| EPA EJSCREEN | LOWINCPCT | Demographic | Percent Low Income |
| EPA EJSCREEN | LESSHSPCT | Demographic | Percent of Adults with Less than High School Education |
| EPA EJSCREEN | LINGISOPCT | Demographic | Percent Linguistically Isolated |
| EPA EJSCREEN | OVER64PCT | Demographic | Percent Over Age 64 |
| EPA EJSCREEN | UNDER5PCT | Demographic | Percent Under Age 5 |
| EPA EJSCREEN | UNEMPPCT | Demographic | Percent Unemployment |
| AirToxScreen 2018 | 1-BROMOPROPANE | Exposome | Ambient Chemical Air Concentration |
| AirToxScreen 2018 | 1,1-DIMETHYLHYDRAZINE | Exposome | Ambient Chemical Air Concentration |
| AirToxScreen 2018 | 1,1,2-TRICHLOROETHANE | Exposome | Ambient Chemical Air Concentration |
| AirToxScreen 2018 | 1,1,2,2-TETRACHLOROETHANE | Exposome | Ambient Chemical Air Concentration |
| AirToxScreen 2018 | 1,2-DIBROMO-3-CHLOROPROPANE | Exposome | Ambient Chemical Air Concentration |
| AirToxScreen 2018 | 1,2-DIPHENYLHYDRAZINE | Exposome | Ambient Chemical Air Concentration |
| AirToxScreen 2018 | 1,2-EPOXYBUTANE | Exposome | Ambient Chemical Air Concentration |
| AirToxScreen 2018 | 1,2-PROPYLENEIMINE | Exposome | Ambient Chemical Air Concentration |
| AirToxScreen 2018 | 1,2,3,4,5,6-HEXACHLOROCYCLYHEXANE | Exposome | Ambient Chemical Air Concentration |
| AirToxScreen 2018 | 1,2,4-TRICHLOROBENZENE | Exposome | Ambient Chemical Air Concentration |
| AirToxScreen 2018 | 1,3-BUTADIENE | Exposome | Ambient Chemical Air Concentration |
| AirToxScreen 2018 | 1,3-DICHLOROPROPENE | Exposome | Ambient Chemical Air Concentration |
| AirToxScreen 2018 | 1,3-PROPANE SULTONE | Exposome | Ambient Chemical Air Concentration |
| AirToxScreen 2018 | 1,4-DICHLOROBENZENE | Exposome | Ambient Chemical Air Concentration |
| AirToxScreen 2018 | 2-ACETYLAMINOFLUORENE | Exposome | Ambient Chemical Air Concentration |
| AirToxScreen 2018 | 2- | Exposome | Ambient Chemical Air Concentration |

| | | | |
|---|---|---|---|
| | CHLOROACETOPHENONE | | |
| AirToxScreen 2018 | 2-NITROPROPANE | Exposome | Ambient Chemical Air Concentration |
| AirToxScreen 2018 | 2,2,4-TRIMETHYLPENTANE | Exposome | Ambient Chemical Air Concentration |
| AirToxScreen 2018 | 2,4-D, SALTS AND ESTERS | Exposome | Ambient Chemical Air Concentration |
| AirToxScreen 2018 | 2,4-DINITROPHENOL | Exposome | Ambient Chemical Air Concentration |
| AirToxScreen 2018 | 2,4-DINITROTOLUENE | Exposome | Ambient Chemical Air Concentration |
| AirToxScreen 2018 | 2,4-TOLUENE DIISOCYANATE | Exposome | Ambient Chemical Air Concentration |
| AirToxScreen 2018 | 2,4,5-TRICHLOROPHENOL | Exposome | Ambient Chemical Air Concentration |
| AirToxScreen 2018 | 2,4,6-TRICHLOROPHENOL | Exposome | Ambient Chemical Air Concentration |
| AirToxScreen 2018 | 3,3'-DICHLOROBENZIDINE | Exposome | Ambient Chemical Air Concentration |
| AirToxScreen 2018 | 3,3'-DIMETHOXYBENZIDINE | Exposome | Ambient Chemical Air Concentration |
| AirToxScreen 2018 | 3,3'-DIMETHYLBENZIDINE | Exposome | Ambient Chemical Air Concentration |
| AirToxScreen 2018 | 4-AMINOBIPHENYL | Exposome | Ambient Chemical Air Concentration |
| AirToxScreen 2018 | 4-DIMETHYLAMINOAZOBENZENE | Exposome | Ambient Chemical Air Concentration |
| AirToxScreen 2018 | 4-NITROBIPHENYL | Exposome | Ambient Chemical Air Concentration |
| AirToxScreen 2018 | 4-NITROPHENOL | Exposome | Ambient Chemical Air Concentration |
| AirToxScreen 2018 | 4,4'-METHYLENE BIS(2-CHLOROANILINE) | Exposome | Ambient Chemical Air Concentration |
| AirToxScreen 2018 | 4,4'-METHYLENEDIANILINE | Exposome | Ambient Chemical Air Concentration |
| AirToxScreen 2018 | 4,4'-METHYLENEDIPHENYL DIISOCYANATE (MDI) | Exposome | Ambient Chemical Air Concentration |
| AirToxScreen 2018 | 4,6-DINITRO-O-CRESOL (INCLUDING SALTS) | Exposome | Ambient Chemical Air Concentration |
| AirToxScreen 2018 | ACETALDEHYDE | Exposome | Ambient Chemical Air Concentration |
| AirToxScreen 2018 | ACETAMIDE | Exposome | Ambient Chemical Air Concentration |
| AirToxScreen 2018 | ACETONITRILE | Exposome | Ambient Chemical Air Concentration |
| AirToxScreen 2018 | ACETOPHENONE | Exposome | Ambient Chemical Air Concentration |
| AirToxScreen 2018 | ACROLEIN | Exposome | Ambient Chemical Air Concentration |
| AirToxScreen 2018 | ACRYLAMIDE | Exposome | Ambient Chemical Air Concentration |
| AirToxScreen 2018 | ACRYLIC ACID | Exposome | Ambient Chemical Air Concentration |
| AirToxScreen 2018 | ACRYLONITRILE | Exposome | Ambient Chemical Air Concentration |
| AirToxScreen 2018 | ALLYL CHLORIDE | Exposome | Ambient Chemical Air Concentration |
| AirToxScreen 2018 | ANILINE | Exposome | Ambient Chemical Air Concentration |
| AirToxScreen 2018 | ANTIMONY COMPOUNDS | Exposome | Ambient Chemical Air Concentration |
| AirToxScreen 2018 | ARSENIC COMPOUNDS(INORGANIC INCLUDING ARSINE) | Exposome | Ambient Chemical Air Concentration |
| AirToxScreen 2018 | BENZENE | Exposome | Ambient Chemical Air Concentration |
| AirToxScreen 2018 | BENZIDINE | Exposome | Ambient Chemical Air Concentration |
| AirToxScreen 2018 | BENZOAPYRENE | Exposome | Ambient Chemical Air Concentration |
| AirToxScreen 2018 | BENZOTRICHLORIDE | Exposome | Ambient Chemical Air Concentration |
| AirToxScreen 2018 | BENZYL CHLORIDE | Exposome | Ambient Chemical Air Concentration |
| AirToxScreen 2018 | BERYLLIUM COMPOUNDS | Exposome | Ambient Chemical Air Concentration |
| AirToxScreen 2018 | BIPHENYL | Exposome | Ambient Chemical Air Concentration |
| AirToxScreen 2018 | BIS(2-ETHYLHEXYL)PHTHALATE (DEHP) | Exposome | Ambient Chemical Air Concentration |
| AirToxScreen 2018 | BIS(CHLOROMETHYL) ETHER | Exposome | Ambient Chemical Air Concentration |
| AirToxScreen 2018 | BROMOFORM | Exposome | Ambient Chemical Air Concentration |
| AirToxScreen 2018 | CADMIUM COMPOUNDS | Exposome | Ambient Chemical Air Concentration |
| AirToxScreen 2018 | CALCIUM CYANAMIDE | Exposome | Ambient Chemical Air Concentration |
| AirToxScreen 2018 | CAPTAN | Exposome | Ambient Chemical Air Concentration |
| AirToxScreen 2018 | CARBARYL | Exposome | Ambient Chemical Air Concentration |
| AirToxScreen 2018 | CARBON DISULFIDE | Exposome | Ambient Chemical Air Concentration |
| AirToxScreen 2018 | CARBON TETRACHLORIDE | Exposome | Ambient Chemical Air Concentration |
| AirToxScreen 2018 | CARBONYL SULFIDE | Exposome | Ambient Chemical Air Concentration |
| AirToxScreen 2018 | CATECHOL | Exposome | Ambient Chemical Air Concentration |
| AirToxScreen 2018 | Chloramben | Exposome | Ambient Chemical Air Concentration |
| AirToxScreen 2018 | CHLORDANE | Exposome | Ambient Chemical Air Concentration |
| AirToxScreen 2018 | CHLORINE | Exposome | Ambient Chemical Air Concentration |
| AirToxScreen 2018 | CHLOROACETIC ACID | Exposome | Ambient Chemical Air Concentration |
| AirToxScreen 2018 | CHLOROBENZENE | Exposome | Ambient Chemical Air Concentration |
| AirToxScreen 2018 | CHLOROBENZILATE | Exposome | Ambient Chemical Air Concentration |
| AirToxScreen 2018 | CHLOROFORM | Exposome | Ambient Chemical Air Concentration |

| | | | |
|---|---|---|---|
| AirToxScreen 2018 | CHLOROMETHYL METHYL ETHER | Exposome | Ambient Chemical Air Concentration |
| AirToxScreen 2018 | CHLOROPRENE | Exposome | Ambient Chemical Air Concentration |
| AirToxScreen 2018 | CHROMIUM VI (HEXAVALENT) | Exposome | Ambient Chemical Air Concentration |
| AirToxScreen 2018 | COBALT COMPOUNDS | Exposome | Ambient Chemical Air Concentration |
| AirToxScreen 2018 | COKE OVEN EMISSIONS | Exposome | Ambient Chemical Air Concentration |
| AirToxScreen 2018 | CRESOL_CRESYLIC ACID (MIXED ISOMERS) | Exposome | Ambient Chemical Air Concentration |
| AirToxScreen 2018 | CUMENE | Exposome | Ambient Chemical Air Concentration |
| AirToxScreen 2018 | CYANIDE COMPOUNDS | Exposome | Ambient Chemical Air Concentration |
| AirToxScreen 2018 | DIBENZOFURAN | Exposome | Ambient Chemical Air Concentration |
| AirToxScreen 2018 | DIBUTYLPHTHALATE | Exposome | Ambient Chemical Air Concentration |
| AirToxScreen 2018 | DICHLOROETHYL ETHER (BIS[2-CHLOROETHYL]ETHER) | Exposome | Ambient Chemical Air Concentration |
| AirToxScreen 2018 | DICHLORVOS | Exposome | Ambient Chemical Air Concentration |
| AirToxScreen 2018 | DIESEL PM | Exposome | Ambient Chemical Air Concentration |
| AirToxScreen 2018 | DIETHANOLAMINE | Exposome | Ambient Chemical Air Concentration |
| AirToxScreen 2018 | DIETHYL SULFATE | Exposome | Ambient Chemical Air Concentration |
| AirToxScreen 2018 | DIMETHYL PHTHALATE | Exposome | Ambient Chemical Air Concentration |
| AirToxScreen 2018 | DIMETHYL SULFATE | Exposome | Ambient Chemical Air Concentration |
| AirToxScreen 2018 | DIMETHYLCARBAMOYL CHLORIDE | Exposome | Ambient Chemical Air Concentration |
| AirToxScreen 2018 | EPICHLOROHYDRIN | Exposome | Ambient Chemical Air Concentration |
| AirToxScreen 2018 | ETHYL ACRYLATE | Exposome | Ambient Chemical Air Concentration |
| AirToxScreen 2018 | ETHYLBENZENE | Exposome | Ambient Chemical Air Concentration |
| AirToxScreen 2018 | ETHYL CARBAMATE (URETHANE) CHLORIDE (CHLOROETHANE) | Exposome | Ambient Chemical Air Concentration |
| AirToxScreen 2018 | ETHYL CHLORIDE | Exposome | Ambient Chemical Air Concentration |
| AirToxScreen 2018 | ETHYLENE DIBROMIDE (DIBROMOETHANE) | Exposome | Ambient Chemical Air Concentration |
| AirToxScreen 2018 | ETHYLENE DICHLORIDE (1,2-DICHLOROETHANE) | Exposome | Ambient Chemical Air Concentration |
| AirToxScreen 2018 | ETHYLENE GLYCOL | Exposome | Ambient Chemical Air Concentration |
| AirToxScreen 2018 | ETHYLENE OXIDE | Exposome | Ambient Chemical Air Concentration |
| AirToxScreen 2018 | ETHYLENE THIOUREA | Exposome | Ambient Chemical Air Concentration |
| AirToxScreen 2018 | ETHYLENEIMINE (AZIRIDINE) | Exposome | Ambient Chemical Air Concentration |
| AirToxScreen 2018 | ETHYLIDENE DICHLORIDE (1,1-DICHLOROETHANE) | Exposome | Ambient Chemical Air Concentration |
| AirToxScreen 2018 | FORMALDEHYDE | Exposome | Ambient Chemical Air Concentration |
| AirToxScreen 2018 | GLYCOL ETHERS | Exposome | Ambient Chemical Air Concentration |
| AirToxScreen 2018 | HEPTACHLOR | Exposome | Ambient Chemical Air Concentration |
| AirToxScreen 2018 | HEXACHLOROBENZENE | Exposome | Ambient Chemical Air Concentration |
| AirToxScreen 2018 | HEXACHLOROBUTADIENE | Exposome | Ambient Chemical Air Concentration |
| AirToxScreen 2018 | HEXACHLOROCYCLOPENTADIENE | Exposome | Ambient Chemical Air Concentration |
| AirToxScreen 2018 | HEXACHLOROETHANE | Exposome | Ambient Chemical Air Concentration |
| AirToxScreen 2018 | HEXAMETHYLENE DIISOCYANATE | Exposome | Ambient Chemical Air Concentration |
| AirToxScreen 2018 | HEXAMETHYLPHOSPHORAMIDE | Exposome | Ambient Chemical Air Concentration |
| AirToxScreen 2018 | HEXANE | Exposome | Ambient Chemical Air Concentration |
| AirToxScreen 2018 | HYDRAZINE | Exposome | Ambient Chemical Air Concentration |
| AirToxScreen 2018 | HYDROCHLORIC ACID (HYDROGEN CHLORIDE [GAS ONLY]) | Exposome | Ambient Chemical Air Concentration |
| AirToxScreen 2018 | HYDROGEN FLUORIDE (HYDROFLUORIC ACID) | Exposome | Ambient Chemical Air Concentration |
| AirToxScreen 2018 | HYDROQUINONE | Exposome | Ambient Chemical Air Concentration |
| AirToxScreen 2018 | ISOPHORONE | Exposome | Ambient Chemical Air Concentration |
| AirToxScreen 2018 | LEAD COMPOUNDS | Exposome | Ambient Chemical Air Concentration |
| AirToxScreen 2018 | MALEIC ANHYDRIDE | Exposome | Ambient Chemical Air Concentration |
| AirToxScreen 2018 | MANGANESE COMPOUNDS | Exposome | Ambient Chemical Air Concentration |
| AirToxScreen 2018 | MERCURY COMPOUNDS | Exposome | Ambient Chemical Air Concentration |
| AirToxScreen 2018 | METHANOL | Exposome | Ambient Chemical Air Concentration |
| AirToxScreen 2018 | METHOXYCHLOR | Exposome | Ambient Chemical Air Concentration |
| AirToxScreen 2018 | METHYL BROMIDE (BROMOMETHANE) | Exposome | Ambient Chemical Air Concentration |

| | | | |
|---|---|---|---|
| AirToxScreen 2018 | METHYL CHLORIDE (CHLOROMETHANE) | Exposome | Ambient Chemical Air Concentration |
| AirToxScreen 2018 | 1,1,1-TRICHLOROETHANE | Exposome | Ambient Chemical Air Concentration |
| AirToxScreen 2018 | METHYL IODIDE (IODOMETHANE) | Exposome | Ambient Chemical Air Concentration |
| AirToxScreen 2018 | METHYL ISOBUTYL KETONE (HEXONE) | Exposome | Ambient Chemical Air Concentration |
| AirToxScreen 2018 | METHYL ISOCYANATE | Exposome | Ambient Chemical Air Concentration |
| AirToxScreen 2018 | METHYL METHACRYLATE | Exposome | Ambient Chemical Air Concentration |
| AirToxScreen 2018 | METHYL TERT-BUTYL ETHER | Exposome | Ambient Chemical Air Concentration |
| AirToxScreen 2018 | METHYLENE CHLORIDE | Exposome | Ambient Chemical Air Concentration |
| AirToxScreen 2018 | METHYLHYDRAZINE | Exposome | Ambient Chemical Air Concentration |
| AirToxScreen 2018 | N-Nitroso-N-Methylurea | Exposome | Ambient Chemical Air Concentration |
| AirToxScreen 2018 | N-NITROSODIMETHYLAMINE | Exposome | Ambient Chemical Air Concentration |
| AirToxScreen 2018 | N-NITROSOMORPHOLINE | Exposome | Ambient Chemical Air Concentration |
| AirToxScreen 2018 | N,N-DIMETHYLANILINE | Exposome | Ambient Chemical Air Concentration |
| AirToxScreen 2018 | DIMETHYL FORMAMIDE | Exposome | Ambient Chemical Air Concentration |
| AirToxScreen 2018 | NAPHTHALENE | Exposome | Ambient Chemical Air Concentration |
| AirToxScreen 2018 | NICKEL COMPOUNDS | Exposome | Ambient Chemical Air Concentration |
| AirToxScreen 2018 | NITROBENZENE | Exposome | Ambient Chemical Air Concentration |
| AirToxScreen 2018 | ANISIDINE | Exposome | Ambient Chemical Air Concentration |
| AirToxScreen 2018 | O-TOLUIDINE | Exposome | Ambient Chemical Air Concentration |
| AirToxScreen 2018 | 1,4-DIOXANE | Exposome | Ambient Chemical Air Concentration |
| AirToxScreen 2018 | P-PHENYLENEDIAMINE | Exposome | Ambient Chemical Air Concentration |
| AirToxScreen 2018 | PAHPOM | Exposome | Ambient Chemical Air Concentration |
| AirToxScreen 2018 | Parathion | Exposome | Ambient Chemical Air Concentration |
| AirToxScreen 2018 | POLYCHLORINATED BIPHENYLS (AROCLORS) | Exposome | Ambient Chemical Air Concentration |
| AirToxScreen 2018 | PENTACHLORONITROBENZENE (QUINTOBENZENE) | Exposome | Ambient Chemical Air Concentration |
| AirToxScreen 2018 | PENTACHLOROPHENOL | Exposome | Ambient Chemical Air Concentration |
| AirToxScreen 2018 | PHENOL | Exposome | Ambient Chemical Air Concentration |
| AirToxScreen 2018 | PHOSGENE | Exposome | Ambient Chemical Air Concentration |
| AirToxScreen 2018 | PHOSPHINE | Exposome | Ambient Chemical Air Concentration |
| AirToxScreen 2018 | PHOSPHORUS | Exposome | Ambient Chemical Air Concentration |
| AirToxScreen 2018 | PHTHALIC ANHYDRIDE | Exposome | Ambient Chemical Air Concentration |
| AirToxScreen 2018 | PROPIONALDEHYDE | Exposome | Ambient Chemical Air Concentration |
| AirToxScreen 2018 | PROPOXUR (BAYGON) | Exposome | Ambient Chemical Air Concentration |
| AirToxScreen 2018 | PROPYLENE DICHLORIDE (1,2-DICHLOROPROPANE) | Exposome | Ambient Chemical Air Concentration |
| AirToxScreen 2018 | PROPYLENE OXIDE | Exposome | Ambient Chemical Air Concentration |
| AirToxScreen 2018 | QUINOLINE | Exposome | Ambient Chemical Air Concentration |
| AirToxScreen 2018 | QUINONE (P-BENZOQUINONE) | Exposome | Ambient Chemical Air Concentration |
| AirToxScreen 2018 | SELENIUM COMPOUNDS | Exposome | Ambient Chemical Air Concentration |
| AirToxScreen 2018 | STYRENE | Exposome | Ambient Chemical Air Concentration |
| AirToxScreen 2018 | STYRENE OXIDE | Exposome | Ambient Chemical Air Concentration |
| AirToxScreen 2018 | TETRACHLOROETHYLENE | Exposome | Ambient Chemical Air Concentration |
| AirToxScreen 2018 | TITANIUM TETRACHLORIDE | Exposome | Ambient Chemical Air Concentration |
| AirToxScreen 2018 | 2,4-TOLUENE DIAMINE | Exposome | Ambient Chemical Air Concentration |
| AirToxScreen 2018 | TOLUENE | Exposome | Ambient Chemical Air Concentration |
| AirToxScreen 2018 | TOXAPHENE (CHLORINATED CAMPHENE) | Exposome | Ambient Chemical Air Concentration |
| AirToxScreen 2018 | TRICHLOROETHYLENE | Exposome | Ambient Chemical Air Concentration |
| AirToxScreen 2018 | TRIETHYLAMINE | Exposome | Ambient Chemical Air Concentration |
| AirToxScreen 2018 | TRIFLURALIN | Exposome | Ambient Chemical Air Concentration |
| AirToxScreen 2018 | VINYL ACETATE | Exposome | Ambient Chemical Air Concentration |
| AirToxScreen 2018 | VINYL BROMIDE | Exposome | Ambient Chemical Air Concentration |
| AirToxScreen 2018 | VINYL CHLORIDE | Exposome | Ambient Chemical Air Concentration |
| AirToxScreen 2018 | VINYLIDENE CHLORIDE | Exposome | Ambient Chemical Air Concentration |
| AirToxScreen 2018 | XYLENES (MIXED ISOMERS) | Exposome | Ambient Chemical Air Concentration |

**Supplementary Table 1.** Data sources for disease, healthcare, demographic and pollution measures and the models into which they were incorporated (the “Model” column). Data sources included CDC places 2021, EPA EJSCREEN, and the Air Tox Screen from 2018. Note the pollution data was collected at an earlier time point to the healthcare and disease prevalence data.

1. Arthritis

| Variable | Univariate β | *p* | $p_{adj}$ | $r^2$ | Multivariate β | *p* | $p_{adj}$ | $r^2$ |
|---|---|---|---|---|---|---|---|---|
| Carbon Tetrachloride | 535.63 | 3.96E-9 | * | 0.01 | 724.12 | 4.59E-7 | * | 0.47 |
| Acetaldehyde | 183.83 | 5.96E-50 | * | 0.07 | 398.86 | 1.21E-17 | * | 0.47 |
| Lead Paint | 9.72 | 3.40E-1 | | -0.00 | 144.35 | 2.51E-37 | * | 0.47 |
| 4-Nitrophenol | -85.12 | 4.50E-67 | * | 0.09 | 80.79 | 5.27E-12 | * | 0.47 |
| Xylenes (Mixed Isomers) | -86.25 | 7.10E-46 | * | 0.06 | 73.74 | 2.28E-8 | * | 0.47 |
| 2,2,4-Trimethylpentane | -106.97 | 7.21E-67 | * | 0.09 | 57.01 | 6.37E-6 | * | 0.47 |
| Cyanide Compounds | -33.64 | 6.98E-11 | * | 0.01 | 29.01 | 3.84E-6 | * | 0.47 |
| 2-Chloroacetophenone | -7.29 | 1.26E-1 | | 0.00 | 22.30 | 1.46E-5 | * | 0.47 |
| Wastewater Discharge | 19.91 | 2.17E-5 | * | 0.01 | 15.87 | 5.58E-6 | * | 0.47 |
| Traffic Proximity | -59.90 | 2.97E-36 | * | 0.05 | -20.66 | 1.14E-5 | * | 0.47 |
| Underground Storage Tanks | -108.16 | 1.01E-80 | * | 0.11 | -33.33 | 2.01E-7 | * | 0.47 |
| Beryllium Compounds | -61.62 | 5.92E-32 | * | 0.04 | -39.72 | 2.53E-10 | * | 0.47 |
| Rmp Facility Proximity | -111.85 | 1.24E-69 | * | 0.09 | -65.67 | 1.40E-25 | * | 0.47 |
| Benzene | -97.84 | 3.34E-19 | * | 0.02 | -87.47 | 9.37E-8 | * | 0.47 |
| Ethylene Glycol | -107.04 | 3.86E-102 | * | 0.14 | -106.57 | 2.20E-5 | * | 0.47 |

2. Hypertension

| Variable | Univariate β | *p* | $p_{adj}$ | $r^2$ | Multivariate β | *p* | $p_{adj}$ | $r^2$ |
|---|---|---|---|---|---|---|---|---|
| Lead Paint | -91.73 | 5.78E-11 | * | 0.01 | 149.54 | 3.72E-26 | * | 0.56 |
| 4-Nitrophenol | -121.64 | 6.63E-72 | * | 0.10 | 72.81 | 7.04E-7 | * | 0.56 |
| Superfund Proximity | -117.17 | 1.69E-56 | * | 0.08 | -28.70 | 1.23E-6 | * | 0.56 |
| Pahpom | -109.36 | 1.92E-19 | * | 0.03 | -39.47 | 5.67E-6 | * | 0.56 |
| Beryllium Compounds | -79.76 | 3.48E-28 | * | 0.04 | -40.36 | 2.99E-7 | * | 0.56 |
| Rmp Facility Proximity | -123.64 | 1.85E-44 | * | 0.06 | -46.33 | 3.40E-9 | * | 0.56 |
| Underground Storage Tanks | -183.36 | 2.33E-124 | * | 0.16 | -81.10 | 1.37E-23 | * | 0.56 |
| Ethylbenzene | -193.70 | 1.03E-100 | * | 0.13 | -283.12 | 9.40E-12 | * | 0.56 |

3. Asthma

| Variable | Univariate β | *p* | $p_{adj}$ | $r^2$ | Multivariate β | *p* | $p_{adj}$ | $r^2$ |
|---|---|---|---|---|---|---|---|---|
| Acetaldehyde | 39.64 | 6.70E-61 | * | 0.08 | 134.71 | 3.13E-36 | * | 0.28 |
| Ozone | 2.36 | 4.19E-1 | | -0.00 | 32.51 | 2.67E-5 | * | 0.28 |
| Xylenes (Mixed Isomers) | 1.26 | 2.97E-1 | | 0.00 | 13.84 | 4.16E-6 | * | 0.28 |
| Hazardous Waste Proximity | 0.14 | 8.91E-1 | | -0.00 | 8.61 | 2.50E-8 | * | 0.28 |
| Underground Storage Tanks | -0.69 | 5.44E-1 | | -0.00 | 6.43 | 1.08E-5 | * | 0.28 |
| Tetrachloroethylene | 2.49 | 9.33E-3 | | 0.00 | 4.76 | 2.20E-5 | * | 0.28 |
| Wastewater Discharge | 4.01 | 1.21E-5 | * | 0.01 | 4.05 | 3.81E-7 | * | 0.28 |
| Lead Paint | -15.82 | 1.28E-15 | * | 0.02 | -12.17 | 1.81E-6 | * | 0.28 |
| Rmp Facility Proximity | -17.51 | 1.43E-44 | * | 0.06 | -12.33 | 6.32E-18 | * | 0.28 |
| Benzene | -0.55 | 7.96E-1 | | -0.00 | -17.59 | 2.49E-6 | * | 0.28 |
| Ethylene Glycol | -3.65 | 3.04E-4 | | 0.00 | -30.53 | 1.01E-7 | * | 0.28 |
| Formaldehyde | 33.82 | 1.45E-33 | * | 0.05 | -144.16 | 3.68E-29 | * | 0.28 |

4. Cancer

| Variable | Univariate β | *p* | $p_{adj}$ | $r^2$ | Multivariate β | *p* | $p_{adj}$ | $r^2$ |
|---|---|---|---|---|---|---|---|---|
| Carbon Tetrachloride | -12.65 | 5.76E-1 | | -0.00 | 202.86 | 6.19E-8 | * | 0.41 |
| Acetaldehyde | -34.53 | 5.24E-29 | * | 0.04 | 57.45 | 2.12E-6 | * | 0.41 |
| Lead Paint | 43.47 | 6.84E-70 | * | 0.09 | 53.71 | 3.90E-72 | * | 0.41 |
| Methyl Chloride (Chloromethane) | 15.06 | 3.76E-2 | | 0.00 | 39.75 | 3.29E-8 | * | 0.41 |
| 2,2,4-Trimethylpentane | -24.99 | 2.43E-59 | * | 0.08 | 36.59 | 3.41E-28 | * | 0.41 |
| 4-Nitrophenol | -18.41 | 5.67E-51 | * | 0.07 | 21.29 | 3.26E-12 | * | 0.41 |
| Acetonitrile | 4.02 | 2.12E-2 | | 0.00 | 20.32 | 4.71E-9 | * | 0.41 |
| Traffic Proximity | -13.12 | 1.56E-28 | * | 0.04 | -5.50 | 7.56E-6 | * | 0.41 |
| Beryllium Compounds | -11.76 | 1.84E-19 | * | 0.03 | -6.96 | 2.11E-5 | * | 0.41 |
| Rmp Facility Proximity | -11.90 | 1.17E-13 | * | 0.02 | -10.89 | 2.40E-11 | * | 0.41 |
| Underground Storage Tanks | -26.12 | 1.65E-76 | * | 0.10 | -17.90 | 2.31E-26 | * | 0.41 |
| Particulate Matter 2.5 | -33.13 | 1.82E-22 | * | 0.03 | -43.29 | 1.15E-7 | * | 0.41 |
| Formaldehyde | -53.24 | 2.11E-51 | * | 0.07 | -72.59 | 6.43E-7 | * | 0.41 |

5. Coronary Heart Disease

| Variable | Univariate β | *p* | $p_{adj}$ | $r^2$ | Multivariate β | *p* | $p_{adj}$ | $r^2$ |
|---|---|---|---|---|---|---|---|---|
| Carbon Tetrachloride | 2.37 | 9.39E-1 | | -0.00 | 242.91 | 5.45E-7 | * | 0.47 |
| Lead Paint | 12.63 | 2.22E-4 | | 0.00 | 55.93 | 3.46E-48 | * | 0.47 |
| 4-Nitrophenol | -33.83 | 1.36E-94 | * | 0.13 | 28.51 | 5.74E-13 | * | 0.47 |
| Xylenes (Mixed Isomers) | -43.23 | 1.29E-103 | * | 0.14 | 21.94 | 8.36E-7 | * | 0.47 |
| Beryllium Compounds | -22.70 | 4.20E-38 | * | 0.05 | -10.25 | 1.30E-6 | * | 0.47 |
| Rmp Facility Proximity | -33.37 | 1.61E-54 | * | 0.07 | -21.41 | 5.44E-24 | * | 0.47 |
| Benzene | -55.54 | 8.78E-53 | * | 0.07 | -27.26 | 8.26E-7 | * | 0.47 |

6. COPD

| Variable | Univariate β | *p* | $p_{adj}$ | $r^2$ | Multivariate β | *p* | $p_{adj}$ | $r^2$ |
|---|---|---|---|---|---|---|---|---|
| Carbon Tetrachloride | 226.64 | 1.81E-7 | * | 0.01 | 407.29 | 1.03E-8 | * | 0.43 |
| Lead Paint | -22.17 | 4.74E-6 | * | 0.01 | 34.87 | 3.05E-10 | * | 0.43 |
| Xylenes (Mixed Isomers) | -40.20 | 8.35E-44 | * | 0.06 | 34.35 | 1.43E-7 | * | 0.43 |
| 4-Nitrophenol | -37.96 | 1.65E-58 | * | 0.08 | 33.56 | 6.83E-9 | * | 0.43 |
| Cyanide Compounds | -13.52 | 4.01E-8 | * | 0.01 | 14.86 | 1.75E-6 | * | 0.43 |
| 2-Chloroacetophenone | -5.48 | 1.57E-2 | | 0.00 | 11.67 | 4.62E-6 | * | 0.43 |
| Wastewater Discharge | 11.20 | 5.49E-7 | * | 0.01 | 9.27 | 8.62E-8 | * | 0.43 |
| Methylhydrazine | -21.05 | 3.53E-20 | * | 0.03 | -12.72 | 4.80E-6 | * | 0.43 |
| Beryllium Compounds | -27.59 | 2.89E-28 | * | 0.04 | -16.04 | 2.39E-7 | * | 0.43 |
| Rmp Facility Proximity | -52.86 | 2.53E-68 | * | 0.09 | -32.95 | 3.14E-26 | * | 0.43 |
| Benzene | -56.22 | 2.76E-27 | * | 0.04 | -45.96 | 1.46E-8 | * | 0.43 |

7. Depression

| Variable | Univariate β | *p* | $p_{adj}$ | $r^2$ | Multivariate β | *p* | $p_{adj}$ | $r^2$ |
|---|---|---|---|---|---|---|---|---|
| Methyl Chloride (Chloromethane) | 141.39 | 4.93E-13 | * | 0.02 | 106.84 | 1.73E-7 | * | 0.35 |
| Hazardous Waste Proximity | -18.95 | 3.36E-8 | * | 0.01 | 22.81 | 5.43E-6 | * | 0.35 |
| Nickel Compounds | 26.76 | 3.69E-15 | * | 0.02 | 17.11 | 3.39E-6 | * | 0.35 |
| Dibenzofuran | -0.59 | 8.60E-1 | | -0.00 | -20.84 | 1.56E-11 | * | 0.35 |
| Polychlorinated Biphenyls (Aroclors) | -20.00 | 1.12E-9 | * | 0.01 | -24.45 | 1.84E-15 | * | 0.35 |
| Rmp Facility Proximity | -59.35 | 2.48E-43 | * | 0.06 | -29.88 | 1.10E-10 | * | 0.35 |
| Lead Paint | -99.60 | 9.52E-50 | * | 0.07 | -47.08 | 1.36E-8 | * | 0.35 |
| Ethylene Glycol | -27.58 | 1.48E-15 | * | 0.02 | -82.99 | 8.34E-6 | * | 0.35 |

8. Diabetes

| Variable | Univariate β | *p* | $p_{adj}$ | $r^2$ | Multivariate β | *p* | $p_{adj}$ | $r^2$ |
|---|---|---|---|---|---|---|---|---|
| Formaldehyde | 174.13 | 1.31E-110 | * | 0.15 | 352.87 | 7.34E-29 | * | 0.46 |
| Lead Paint | -47.50 | 4.45E-17 | * | 0.02 | 53.05 | 3.73E-17 | * | 0.46 |
| Xylenes (Mixed Isomers) | -43.94 | 3.24E-38 | * | 0.05 | 32.76 | 9.58E-6 | * | 0.46 |
| 4-Nitrophenol | -34.41 | 3.68E-35 | * | 0.05 | 27.72 | 2.39E-5 | * | 0.46 |
| Polychlorinated Biphenyls (Aroclors) | -15.29 | 2.22E-8 | * | 0.01 | -13.56 | 5.26E-9 | * | 0.46 |
| Rmp Facility Proximity | -49.44 | 1.90E-43 | * | 0.06 | -24.62 | 2.28E-12 | * | 0.46 |
| Benzene | -95.79 | 5.54E-57 | * | 0.08 | -62.42 | 1.25E-11 | * | 0.46 |
| Diesel Pm | -60.47 | 5.12E-44 | * | 0.06 | -69.40 | 1.40E-8 | * | 0.46 |
| Ozone | -10.94 | 1.89E-1 | | 0.00 | -90.09 | 2.29E-6 | * | 0.46 |

9. Renal Disease

| Variable | Univariate β | *p* | $p_{adj}$ | $r^2$ | Multivariate β | *p* | $p_{adj}$ | $r^2$ |
|---|---|---|---|---|---|---|---|---|
| Formaldehyde | 20.48 | 2.98E-30 | * | 0.04 | 40.00 | 2.78E-8 | * | 0.44 |
| Lead Paint | -1.48 | 2.43E-1 | | 0.00 | 19.32 | 4.06E-40 | * | 0.44 |
| 4-Nitrophenol | -9.78 | 1.28E-56 | * | 0.08 | 9.63 | 1.69E-10 | * | 0.44 |
| Xylenes (Mixed Isomers) | -14.35 | 4.20E-82 | * | 0.11 | 7.94 | 2.88E-6 | * | 0.44 |
| Rmp Facility Proximity | -12.23 | 3.14E-53 | * | 0.07 | -7.29 | 1.71E-19 | * | 0.44 |
| Benzene | -24.92 | 1.04E-77 | * | 0.10 | -14.92 | 1.69E-12 | * | 0.44 |
| Diesel Pm | -21.17 | 1.91E-110 | * | 0.15 | -16.54 | 3.73E-9 | * | 0.44 |

10. Obesity

| Variable | Univariate β | *p* | $p_{adj}$ | $r^2$ | Multivariate β | *p* | $p_{adj}$ | $r^2$ |
|---|---|---|---|---|---|---|---|---|
| Carbon Tetrachloride | 1485.46 | 1.14E-62 | * | 0.08 | 1199.37 | 2.49E-17 | * | 0.48 |
| Formaldehyde | 293.95 | 1.37E-98 | * | 0.13 | 292.34 | 1.02E-7 | * | 0.48 |
| Particulate Matter 2.5 | 237.05 | 8.44E-70 | * | 0.09 | 158.59 | 2.49E-7 | * | 0.48 |
| Cyanide Compounds | -9.13 | 7.66E-2 | | 0.00 | 28.69 | 3.30E-6 | * | 0.48 |
| Hazardous Waste Proximity | -80.40 | 3.29E-58 | * | 0.08 | -35.23 | 1.18E-7 | * | 0.48 |
| 2,2,4-Trimethylpentane | -109.49 | 1.03E-70 | * | 0.10 | -85.49 | 6.24E-12 | * | 0.48 |

11. Stroke

| Variable | Univariate β | *p* | $p_{adj}$ | $r^2$ | Multivariate β | *p* | $p_{adj}$ | $r^2$ |
|---|---|---|---|---|---|---|---|---|
| Carbon Tetrachloride | 43.11 | 1.01E-2 | | 0.00 | 113.57 | 1.66E-5 | * | 0.47 |
| Acetaldehyde | 44.84 | 5.33E-89 | * | 0.12 | 51.35 | 1.85E-9 | * | 0.47 |
| Lead Paint | -8.24 | 1.01E-5 | * | 0.01 | 21.86 | 4.30E-26 | * | 0.47 |
| 4-Nitrophenol | -13.88 | 1.24E-52 | * | 0.07 | 11.36 | 1.25E-7 | * | 0.47 |
| Xylenes (Mixed Isomers) | -19.64 | 1.20E-70 | * | 0.10 | 10.68 | 1.04E-5 | * | 0.47 |
| Rmp Facility Proximity | -18.78 | 5.82E-58 | * | 0.08 | -10.59 | 3.73E-20 | * | 0.47 |
| Benzene | -32.94 | 2.47E-62 | * | 0.08 | -17.87 | 2.96E-9 | * | 0.47 |

12. Stroke Mortality

| Variable | Univariate β | *p* | $p_{adj}$ | $r^2$ | Multivariate β | *p* | $p_{adj}$ | $r^2$ |
|---|---|---|---|---|---|---|---|---|
| Formaldehyde | 846.64 | 8.70E-250 | * | 0.30 | 565.81 | 2.77E-7 | * | 0.42 |
| Methanol | 683.54 | 1.74E-211 | * | 0.26 | 210.93 | 1.12E-5 | * | 0.42 |
| Underground Storage Tanks | -27.63 | 1.23E-2 | | 0.00 | 62.38 | 7.84E-7 | * | 0.42 |
| Propylene Dichloride (1,2-Dichloropropane) | 91.63 | 1.00E-22 | * | 0.03 | 45.65 | 5.38E-8 | * | 0.42 |
| Rmp Facility Proximity | -105.35 | 5.90E-18 | * | 0.02 | -60.07 | 1.01E-6 | * | 0.42 |

**Supplementary Table 2.** Univariate and multivariate regression showing the top β coefficient values predicting each disease / health indicator (if more than 20 were present, only the top 20 were presented). * = statistically significant with Bonferroni-adjustment ($p_{adj}$).

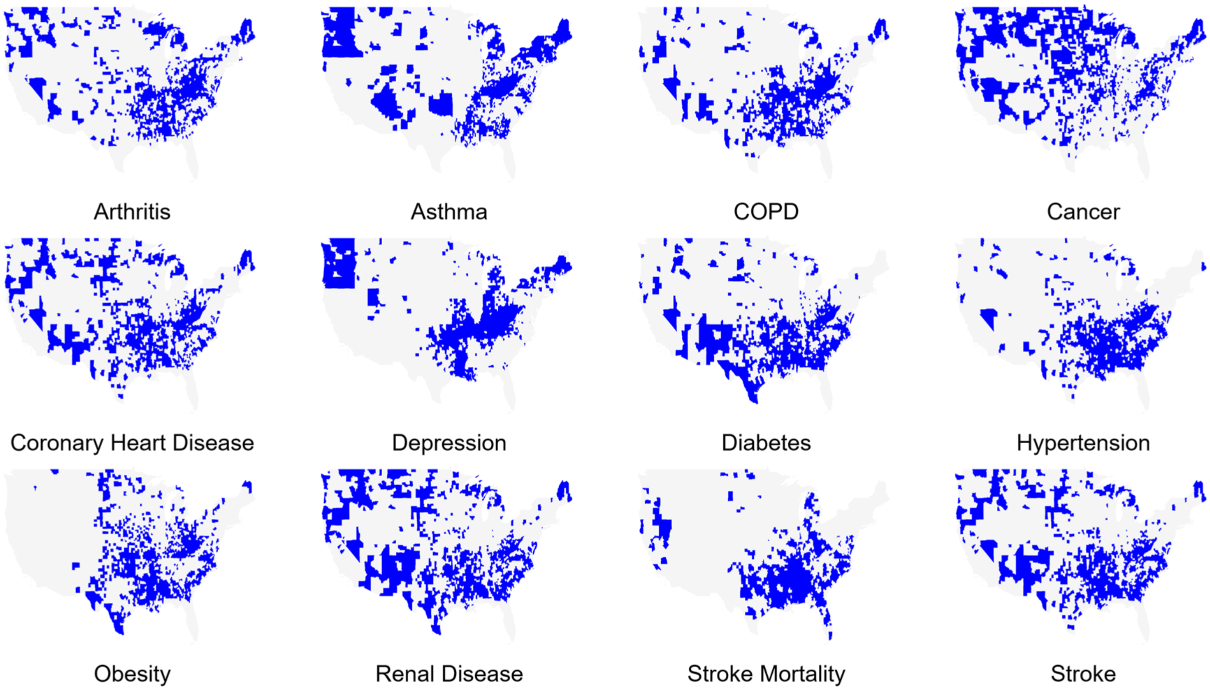


**Supplementary Figure 1.** Chronic disease reference maps where counties with rates ≥ 70th percentile are highlighted in blue.

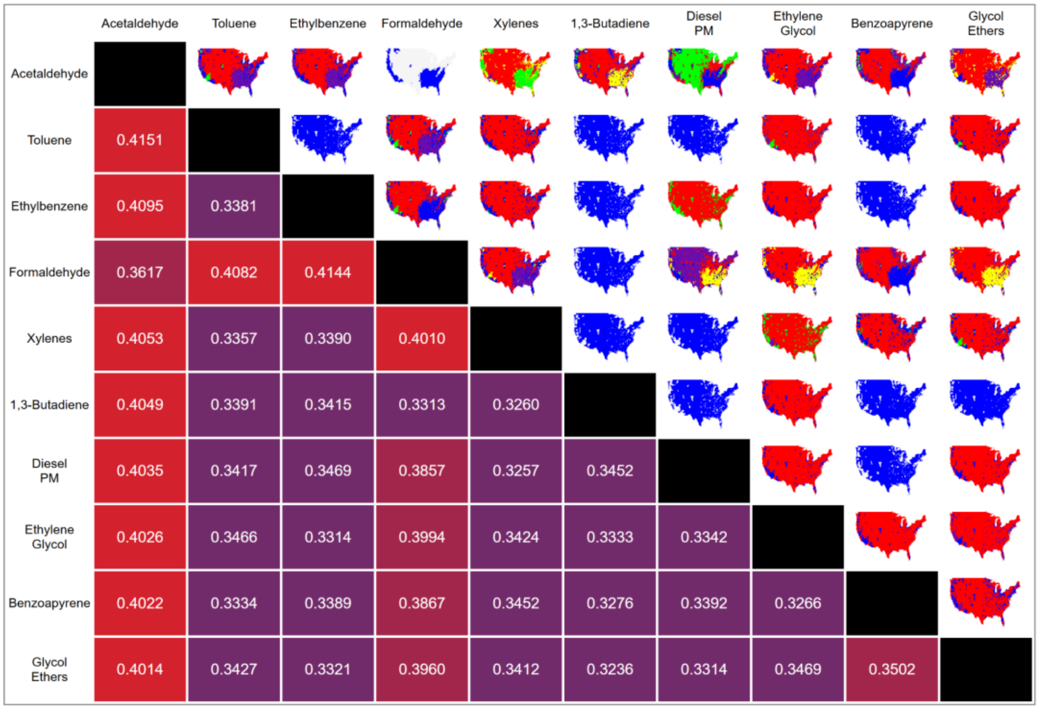


Arthritis

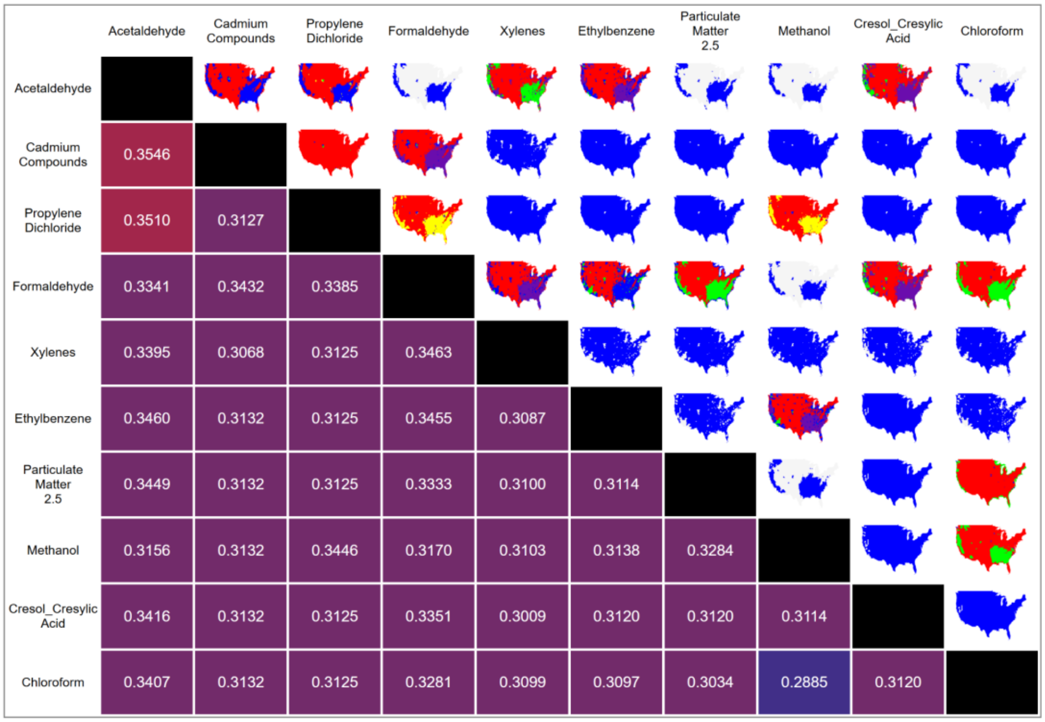


Asthma

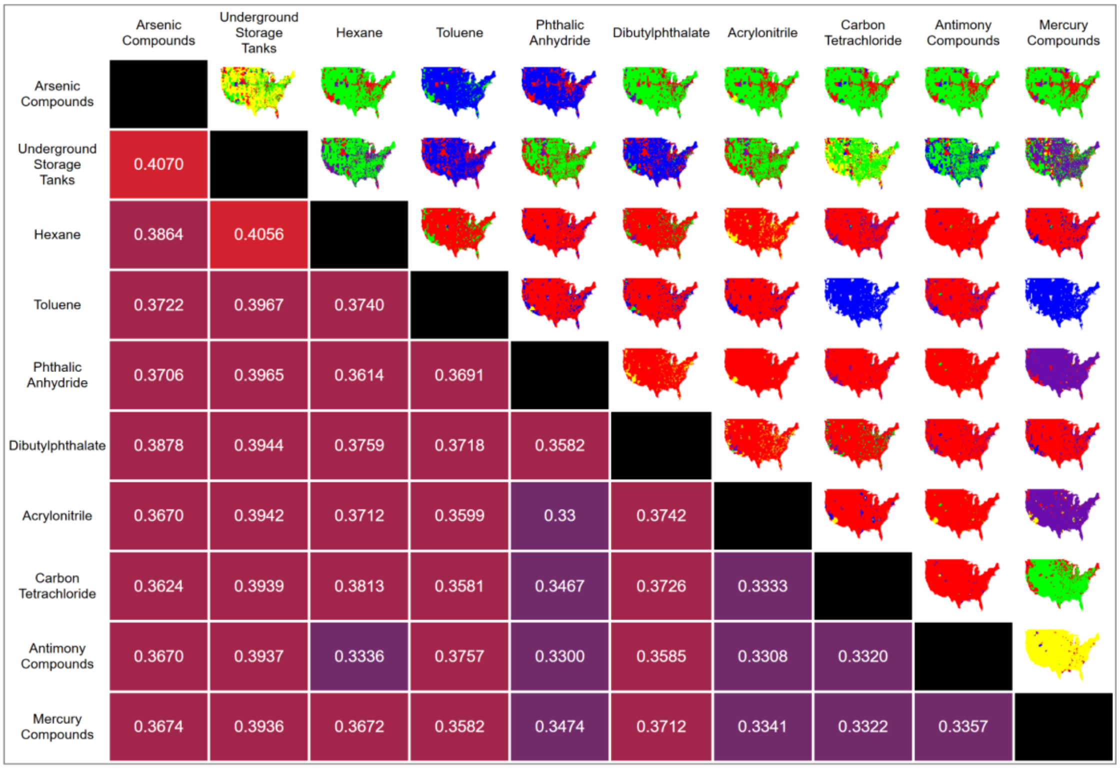
Arsenic Compounds
Underground Storage Tanks
Hexane
Toluene
Phthalic Anhydride
Dibutylphthalate
Acrylonitrile
Carbon Tetrachloride
Antimony Compounds
Mercury Compounds
Arsenic Compounds
Underground Storage Tanks 0.4070
Hexane 0.3864 0.4056
Toluene 0.3722 0.3967 0.3740
Phthalic Anhydride 0.3706 0.3965 0.3614 0.3691
Dibutylphthalate 0.3878 0.3944 0.3759 0.3718 0.3582
Acrylonitrile 0.3670 0.3942 0.3712 0.3599 0.33 0.3742
Carbon Tetrachloride 0.3624 0.3939 0.3813 0.3581 0.3467 0.3726 0.3333
Antimony Compounds 0.3670 0.3937 0.3336 0.3757 0.3300 0.3585 0.3308 0.3320
Mercury Compounds 0.3674 0.3936 0.3672 0.3582 0.3474 0.3712 0.3341 0.3322 0.3357

Cancer

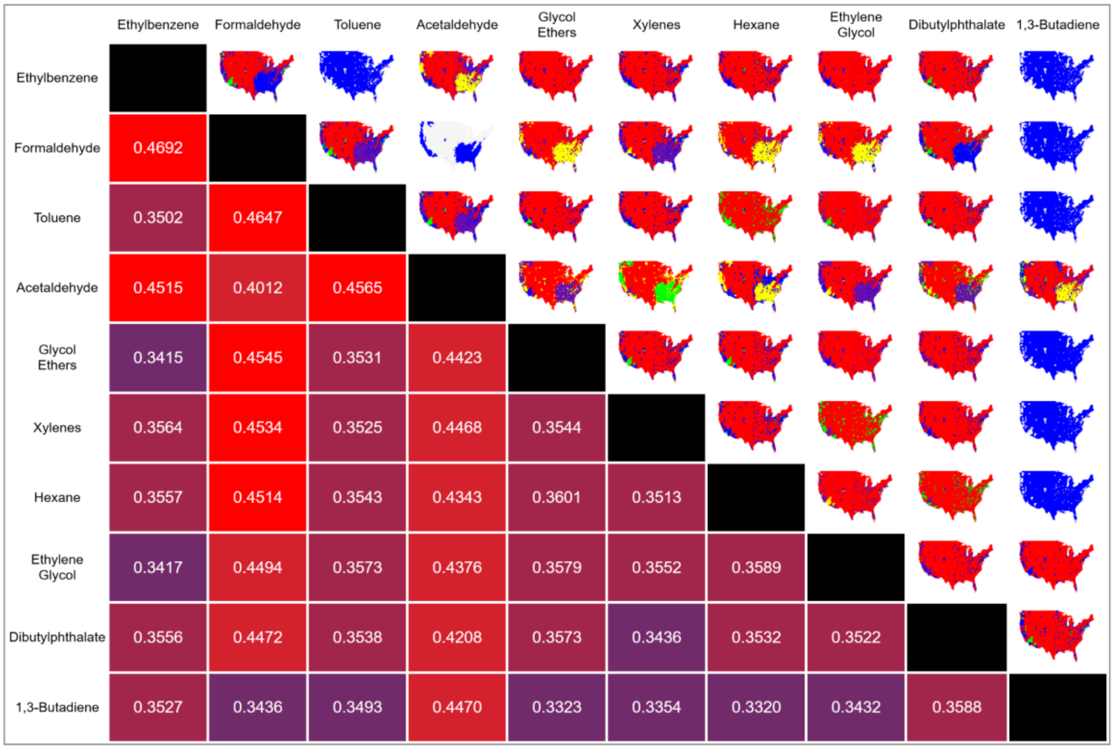
Ethylbenzene
Formaldehyde
Toluene
Acetaldehyde
Glycol Ethers
Xylenes
Hexane
Ethylene Glycol
Dibutylphthalate
1,3-Butadiene
Ethylbenzene
Formaldehyde 0.4692
Toluene 0.3502 0.4647
Acetaldehyde 0.4515 0.4012 0.4565
Glycol Ethers 0.3415 0.4545 0.3531 0.4423
Xylenes 0.3564 0.4534 0.3525 0.4468 0.3544
Hexane 0.3557 0.4514 0.3543 0.4343 0.3601 0.3513
Ethylene Glycol 0.3417 0.4494 0.3573 0.4376 0.3579 0.3552 0.3589
Dibutylphthalate 0.3556 0.4472 0.3538 0.4208 0.3573 0.3436 0.3532 0.3522
1,3-Butadiene 0.3527 0.3436 0.3493 0.4470 0.3323 0.3354 0.3320 0.3432 0.3588

COPD

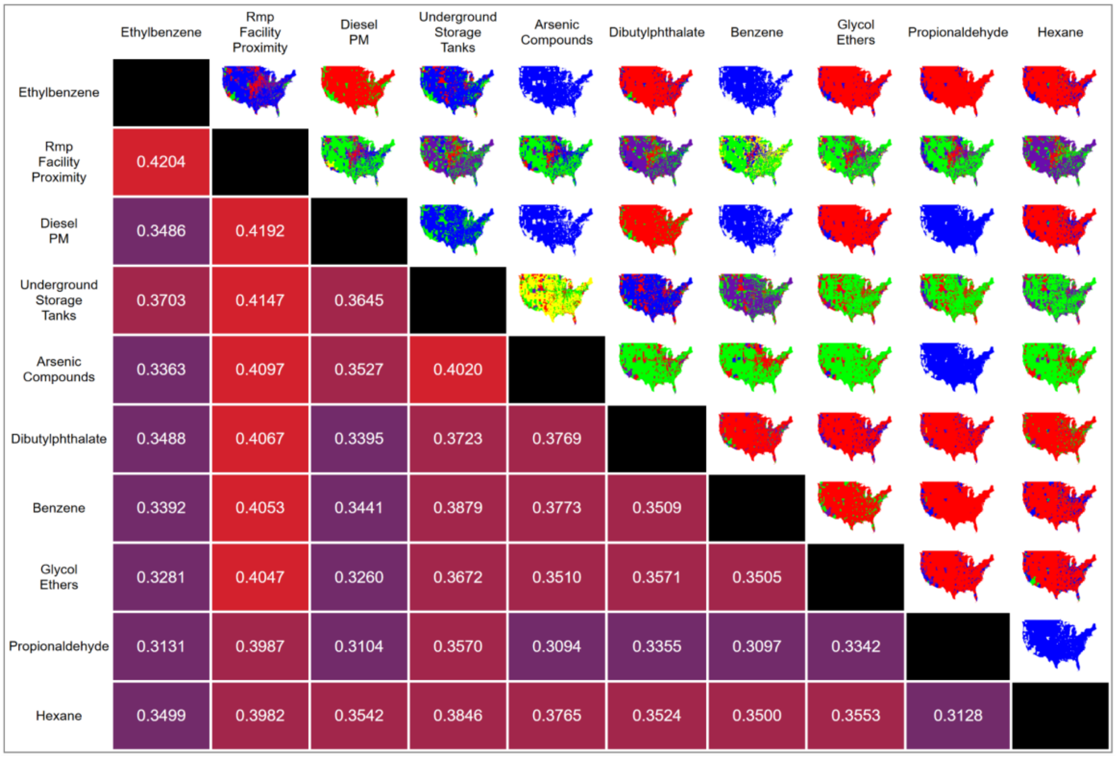


Coronary Artery Disease

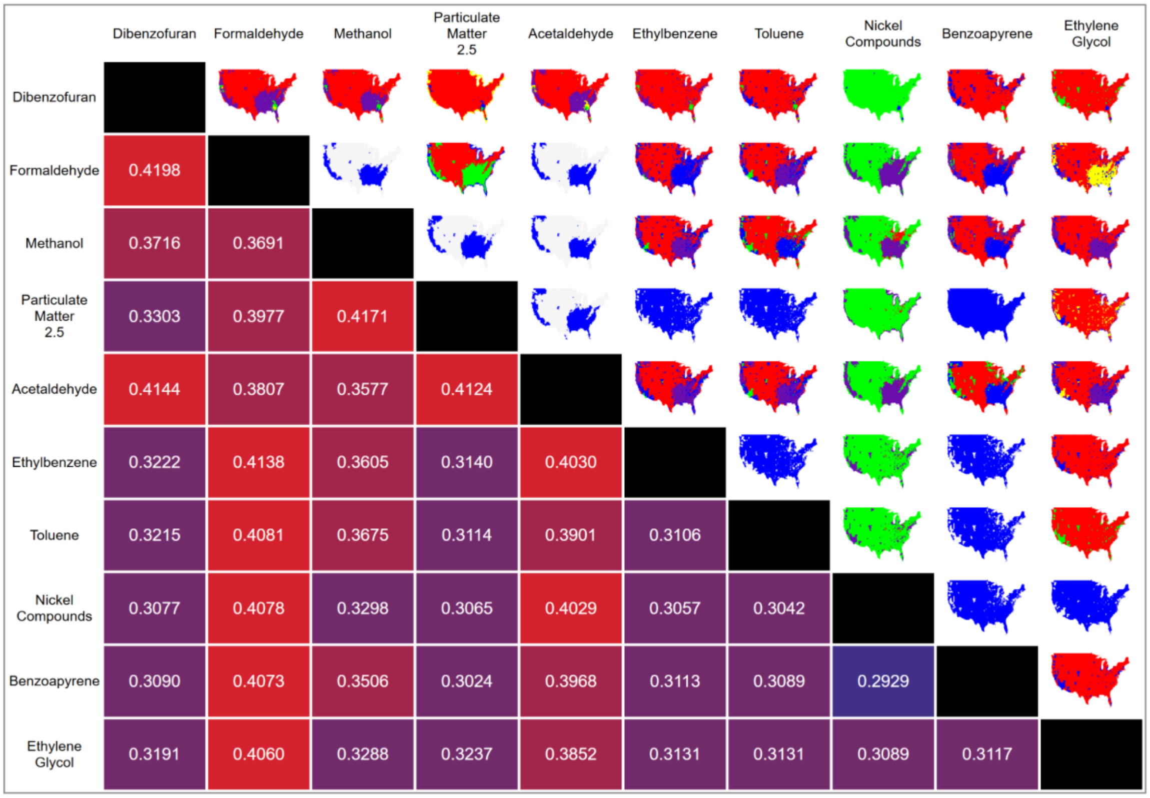


Depression

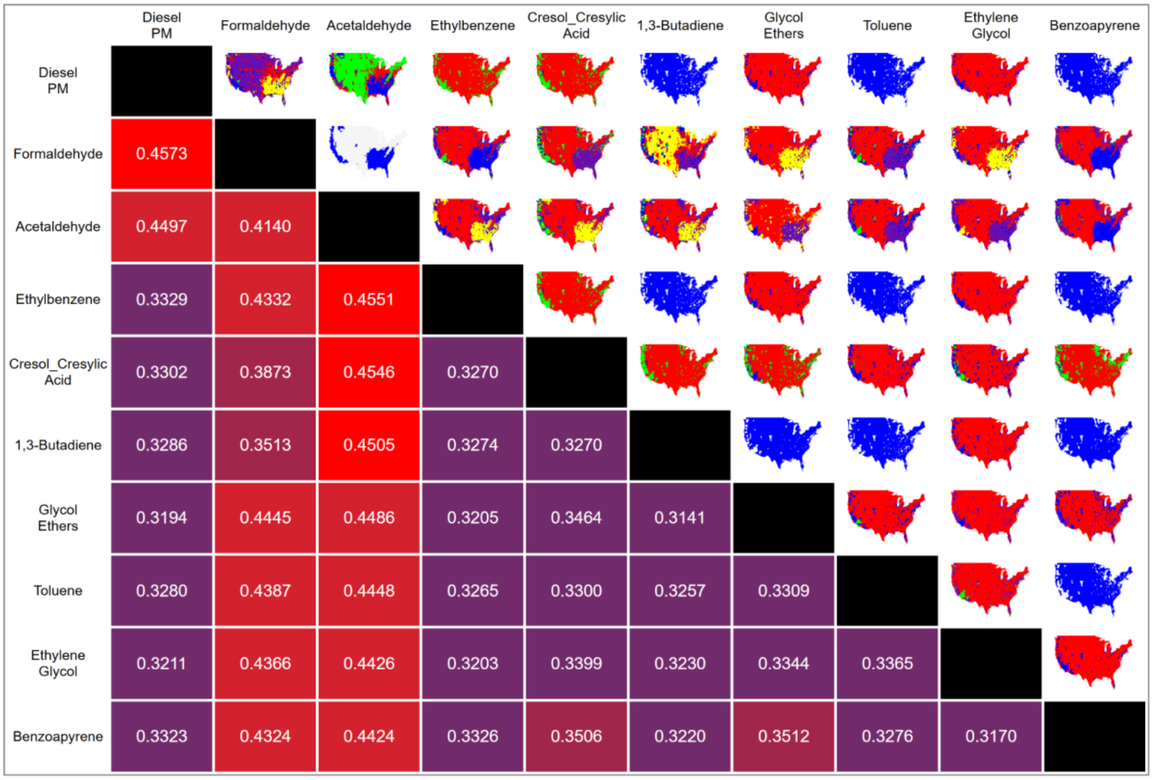


Diabetes

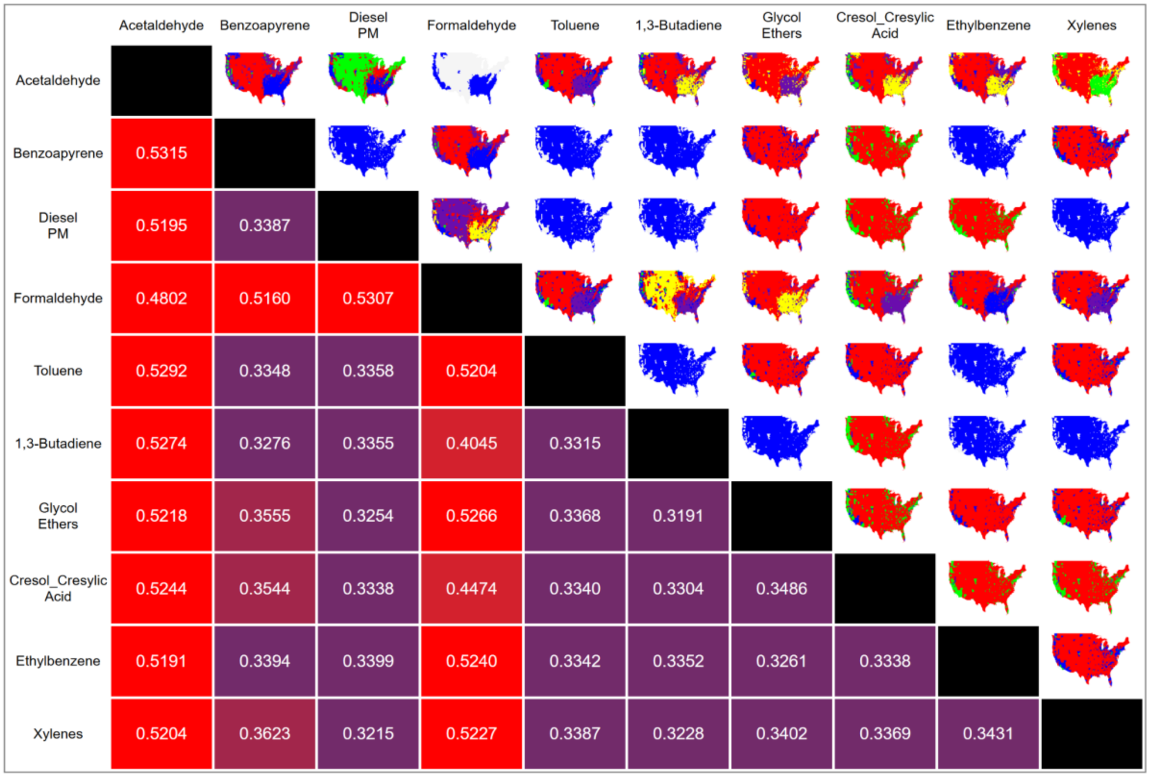


Hypertension

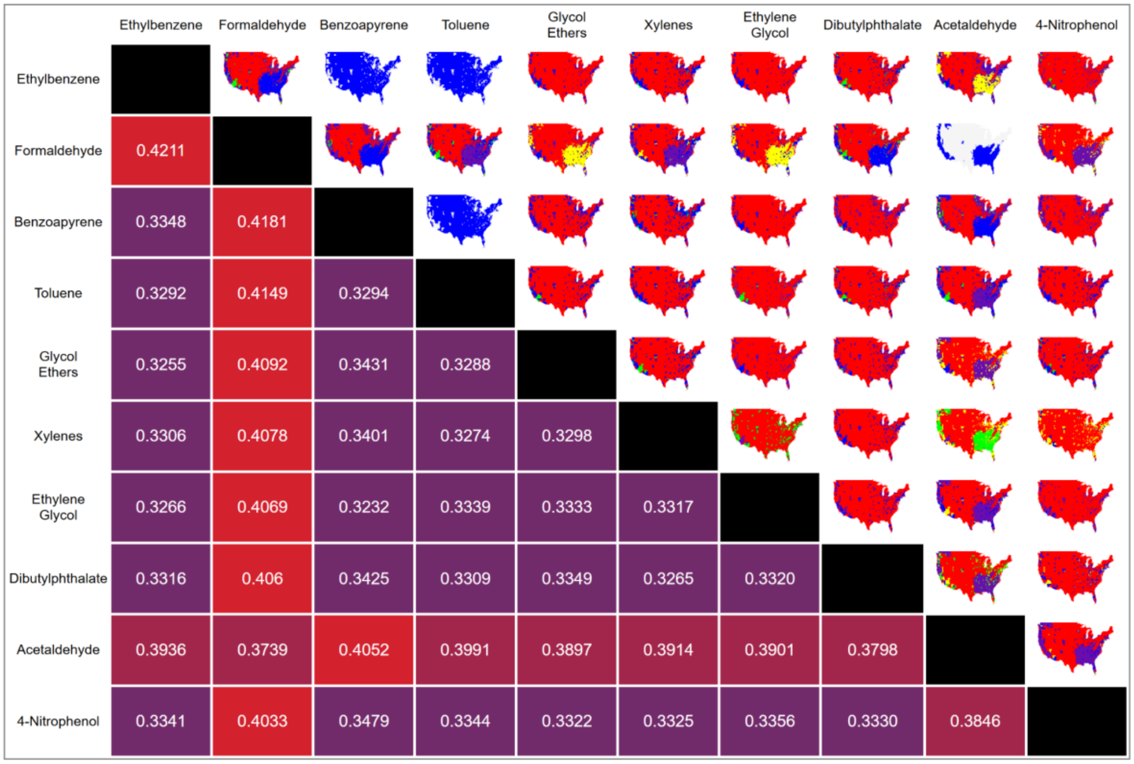


Obesity

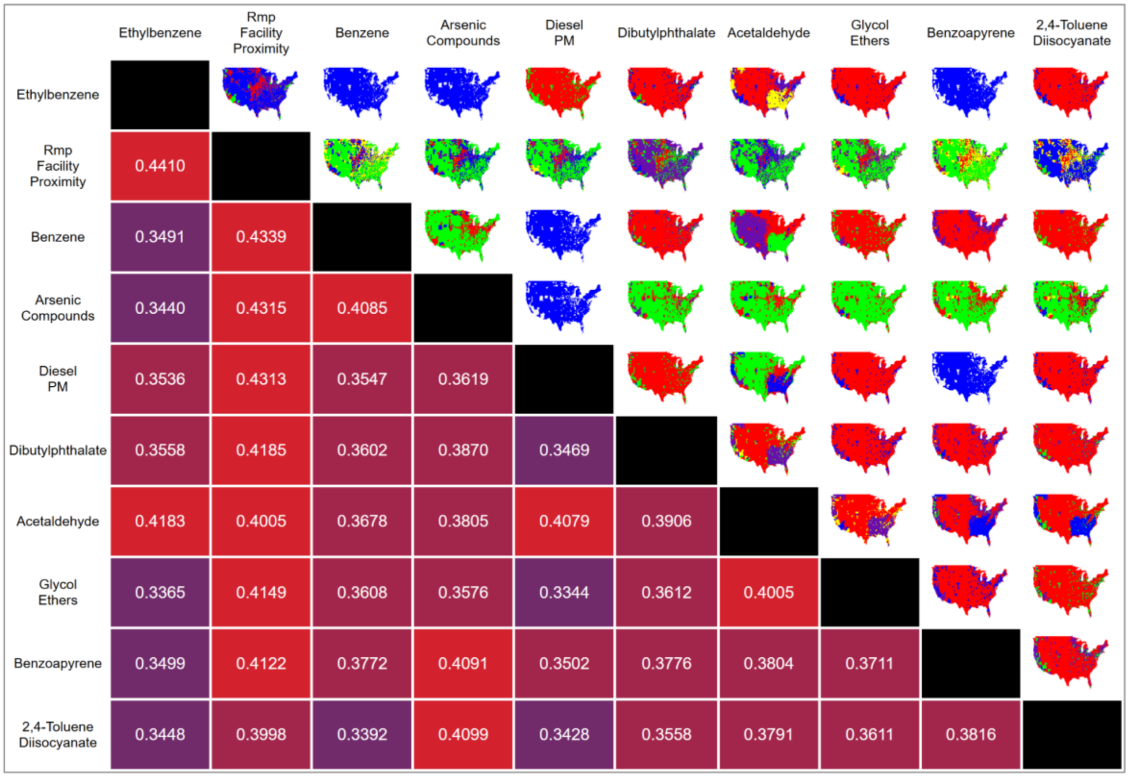


Renal Disease

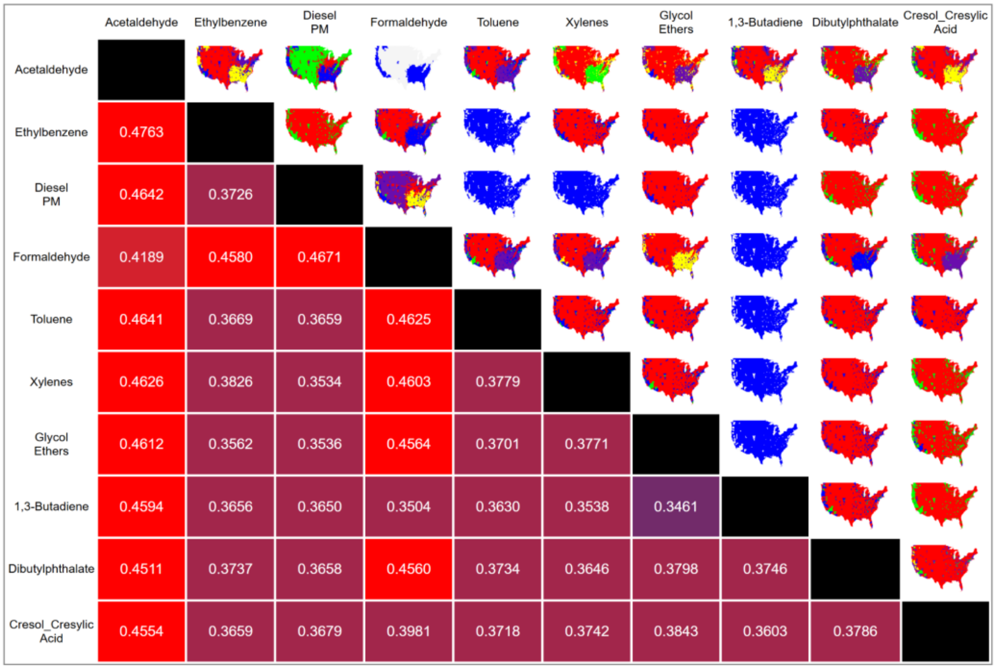


Stroke

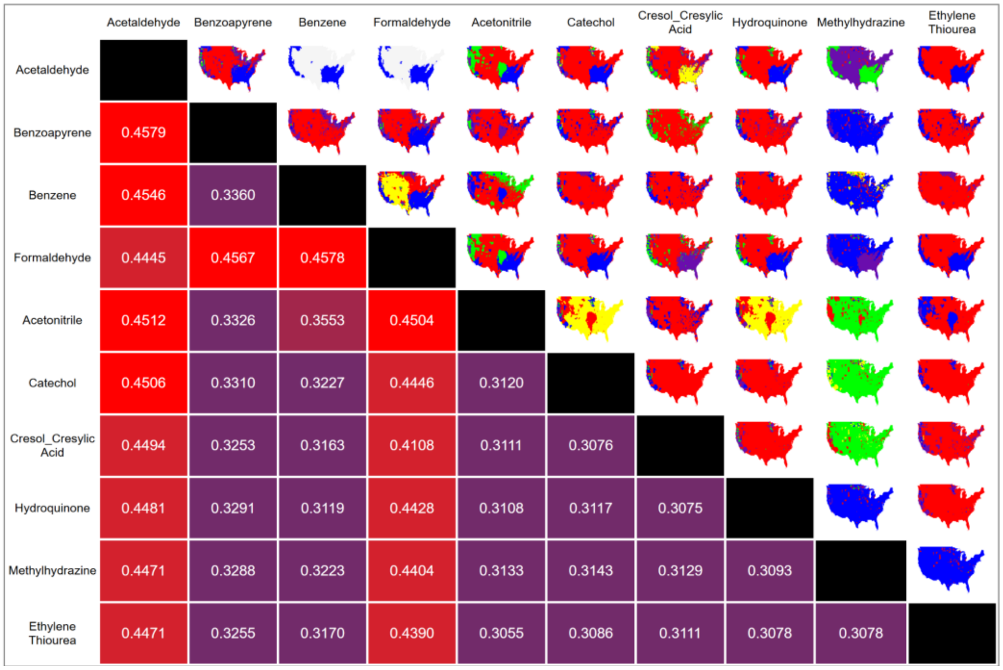


Stroke Mortality

**Supplementary Figure 2. Pairwise pollution correlation matrices for the top 10 pollutants (by Jaccard correlation coefficient) associated with the chronic diseases.** For each pairwise combination of pollutants, a map was calculated using aPEER, and the Jaccard index was calculated relative to a chronic disease (asthma, arthritis, etc). The map with the highest Jaccard index out of the possible maps out of clustering values $k$={2, 3, 4, 5} was then identified for each pairwise combination.

Arthritis

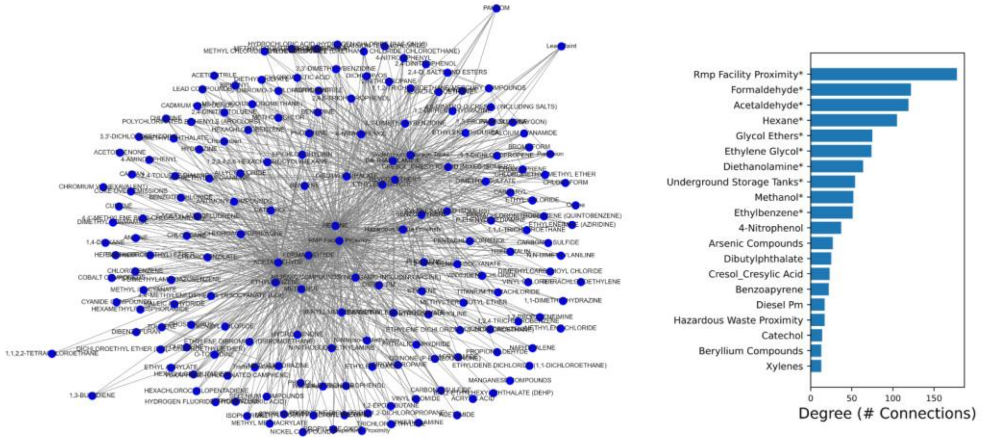


Asthma

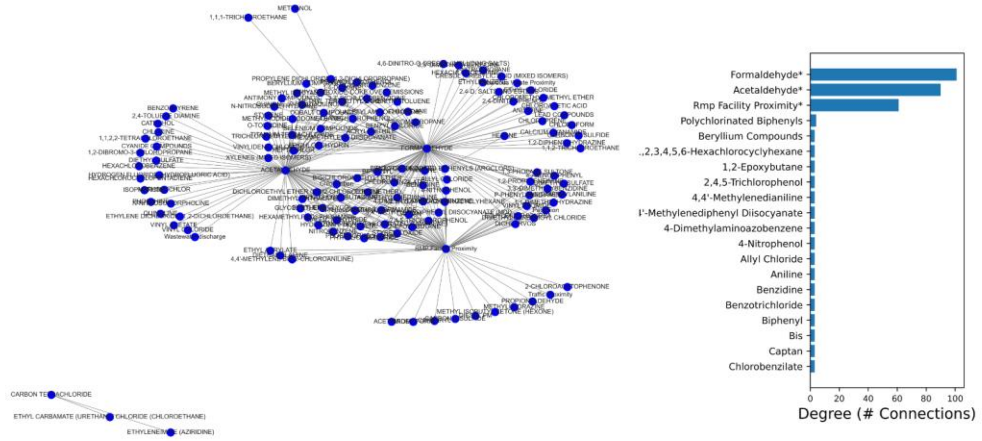


Cancer

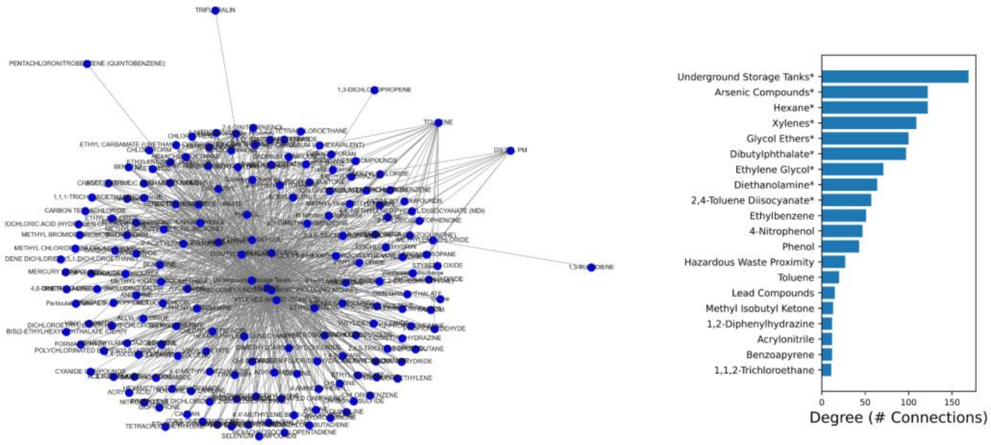

COPD

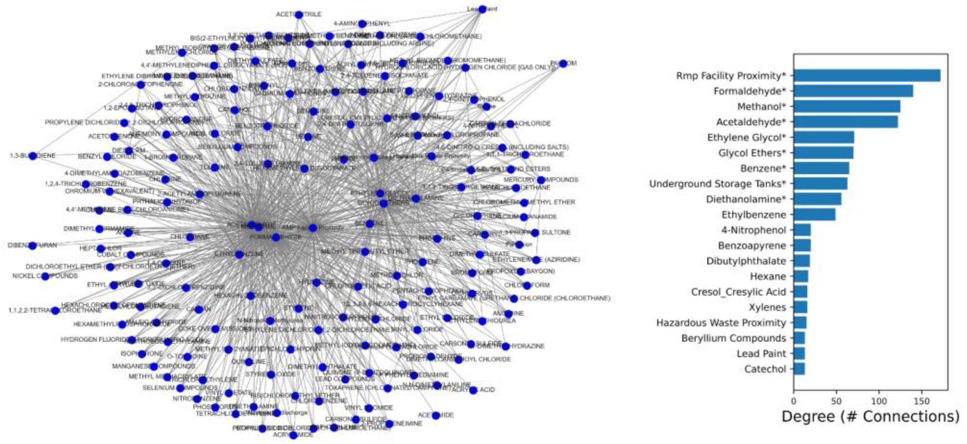


Coronary Heart Disease

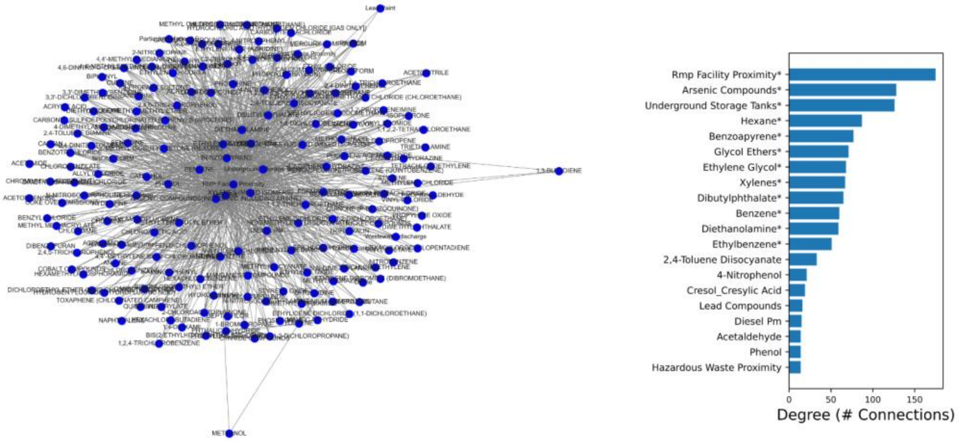


Depression

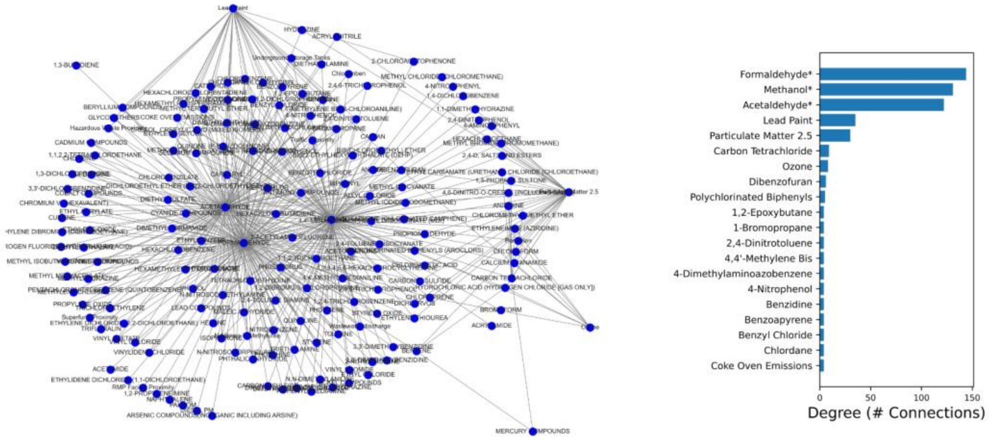

Diabetes

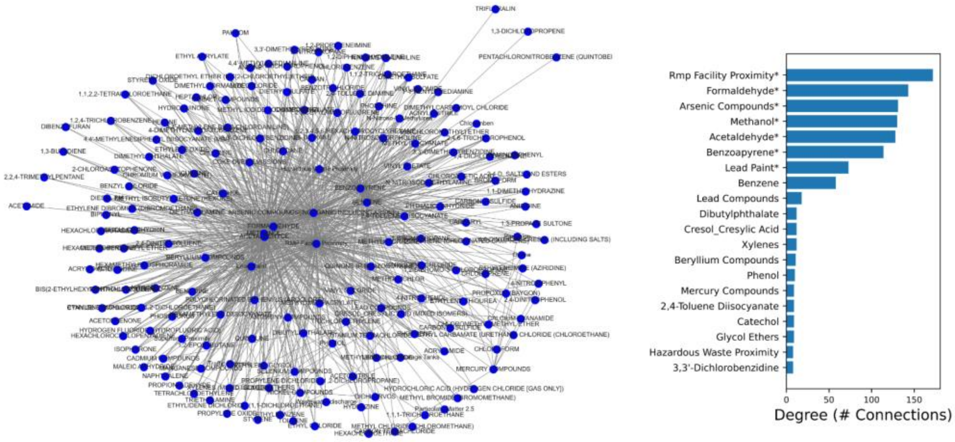


Hypertension

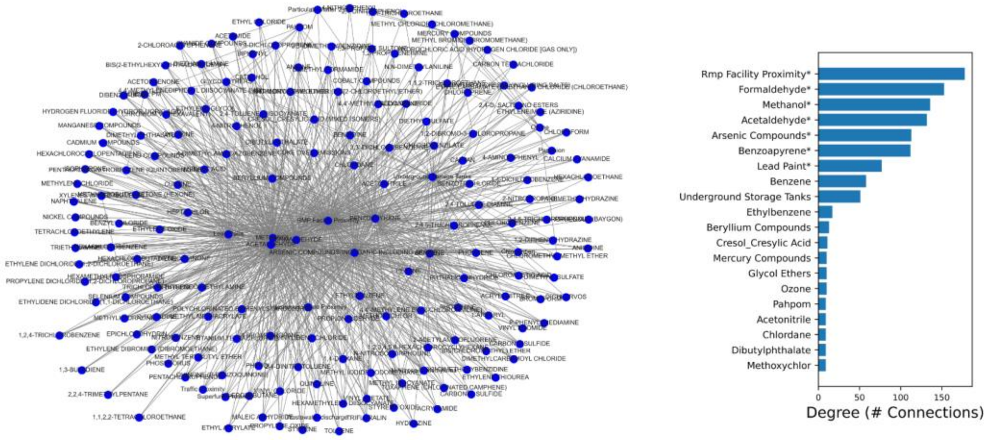


Obesity

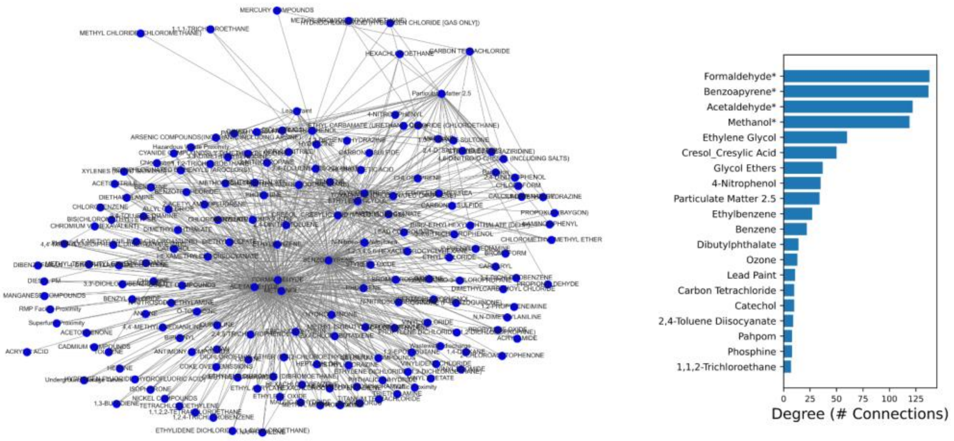

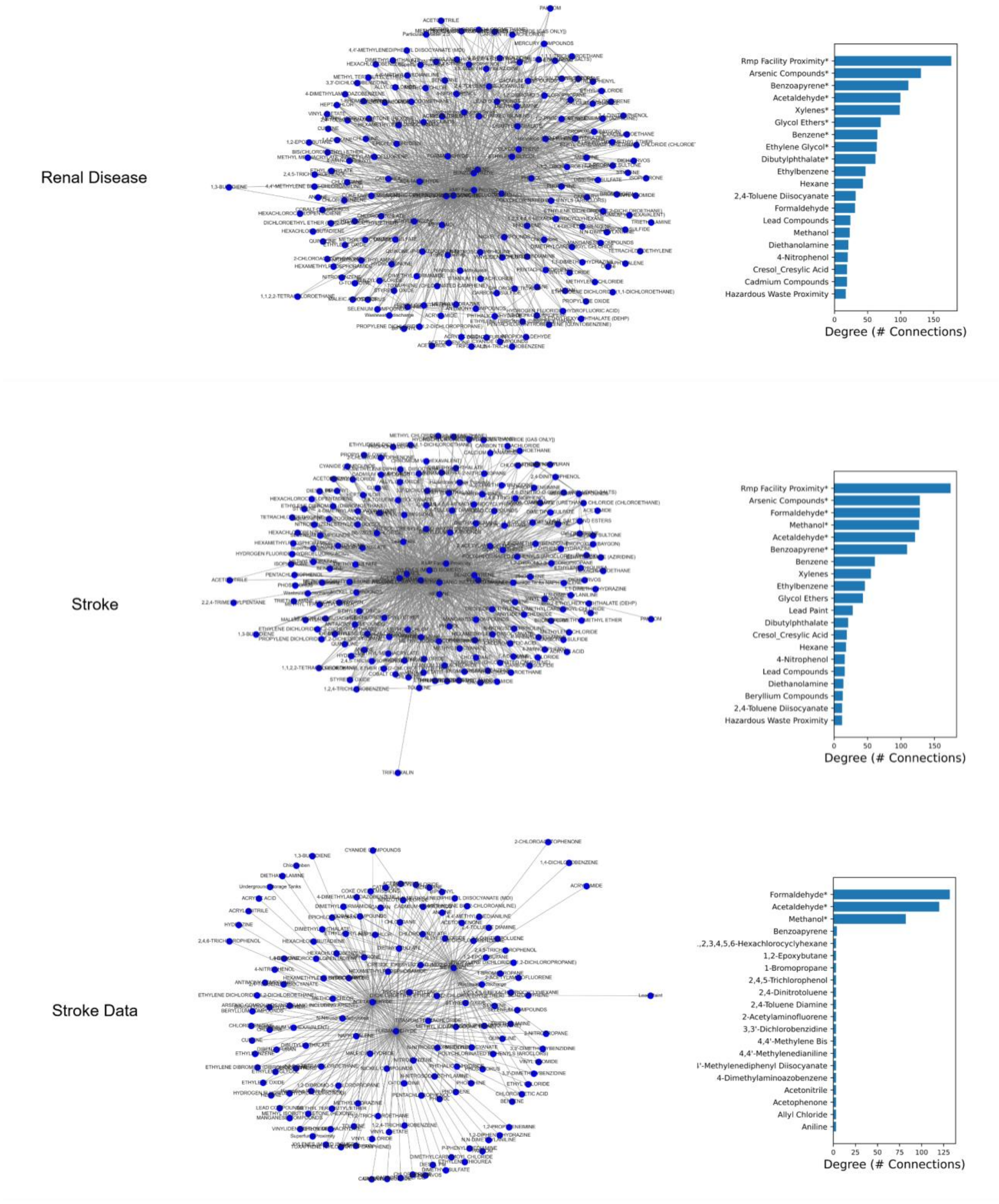


**Supplementary Figure 3**. aPEER pollution networks constructed from Jaccard correlation coefficients for 12 health-related indicators.

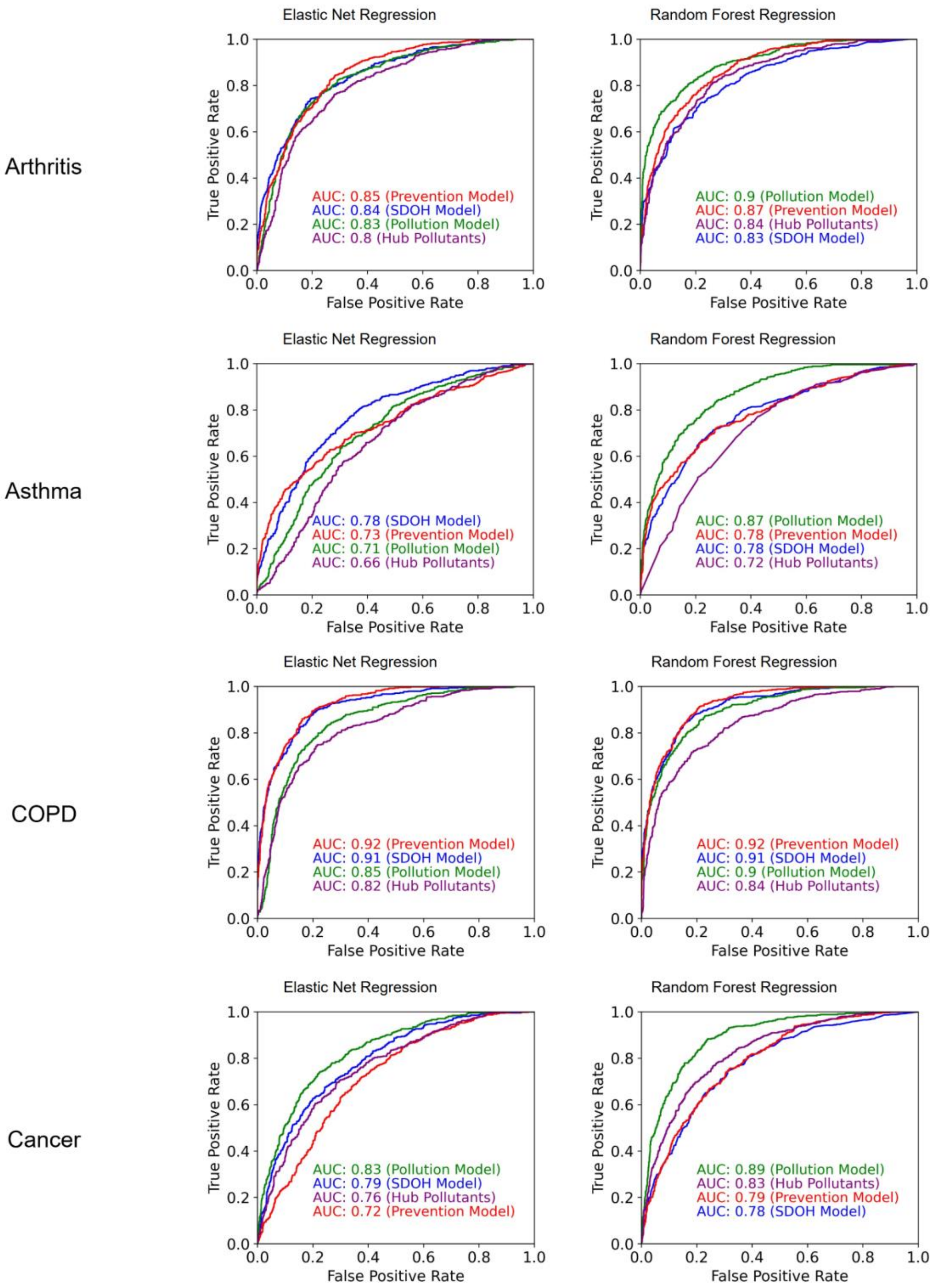
Arthritis
Asthma
COPD
Cancer
Elastic Net Regression
Random Forest Regression
True Positive Rate
False Positive Rate
AUC: 0.85 (Prevention Model)
AUC: 0.84 (SDOH Model)
AUC: 0.83 (Pollution Model)
AUC: 0.8 (Hub Pollutants)
AUC: 0.9 (Pollution Model)
AUC: 0.87 (Prevention Model)
AUC: 0.84 (Hub Pollutants)
AUC: 0.83 (SDOH Model)
AUC: 0.78 (SDOH Model)
AUC: 0.73 (Prevention Model)
AUC: 0.71 (Pollution Model)
AUC: 0.66 (Hub Pollutants)
AUC: 0.87 (Pollution Model)
AUC: 0.78 (Prevention Model)
AUC: 0.78 (SDOH Model)
AUC: 0.72 (Hub Pollutants)
AUC: 0.92 (Prevention Model)
AUC: 0.91 (SDOH Model)
AUC: 0.85 (Pollution Model)
AUC: 0.82 (Hub Pollutants)
AUC: 0.92 (Prevention Model)
AUC: 0.91 (SDOH Model)
AUC: 0.9 (Pollution Model)
AUC: 0.84 (Hub Pollutants)
AUC: 0.83 (Pollution Model)
AUC: 0.79 (SDOH Model)
AUC: 0.76 (Hub Pollutants)
AUC: 0.72 (Prevention Model)
AUC: 0.89 (Pollution Model)
AUC: 0.83 (Hub Pollutants)
AUC: 0.79 (Prevention Model)
AUC: 0.78 (SDOH Model)

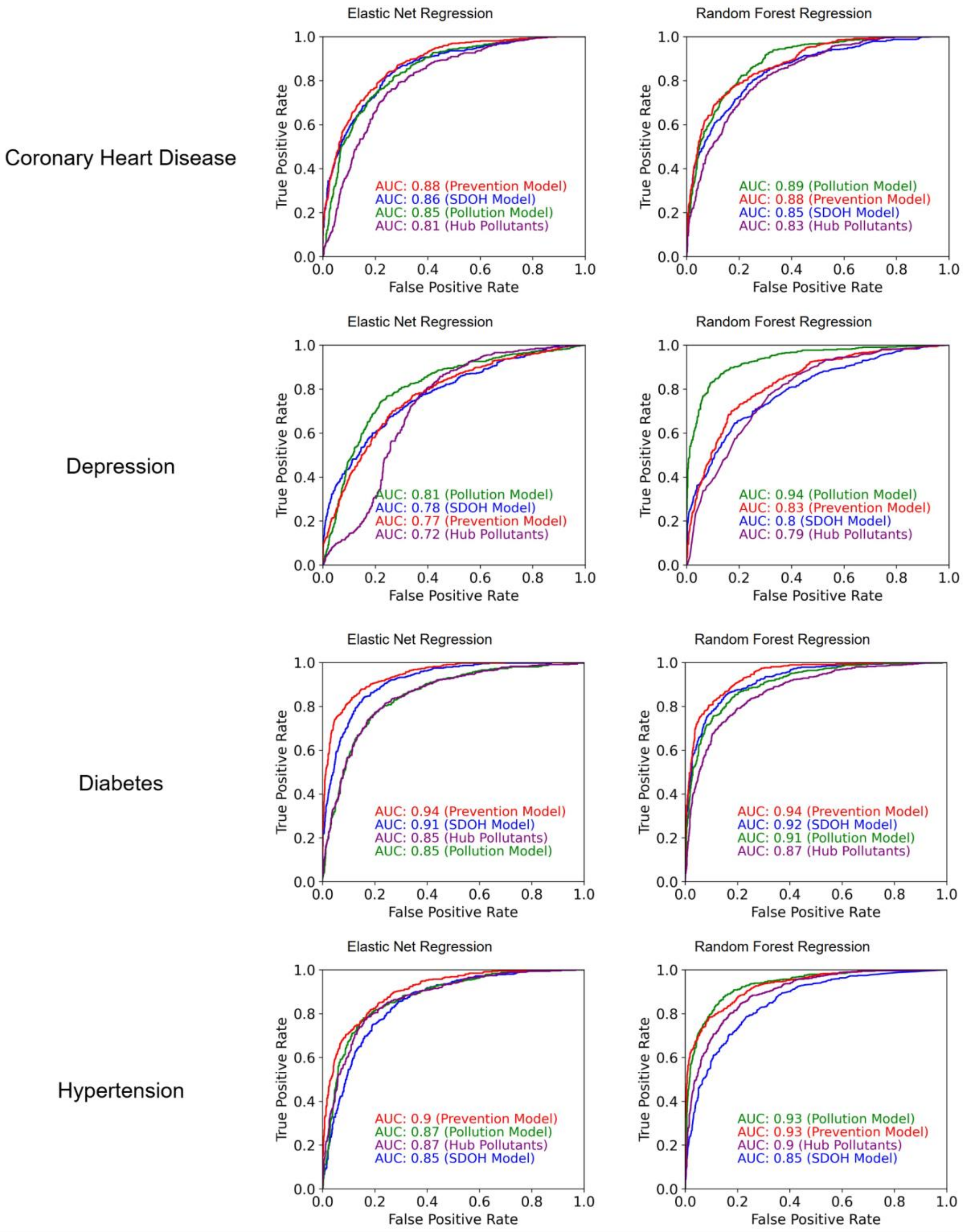

Coronary Heart Disease
Elastic Net Regression
AUC: 0.88 (Prevention Model)
AUC: 0.86 (SDOH Model)
AUC: 0.85 (Pollution Model)
AUC: 0.81 (Hub Pollutants)
Random Forest Regression
AUC: 0.89 (Pollution Model)
AUC: 0.88 (Prevention Model)
AUC: 0.85 (SDOH Model)
AUC: 0.83 (Hub Pollutants)
Depression
Elastic Net Regression
AUC: 0.81 (Pollution Model)
AUC: 0.78 (SDOH Model)
AUC: 0.77 (Prevention Model)
AUC: 0.72 (Hub Pollutants)
Random Forest Regression
AUC: 0.94 (Pollution Model)
AUC: 0.83 (Prevention Model)
AUC: 0.8 (SDOH Model)
AUC: 0.79 (Hub Pollutants)
Diabetes
Elastic Net Regression
AUC: 0.94 (Prevention Model)
AUC: 0.91 (SDOH Model)
AUC: 0.85 (Hub Pollutants)
AUC: 0.85 (Pollution Model)
Random Forest Regression
AUC: 0.94 (Prevention Model)
AUC: 0.92 (SDOH Model)
AUC: 0.91 (Pollution Model)
AUC: 0.87 (Hub Pollutants)
Hypertension
Elastic Net Regression
AUC: 0.9 (Prevention Model)
AUC: 0.87 (Pollution Model)
AUC: 0.87 (Hub Pollutants)
AUC: 0.85 (SDOH Model)
Random Forest Regression
AUC: 0.93 (Pollution Model)
AUC: 0.93 (Prevention Model)
AUC: 0.9 (Hub Pollutants)
AUC: 0.85 (SDOH Model)
True Positive Rate
False Positive Rate

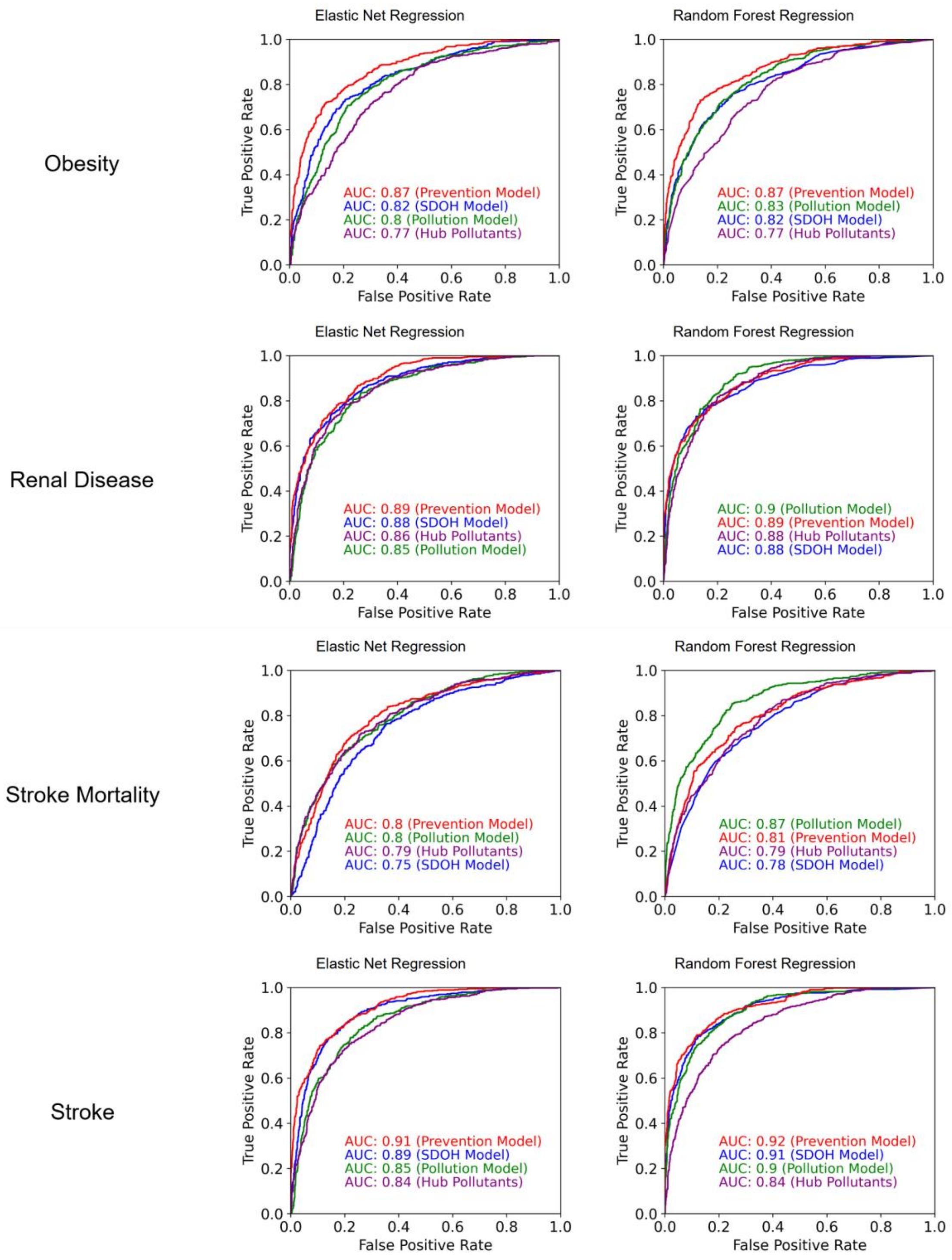


**Supplementary Figure 4.** Receiver-operator (ROC) curves and area under the curve (AUC) values for elastic net and random forest (XGBoost) models predicting whether a county was within (1) or outside (0) a given disease map, where independent variables consisted of (1) preventive healthcare measures, (2) SDOH measures, (3) all pollutants in this study, or (4) just the hub pollutants from aPEER network analysis for a respective disease.

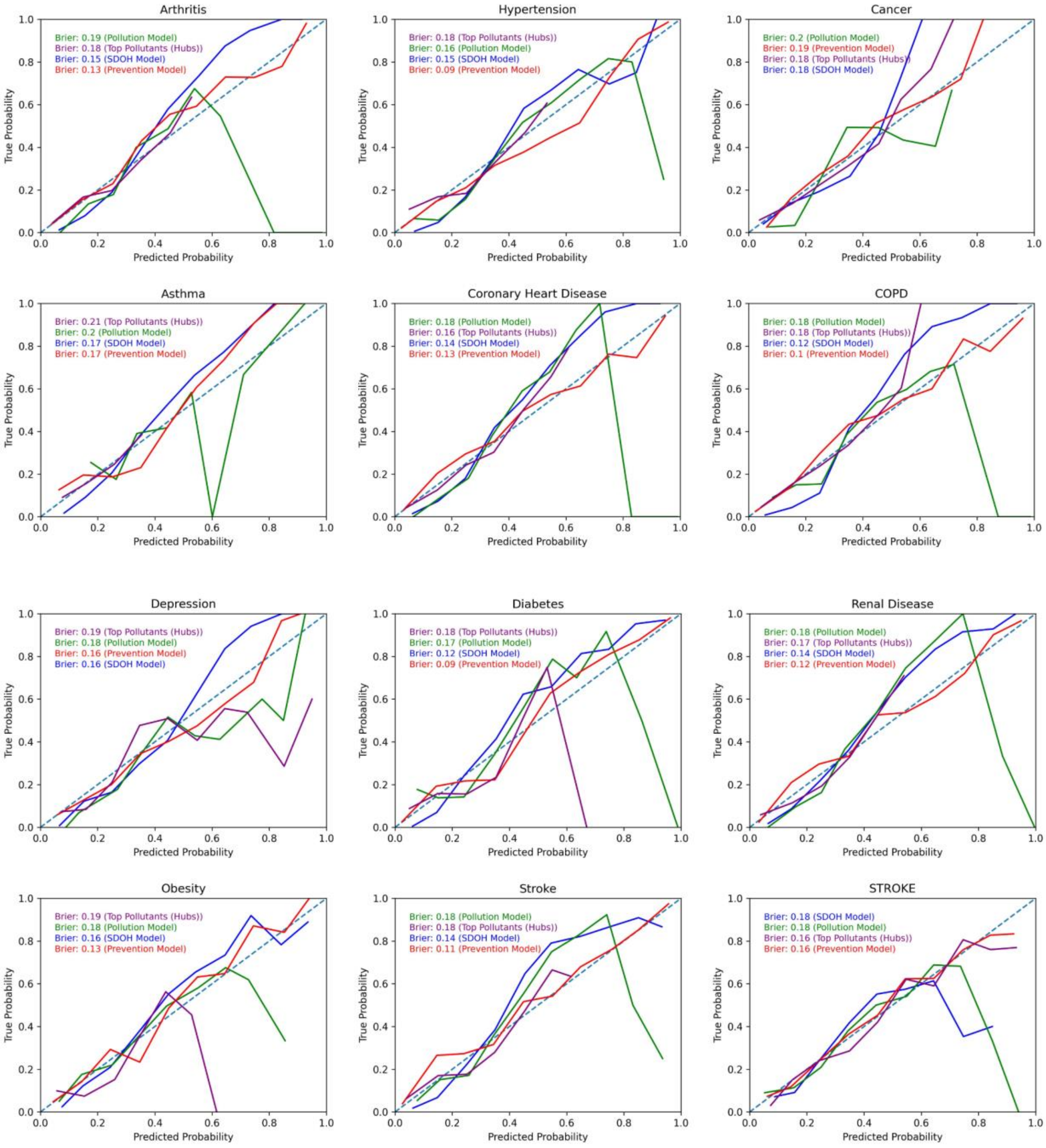


**Supplementary Figure 5A**. Calibration curves for XGBoost random forest models.

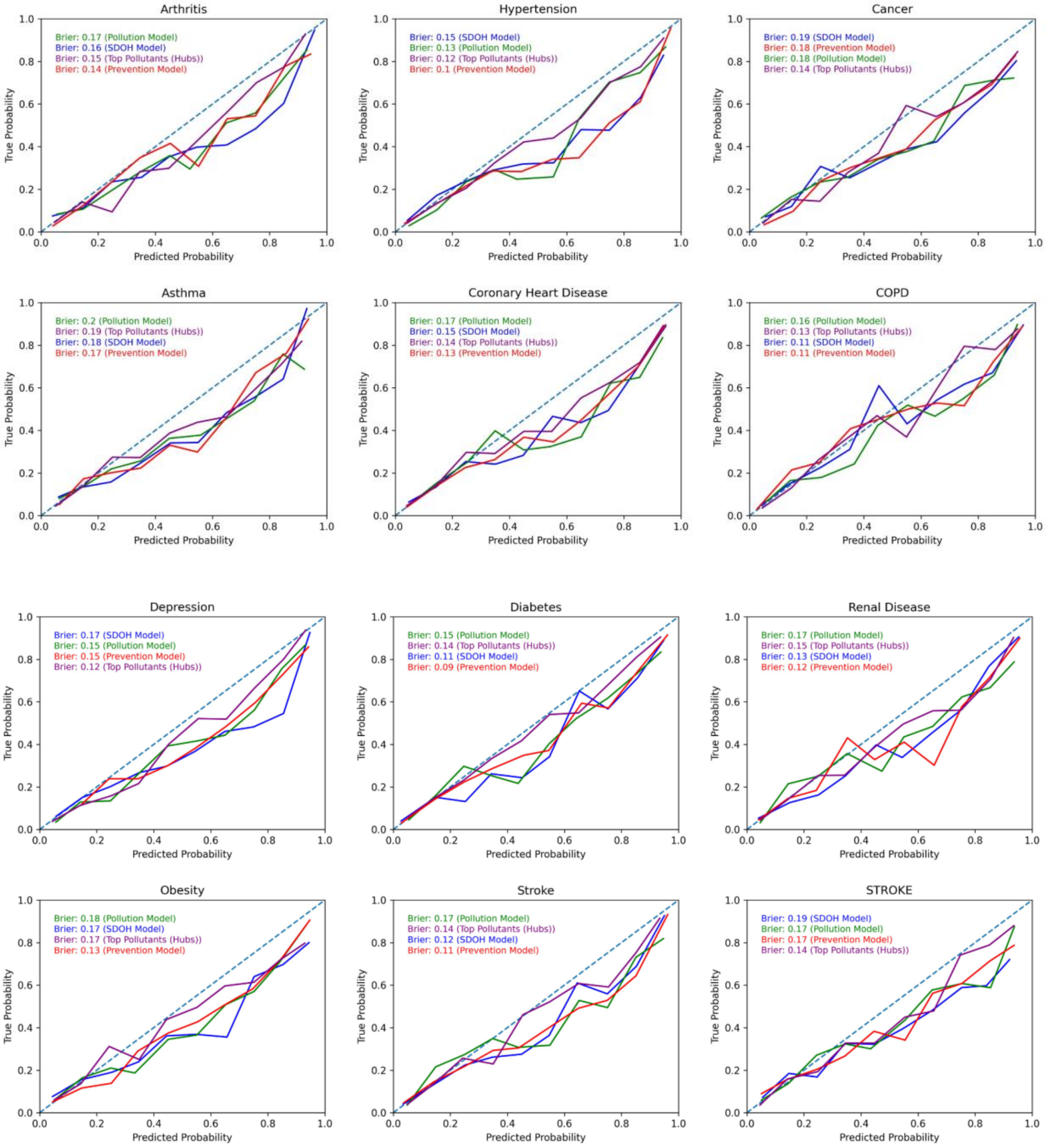


**Supplementary Figure 5B**. Calibration curves for elastic net regression models.

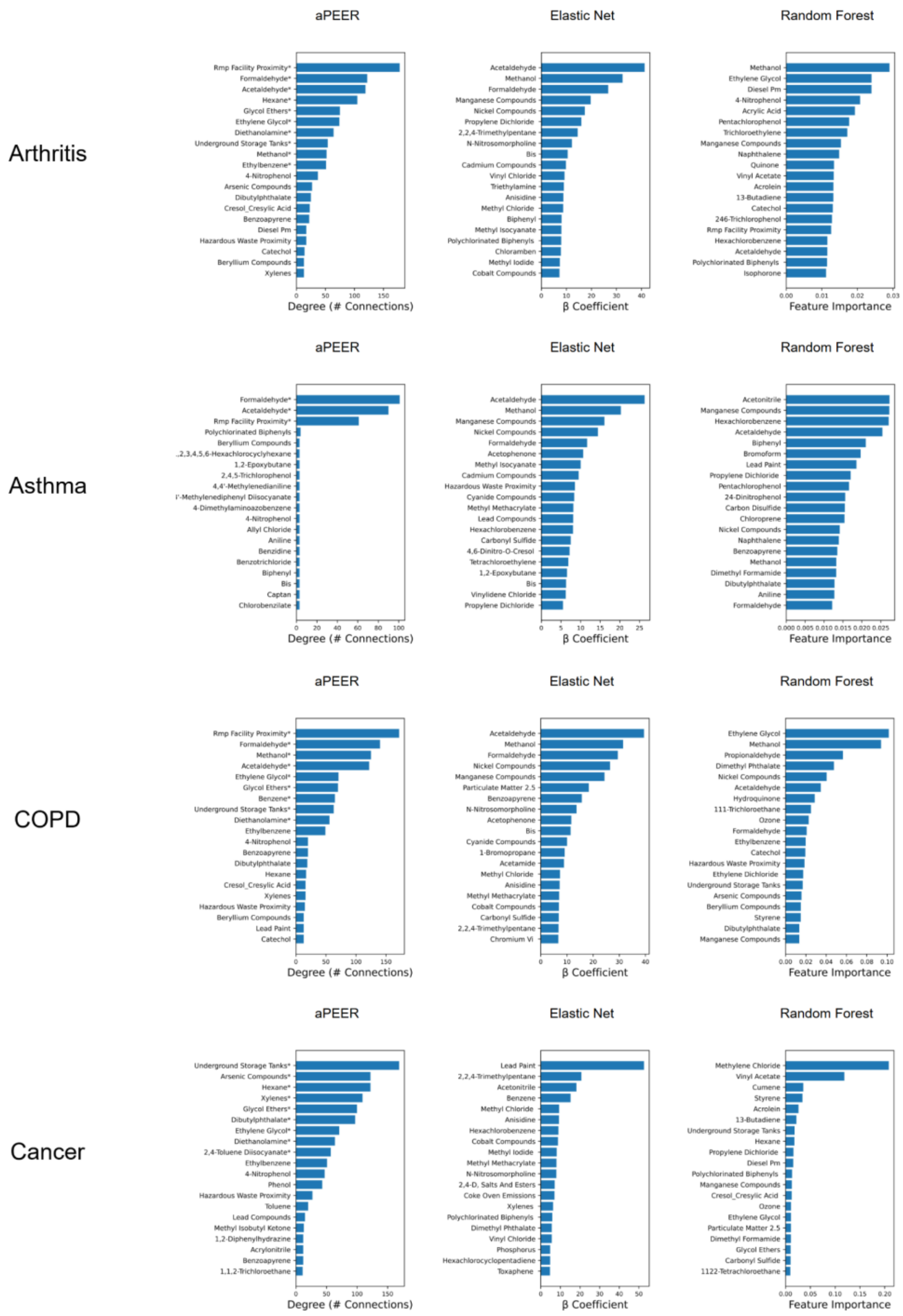
Arthritis
aPEER
Rmp Facility Proximity*
Formaldehyde*
Acetaldehyde*
Hexane*
Glycol Ethers*
Ethylene Glycol*
Diethanolamine*
Underground Storage Tanks*
Methanol*
Ethylbenzene*
4-Nitrophenol
Arsenic Compounds
Dibutylphthalate
Cresol_Cresylic Acid
Benzoapyrene
Diesel Pm
Hazardous Waste Proximity
Catechol
Beryllium Compounds
Xylenes
0 50 100 150
Degree (# Connections)
Elastic Net
Acetaldehyde
Methanol
Formaldehyde
Manganese Compounds
Nickel Compounds
Propylene Dichloride
2,2,4-Trimethylpentane
N-Nitrosomorpholine
Bis
Cadmium Compounds
Vinyl Chloride
Triethylamine
Anisidine
Methyl Chloride
Biphenyl
Methyl Isocyanate
Polychlorinated Biphenyls
Chloramben
Methyl Iodide
Cobalt Compounds
0 10 20 30 40
β Coefficient
Random Forest
Methanol
Ethylene Glycol
Diesel Pm
4-Nitrophenol
Acrylic Acid
Pentachlorophenol
Trichloroethylene
Manganese Compounds
Naphthalene
Quinone
Vinyl Acetate
Acrolein
13-Butadiene
Catechol
246-Trichlorophenol
Rmp Facility Proximity
Hexachlorobenzene
Acetaldehyde
Polychlorinated Biphenyls
Isophorone
0.00 0.01 0.02 0.03
Feature Importance
Asthma
aPEER
Formaldehyde*
Acetaldehyde*
Rmp Facility Proximity*
Polychlorinated Biphenyls
Beryllium Compounds
.,2,3,4,5,6-Hexachlorocyclyhexane
1,2-Epoxybutane
2,4,5-Trichlorophenol
4,4'-Methylenedianiline
'-Methylenediphenyl Diisocyanate
4-Dimethylaminoazobenzene
4-Nitrophenol
Allyl Chloride
Aniline
Benzidine
Benzotrichloride
Biphenyl
Bis
Captan
Chlorobenzilate
0 20 40 60 80 100
Degree (# Connections)
Elastic Net
Acetaldehyde
Methanol
Manganese Compounds
Nickel Compounds
Formaldehyde
Acetophenone
Methyl Isocyanate
Cadmium Compounds
Hazardous Waste Proximity
Cyanide Compounds
Methyl Methacrylate
Lead Compounds
Hexachlorobenzene
Carbonyl Sulfide
4,6-Dinitro-O-Cresol
Tetrachloroethylene
1,2-Epoxybutane
Bis
Vinylidene Chloride
Propylene Dichloride
0 5 10 15 20 25
β Coefficient
Random Forest
Acetonitrile
Manganese Compounds
Hexachlorobenzene
Acetaldehyde
Biphenyl
Bromoform
Lead Paint
Propylene Dichloride
Pentachlorophenol
24-Dinitrophenol
Carbon Disulfide
Chloroprene
Nickel Compounds
Naphthalene
Benzoapyrene
Methanol
Dimethyl Formamide
Dibutylphthalate
Aniline
Formaldehyde
0.000 0.005 0.010 0.015 0.020 0.025
Feature Importance
COPD
aPEER
Rmp Facility Proximity*
Formaldehyde*
Methanol*
Acetaldehyde*
Ethylene Glycol*
Glycol Ethers*
Benzene*
Underground Storage Tanks*
Diethanolamine*
Ethylbenzene
4-Nitrophenol
Benzoapyrene
Dibutylphthalate
Hexane
Cresol_Cresylic Acid
Xylenes
Hazardous Waste Proximity
Beryllium Compounds
Lead Paint
Catechol
0 50 100 150
Degree (# Connections)
Elastic Net
Acetaldehyde
Methanol
Formaldehyde
Nickel Compounds
Manganese Compounds
Particulate Matter 2.5
Benzoapyrene
N-Nitrosomorpholine
Acetophenone
Bis
Cyanide Compounds
1-Bromopropane
Acetamide
Methyl Chloride
Anisidine
Methyl Methacrylate
Cobalt Compounds
Carbonyl Sulfide
2,2,4-Trimethylpentane
Chromium Vi
0 10 20 30 40
β Coefficient
Random Forest
Ethylene Glycol
Methanol
Propionaldehyde
Dimethyl Phthalate
Nickel Compounds
Acetaldehyde
Hydroquinone
111-Trichloroethane
Ozone
Formaldehyde
Ethylbenzene
Catechol
Hazardous Waste Proximity
Ethylene Dichloride
Underground Storage Tanks
Arsenic Compounds
Beryllium Compounds
Styrene
Dibutylphthalate
Manganese Compounds
0.00 0.02 0.04 0.06 0.08 0.10
Feature Importance
Cancer
aPEER
Underground Storage Tanks*
Arsenic Compounds*
Hexane*
Xylenes*
Glycol Ethers*
Dibutylphthalate*
Ethylene Glycol*
Diethanolamine*
2,4-Toluene Diisocyanate*
Ethylbenzene
4-Nitrophenol
Phenol
Hazardous Waste Proximity
Toluene
Lead Compounds
Methyl Isobutyl Ketone
1,2-Diphenylhydrazine
Acrylonitrile
Benzoapyrene
1,1,2-Trichloroethane
0 50 100 150
Degree (# Connections)
Elastic Net
Lead Paint
2,2,4-Trimethylpentane
Acetonitrile
Benzene
Methyl Chloride
Anisidine
Hexachlorobenzene
Cobalt Compounds
Methyl Iodide
Methyl Methacrylate
N-Nitrosomorpholine
2,4-D, Salts And Esters
Coke Oven Emissions
Xylenes
Polychlorinated Biphenyls
Dimethyl Phthalate
Vinyl Chloride
Phosphorus
Hexachlorocyclopentadiene
Toxaphene
0 10 20 30 40 50
β Coefficient
Random Forest
Methylene Chloride
Vinyl Acetate
Cumene
Styrene
Acrolein
13-Butadiene
Underground Storage Tanks
Hexane
Propylene Dichloride
Diesel Pm
Polychlorinated Biphenyls
Manganese Compounds
Cresol_Cresylic Acid
Ozone
Ethylene Glycol
Particulate Matter 2.5
Dimethyl Formamide
Glycol Ethers
Carbonyl Sulfide
1122-Tetrachloroethane
0.00 0.05 0.10 0.15 0.20
Feature Importance

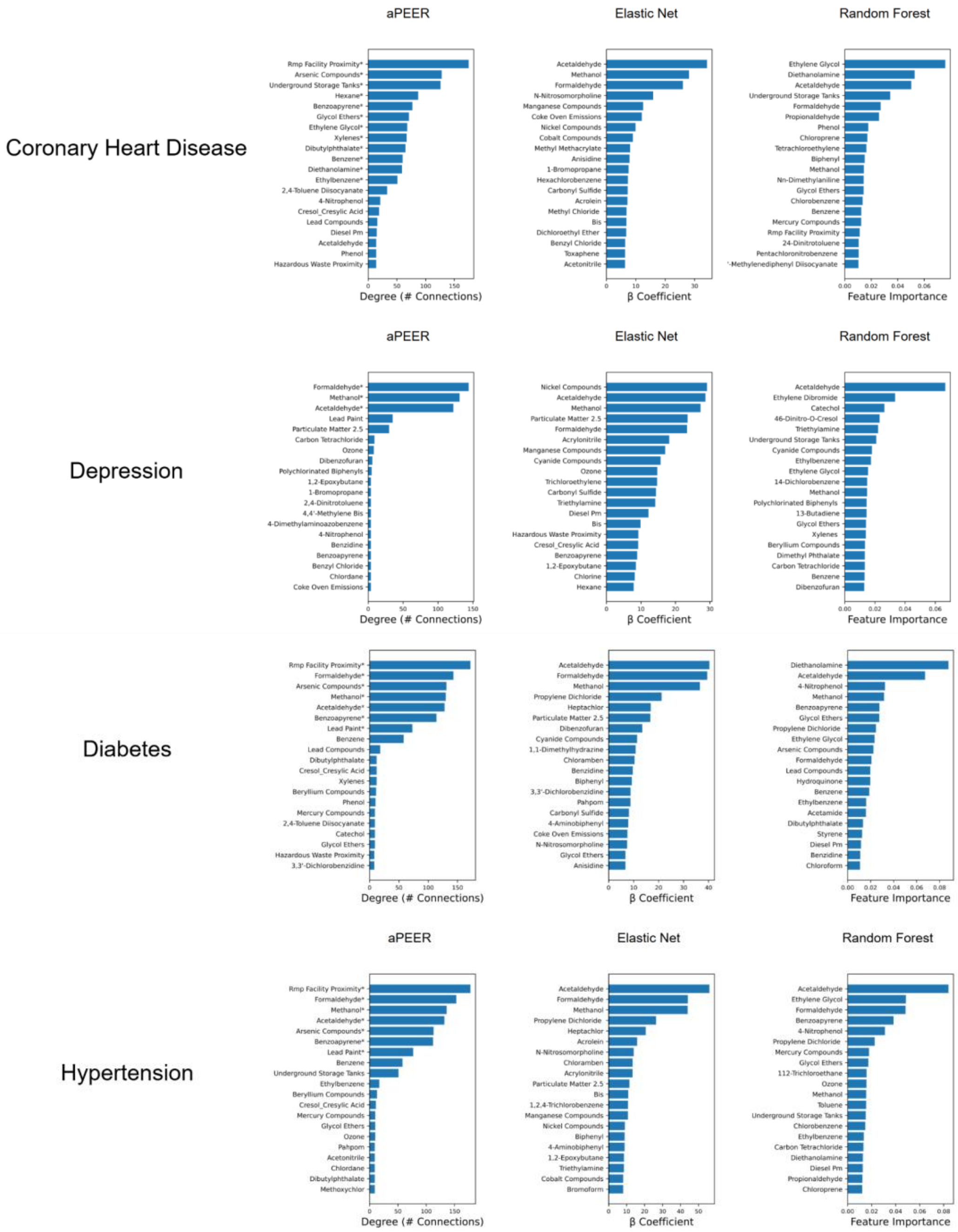
aPEER
Elastic Net
Random Forest
Coronary Heart Disease
Depression
Diabetes
Hypertension
Degree (# Connections)
β Coefficient
Feature Importance

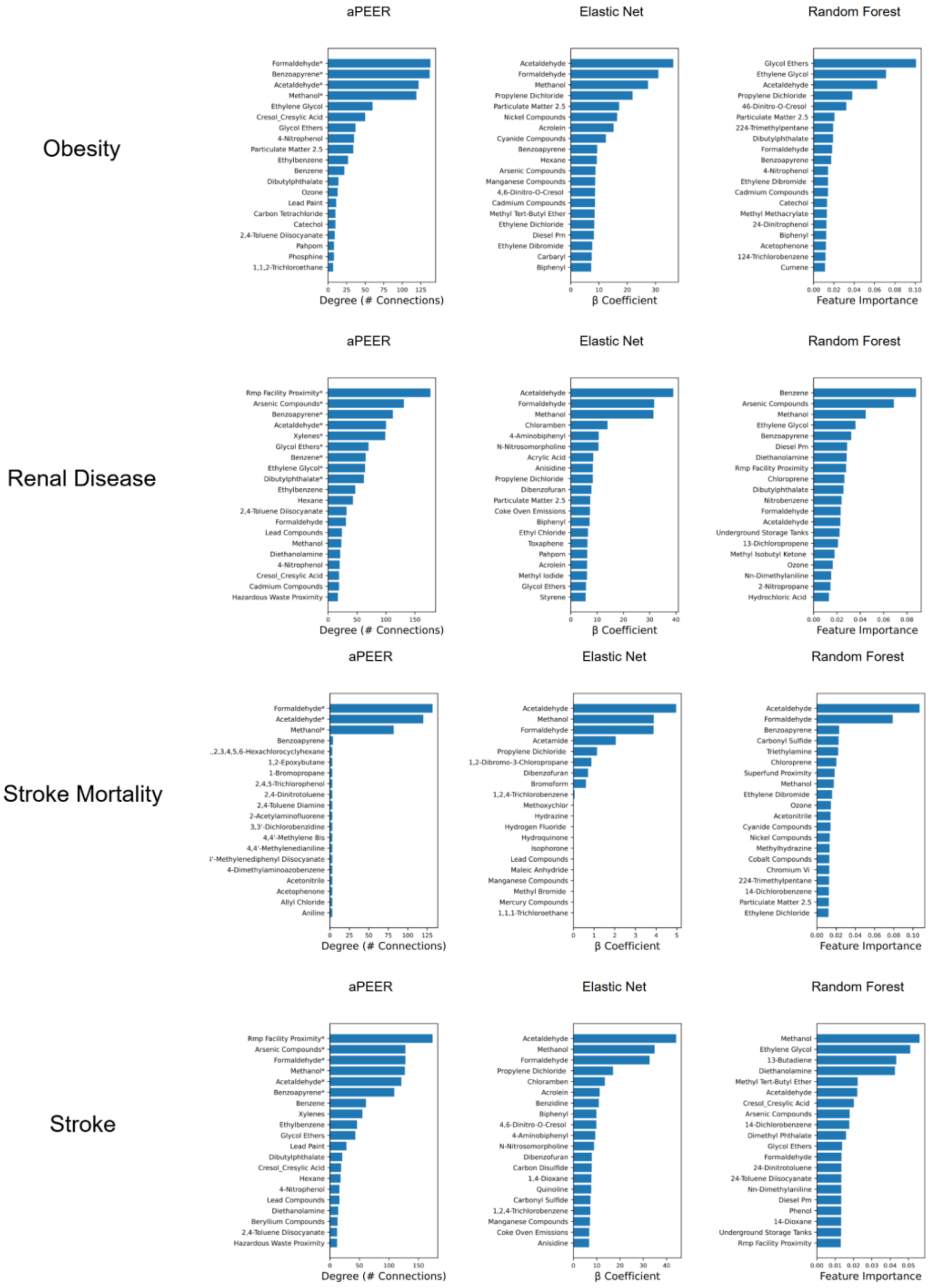


**Supplementary Figure 6.** Comparison of aPEER pollution hubs (left), pollution-associated Elastic Net β coefficients (middle), and random forest-associated pollution feature importance (right) for each disease.

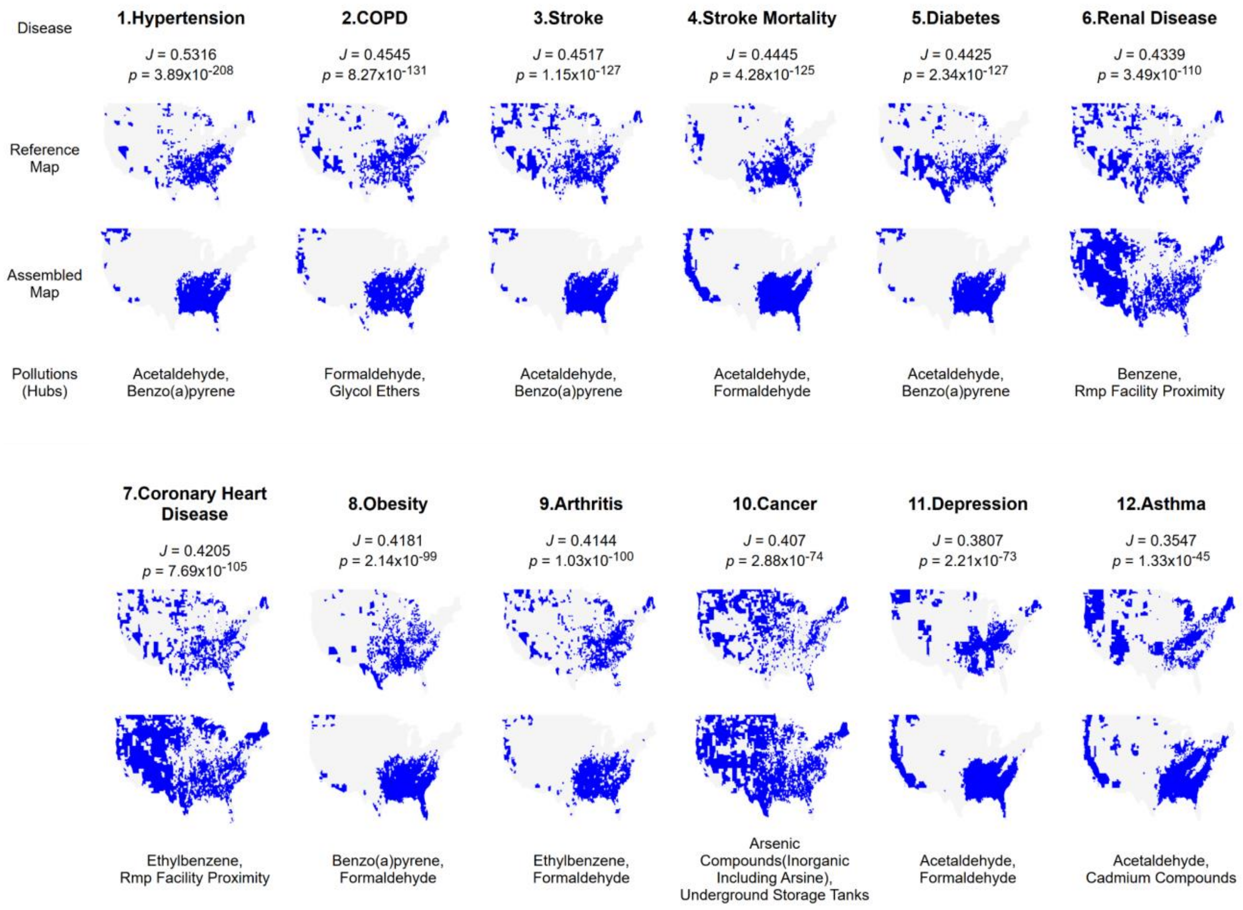


**Supplementary Figure 7.** The strongest disease-pollution associations, ranked by Jaccard correlation coefficient (map assembly derived from the top 2 hub pollutants identified by pairwise Jaccard correlation coefficient).

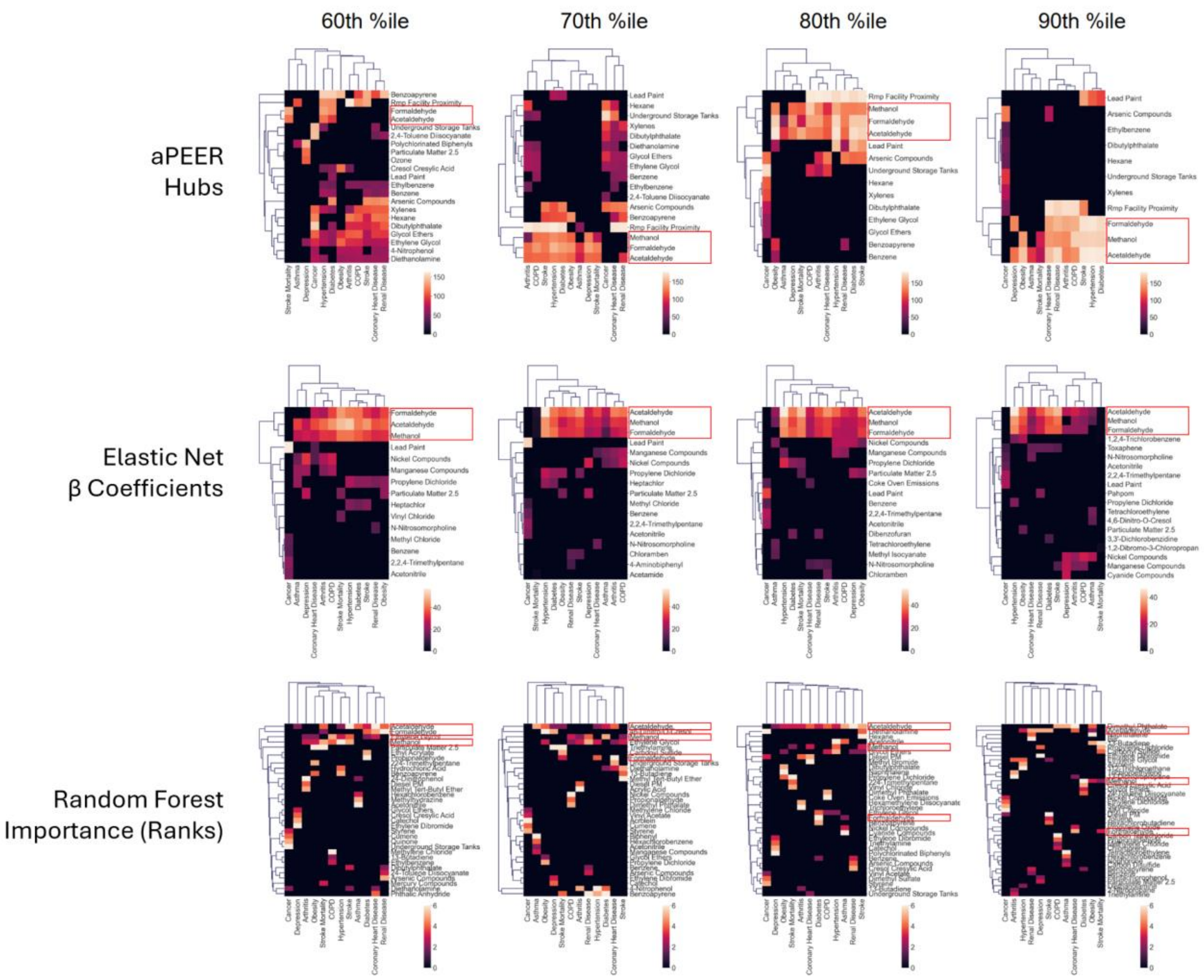


**Supplementary Figure 8**. **Sensitivity analysis of disease-pollution associations.** Clustering the results from aPEER, elastic net, and random forest-derived pollution data at different disease thresholds showed consistent patterns for aPEER and elastic net.

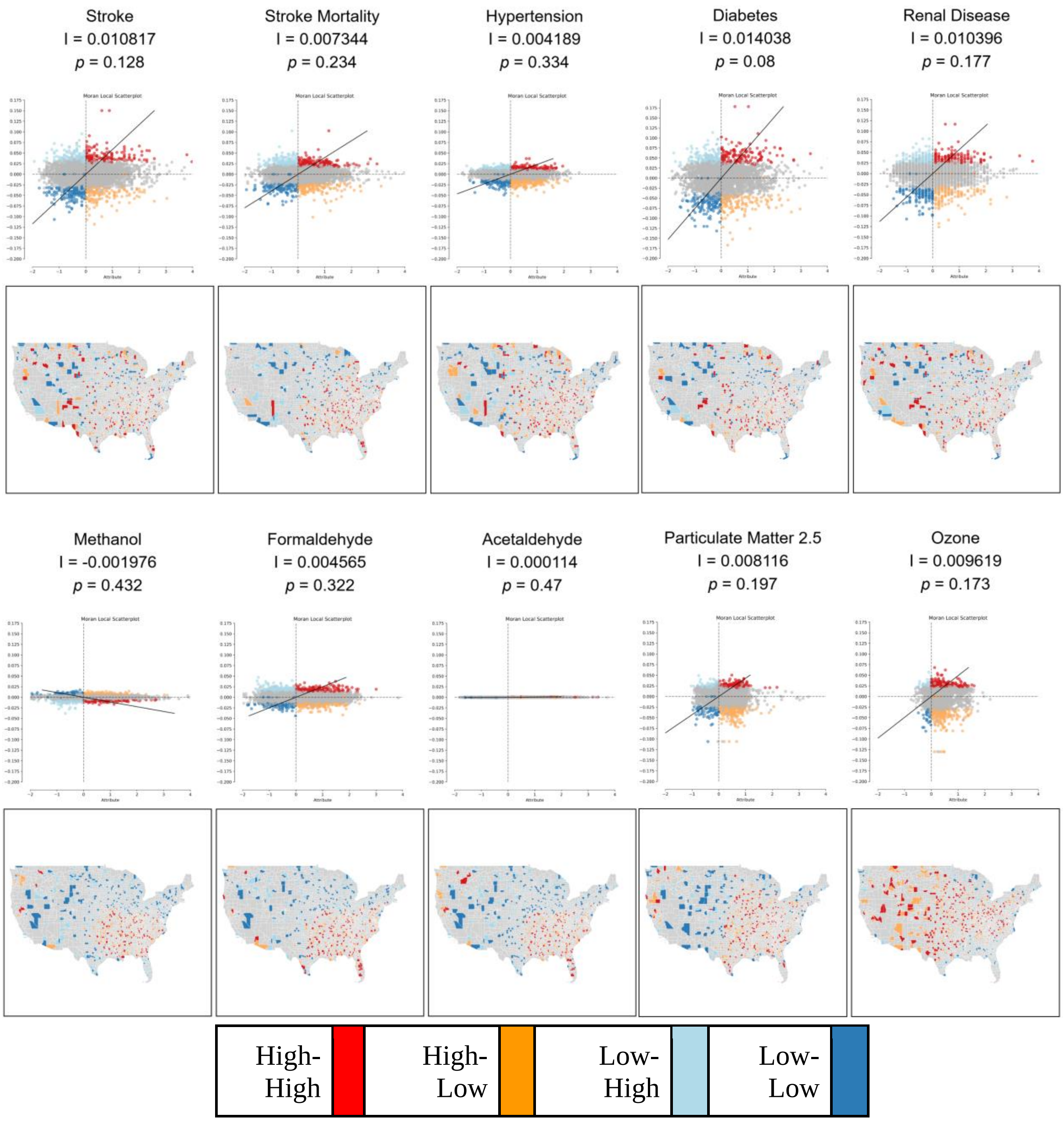


**Supplementary Figure 9**. Spatial analysis benchmarks using Moran's *I* (scatterplots) and LISA (county-level maps of the US). Calculation of Moran's *I* and LISA maps for stroke mortality rate and different pollution indicators. Note that none of the Moran's *I*'s or LISA maps were statistically significant ($p >> 0.1$), and no discernable patterns appeared even with a significantly relaxed *p*-value threshold ($p < 0.1$).

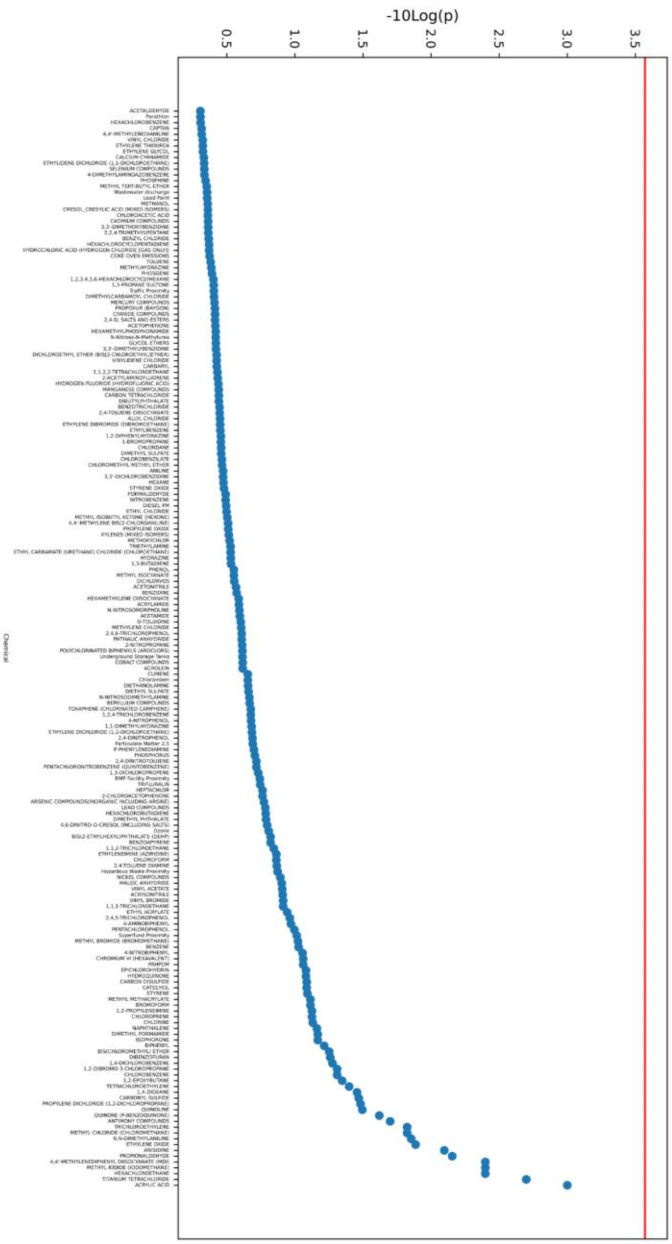


**Supplementary Figure 10. Calculation of Moran's *I* for 186 pollutants.** Manhattan plot of the p-values associated with Moran's I and the Bonferroni-adjusted threshold of statistical significance (red). No pollutants were found to exhibit statistically-significant geospatial clustering.

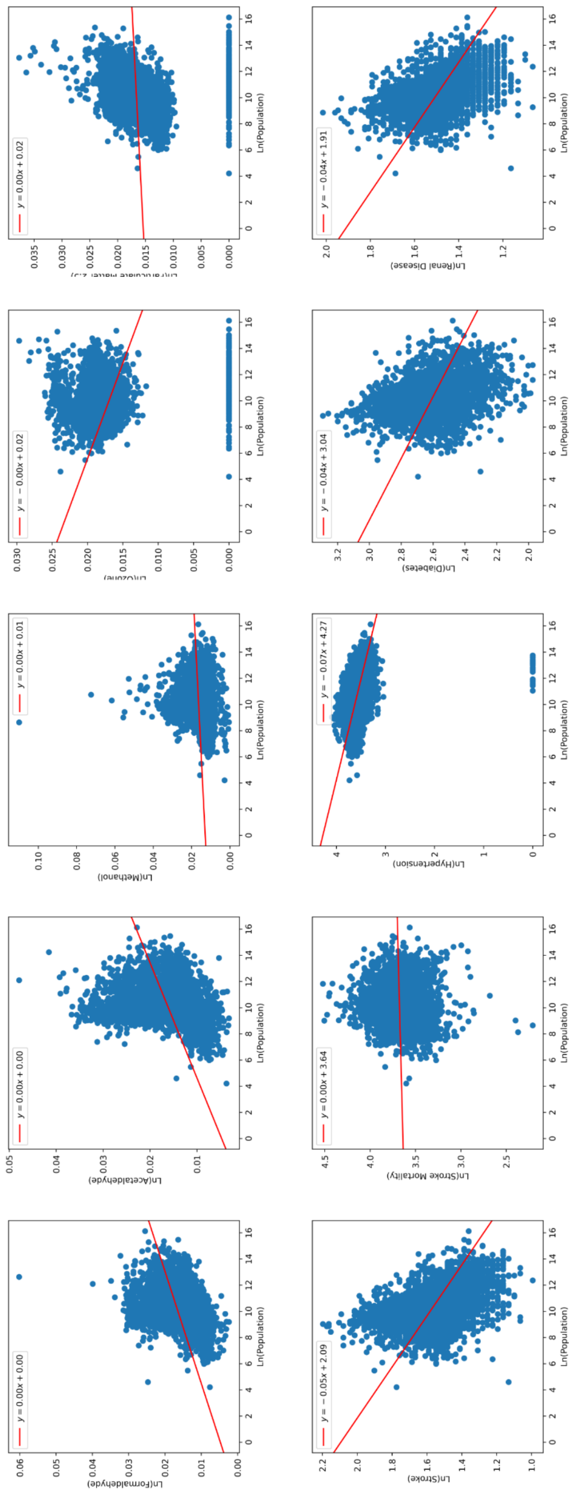


**Supplementary Figure 11**. County-level pollution/population correlation. The correlation between stroke mortality and population at the county level (natural-log transformed).

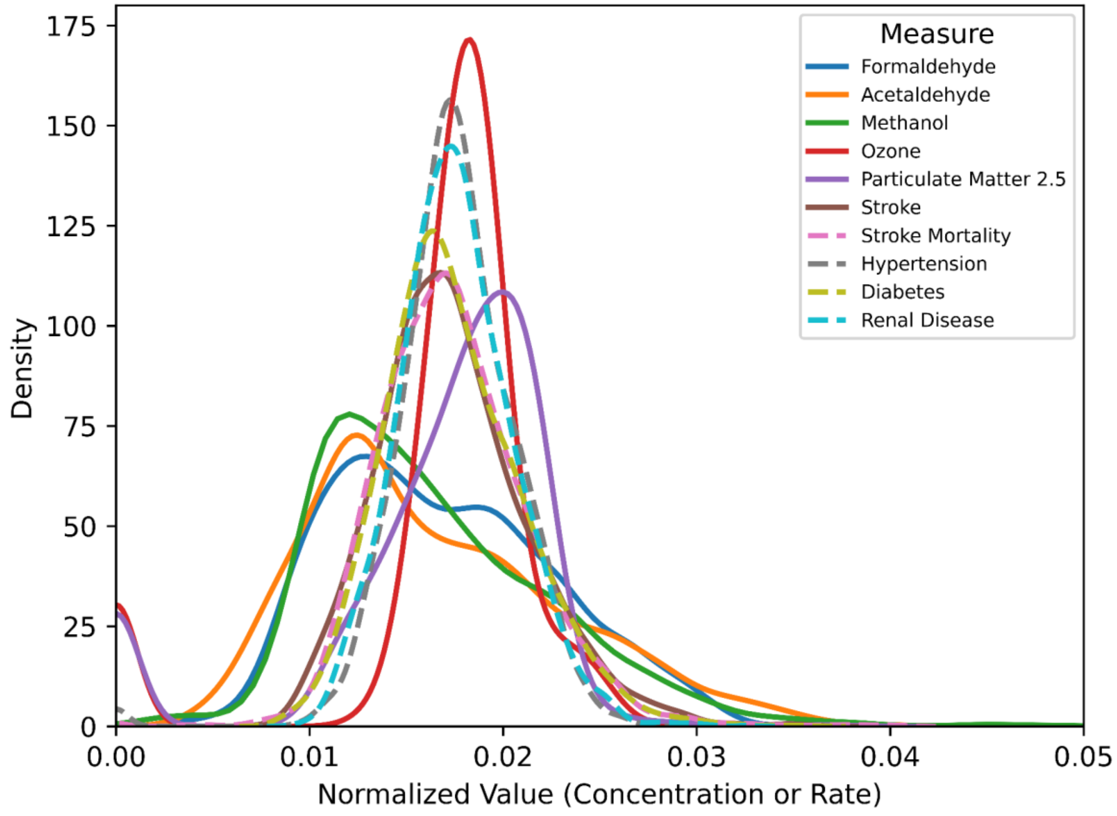


**Supplementary Figure 12**. **County-level pollution density plots.** Density plots with normalized data for selected pollutants and diseases.
